# Impact of COVID-19 lockdown on psychosocial factors, health, and lifestyle in Scottish octogenarians: the Lothian Birth Cohort 1936 Study

**DOI:** 10.1101/2020.10.01.20203711

**Authors:** Adele M. Taylor, Danielle Page, Judith A. Okely, Janie Corley, Miles Welstead, Barbora Skarabela, Paul Redmond, Tom C. Russ, Simon R. Cox

## Abstract

**Background:** Little is known about effects of COVID-19 lockdown on psychosocial factors, health and lifestyle in older adults, particularly those aged over 80 years, despite the risks posed by COVID-19 to this age group.

**Methods:** Lothian Birth Cohort 1936 members, mean age 84 years (SD=0.3), responded to an online questionnaire in May 2020 (*n*=190). We examined responses (experience and knowledge of COVID-19; adherence to guidance; impact on day-to-day living; social contact; self-reported physical and mental health; loneliness; and lifestyle) and relationships between previously-measured characteristics and questionnaire outcomes.

**Results:** Four respondents experienced COVID-19; most had good COVID-19 knowledge (94.7%) and found guidance easy to understand (86.3%). There were modest declines in self-reported physical and mental health, and 48.2% did less physical activity. In multivariable regression models, adherence to guidance by leaving the house less often associated with less professional occupational class (OR=0.71, 95%CI 0.51– 0.98) and poorer self-rated general health (OR=0.62, 95%CI 0.42–0.92). Increased internet use associated with female sex (OR=2.32, 95%CI 1.12–4.86) and higher general cognitive ability (OR=1.53, 95%CI 1.03–2.33). Loneliness associated with living alone (OR=0.15, 95%CI 0.07–0.31) and greater anxiety symptoms (OR=1.76, 95%CI 0.45–1.24). COVID-19 related stress associated with lower emotional stability scores (OR=0.40, 95%CI 0.24–0.62). Decreased physical activity associated with less professional occupational class (OR=1.43, 95%CI 1.04–1.96), and lower general cognitive ability (OR=0.679, 95%CI 0.491–0.931).

**Conclusions:** Characteristics including cognitive function, occupational class, self-rated health, anxiety, and emotional stability, may be related to risk of poorer lockdown-related psychosocial and physical outcomes.

## BACKGROUND

The physical, psychological, and social effects of coronavirus disease 2019 (COVID-19) are unprecedented. Since declaration of a pandemic on 11^th^ March 2020 (1), public health measures have been implemented across the globe to suppress the spread of the virus. In Scotland, lockdown measures introduced on 23^rd^ March 2020 included social and physical distancing, isolation of symptomatic individuals, and restrictions on leaving the home (once daily for essential reasons; Scottish Government, 2020). We are yet to discover the effects of COVID-19 lockdown measures, particularly on older people, who are classed as ‘vulnerable’ and have therefore endured some of the greatest restrictions for the longest period. The current study aimed to examine the impact of the Scottish COVID-19 lockdown on psychosocial factors, health, and lifestyle in older adults aged approximately 84-years from the Lothian Birth Cohort 1936 (LBC1936) study.

Older people are known to be at highest risk of severe illness and death from COVID-19 (3). A prospective cohort study in UK acute care hospitals found the highest proportion of hospitalisations and mortality among those aged 80 and over (4). In Scotland, 77% of all deaths involving COVID-19 to 14^th^ June 2020 were of people aged 75 and over (5). The risk increases for individuals with chronic comorbidities, particularly ageing-related diseases including cardiovascular disease, diabetes, respiratory and chronic pulmonary disease (4,6). Over half of those aged over 80 are estimated to be at high risk due to underlying health conditions (7). Because of this increased risk, those most vulnerable to the virus when lockdown began were asked to ‘shield’, remaining at home and strictly avoiding social contact with anyone outside of their homes for at least 12 weeks.

Effects of lockdown measures on ‘vulnerable’ individuals who remain illness-free are unclear. Social distancing measures inherently limit activities and promote social isolation, potentially to the detriment of physical and mental health (8). In middle-aged and older adults, isolation and loneliness are associated with poor cognitive function, cognitive decline, depression, anxiety, lack of feeling valued, poor physical health including poor cardiovascular function, immunity, and mortality (9–15). There are physical health risks associated with reduced activity during lockdown (16), which warrant consideration given the association between declines in physical fitness and cognitive function (17). Labelling older people as a homogenous group of vulnerable individuals may result in stereotyping or marginalisation (18), and negative consequences of social isolation may be exacerbated by the ‘digital divide’ (8), since older people may disproportionately face barriers to accessing modern technology and information sources. That said, it is possible that many older people are more resilient than commonly portrayed, and have adequate resources to cope well.

Data from older people during the pandemic are surprisingly limited. Studies of this age group are under-represented in COVID-19 literature to date, particularly those which include adults over the age of 80. For example, even in high-quality large-scale studies of COVID-19 with representative samples of hundreds or thousands of participants, the number of individuals sampled over the age of 70 is low, or they are not included. In studies which do include older adults, they often account for only 2-27% of the overall sample (19–22), with results based on fewer than 50 older adults in some cases (23,24). Given the clear risks to older adults of the virus, both in terms of health and the wider psychological, social, and lifestyle impacts resulting from stringent lockdown measures, it is important that the experiences of older adults are well reported. Findings from the general population and past pandemics suggest negative consequences for older people in terms of anxiety and depression (24,25), psychological distress (26–29), and wellbeing (30). At the beginning of UK lockdown, survey participants rated social isolation and practical concerns as being of greater risk to their mental health and wellbeing than fear of contracting COVID-19 (23). Individuals aged over 75 were more than twice as likely to report high anxiety during lockdown compared to those under 24 (31). Physical health may be adversely affected due to the impact of lockdown on behaviours such as sleep (32) and physical activity (33). Furthermore, the experience is likely to vary between individuals based on sociodemographic differences (34–36), physical ability (36), genetics (37), mood and personality (25). One of few studies to report on mostly middle aged and older adults found differences in COVID-19 knowledge, awareness, attitudes, and behaviours across ethnic and socioeconomic groups, and in relation to differing levels of health literacy (22); being unemployed or retired, having poorer health, and having lower health literacy were associated with poorer COVID-19 knowledge and fewer changes to daily routine.

To fully understand the impact of Scotland’s lockdown measures on older people, and inform future interventions in the event of a ‘second wave’ or other health crises, it is important to measure: the ways in which behaviours and routines have been altered; how physical and mental health have been affected; whether some people have fared better than others; and whether there are risk and protective factors associated with these differences. Existing research cohorts are particularly valuable in understanding the impacts of the COVID-19 pandemic, particularly by ‘embedding research on COVID-19 into studies where participants’ mental or cognitive health has previously been ascertained’ (8); this is a key strength of the current study. This study is one of few with a reasonably sized sample of older adults; many others base their findings on the responses of very few older-age participants. We explored the impact of lockdown measures on community-dwelling older adults from the LBC1936 study by linking responses to a COVID-19 questionnaire at age 84 with rich data on cognitive ability, demographics, psychosocial, and health factors previously collected at age 82. The study had two aims. First, to describe responses to the COVID-19 questionnaire. Second, to use bivariate and multivariate analyses to examine relationships between previously collected participant characteristics and psychosocial factors, health and lifestyle during lockdown.

## METHODS

### Participants

Participants were members of the LBC1936 study, a longitudinal study principally investigating non-pathological cognitive and brain ageing. All 1,091 members were born in 1936; most reside in Edinburgh and the surrounding Lothian region of Scotland and took part in the Scottish Mental Survey 1947 (SMS1947; Scottish Council for Research in Education, 1949). Participants were recruited between 2004 and 2007 at mean age 70 years (wave 1; Deary et al., 2007). To date, they have attended four further waves at mean ages 73 (2007-2010, *n=*866), 76 (2011-2013, *n=*697), 79 (2014-2017, *n=*550), and 82 (2017-2019, *n=*431). At each wave, detailed cognitive ability, health, psychosocial, lifestyle, and other data are collected. Information on tracing, recruitment and testing of LBC1936 participants can be found elsewhere (40,41). The current study is based on a subsample of participants (*n*=190) who completed an online COVID-19 questionnaire at mean age 84 (± 0.3) years; this group is referred to as ‘respondents’. Ethical approval was obtained from Multi-Centre Research Ethics Committee for Scotland (MREC/01/0/56; Wave 1), the Lothian Research Ethics Committee (LREC/2003/2/29; Wave 1), and the Scotland A Research Ethics Committee (07/MRE00/58; Waves 2-5). The study complies with Declaration of Helsinki guidelines.

### LBC1936 COVID-19 questionnaire

All LBC1936 participants registered with the study in May 2020 (*n*=454) were invited by letter to take part in an online COVID-19 questionnaire, designed by the LBC1936 team for this study (see supplementary material Appendix 1). Respondents lacking capacity to provide informed consent or unable to complete the questionnaire themselves (*n*=3) were permitted to have assistance (e.g. from guardian or nearest relative). The questionnaire was built using the Qualtrics XM platform, and was live between May 27^th^ and June 8^th^ 2020. The questionnaire took approximately 30 minutes to complete; it consisted of 145 questions examining experience of COVID-19, knowledge and adherence to guidance, impact on day-to-day living, social contact, self-reported physical and mental health, loneliness, and lifestyle factors. Many questions were adapted from other COVID-19 surveys and had Likert-type response scales (22,42); all were optional. Some questions refer to the period ‘since COVID-19 measures were introduced on 23^rd^ March 2020’, hereinafter referred to as ‘lockdown’.

### Measures

#### Questionnaire measures

We examined responses to the COVID-19 questionnaire (experience of COVID-19; knowledge and adherence to guidance; impact on day-to-day living; social contact; self-reported physical and mental health and loneliness; and lifestyle (see supplementary tables 1-6 for the wording of individual items and response options).

#### Covariates

Measures hypothesised to be associated with COVID-19 questionnaire outcomes were selected a priori based on previous associations between these variables and psychosocial factors, health and lifestyle in the LBC1936 cohort. These included: childhood and adulthood occupational social class; age; sex; years of formal full-time education; marital status; living alone; current area of residence; age-11 cognitive ability; Mini-Mental State Examination score (43); fluid cognitive ability ‘gf’; general healthy literacy; chronic comorbidities; undiagnosed diabetes; lung function; grip strength; Townsend Disability Scale Score (44); Body Mass Index (BMI); self-rated general health; emotional stability; extraversion; and conscientiousness. Measurement is described in table 1.

**Table 1:**
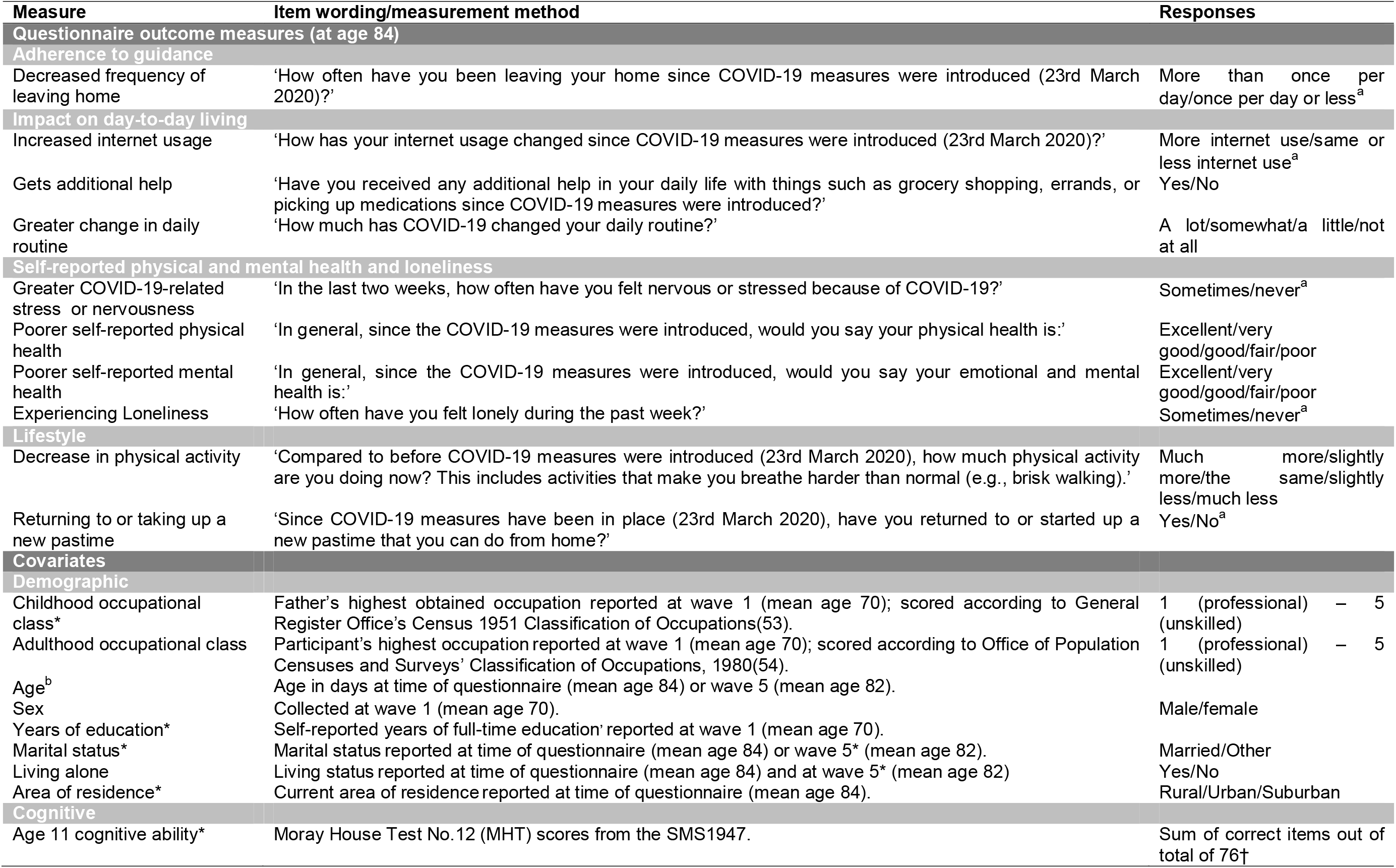
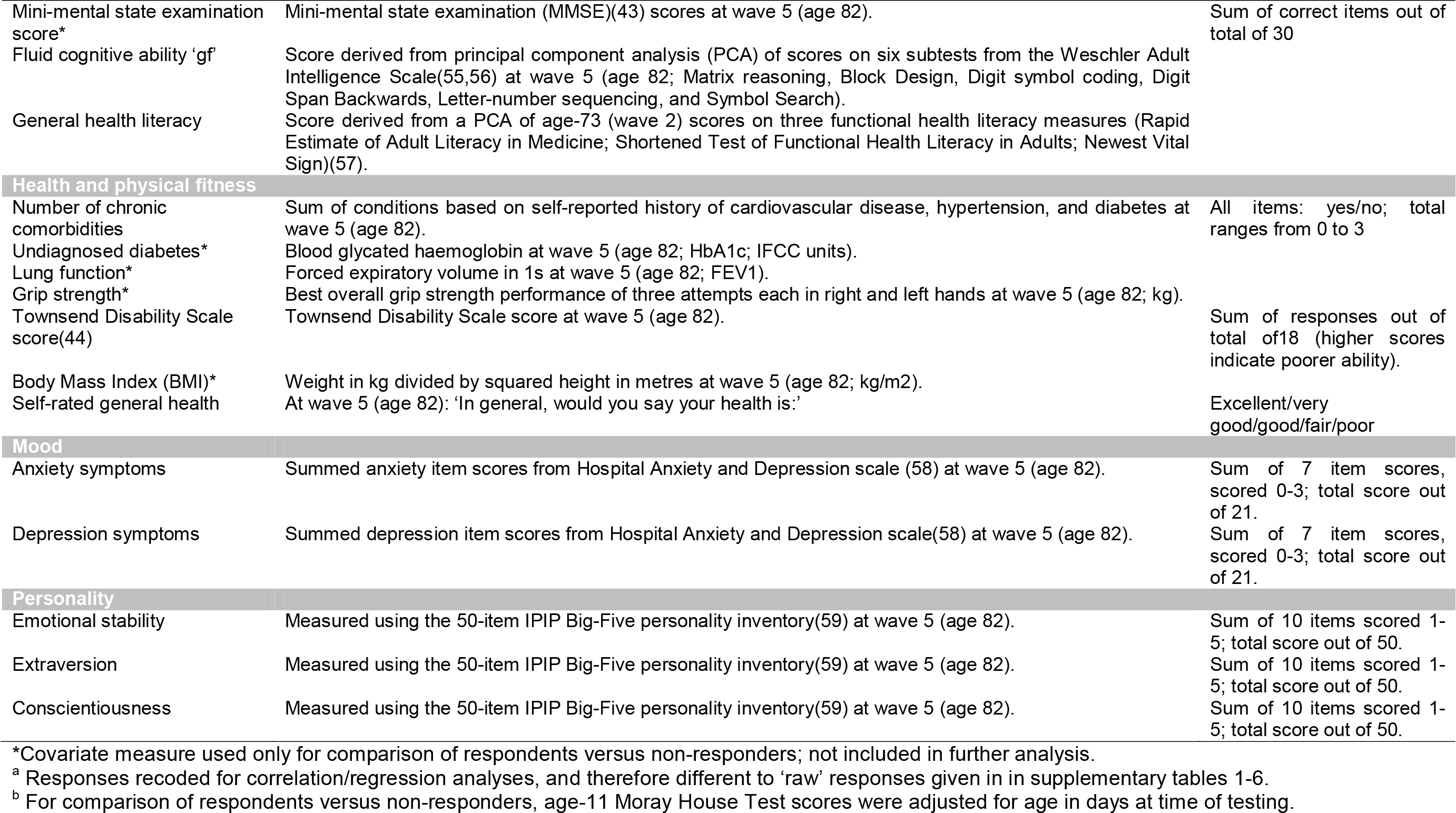
Description of COVID-19 questionnaire outcome measures and covariates used in correlation and regression analyses.

### Statistical analysis

Statistical analyses were conducted using R v3.6.3 (45) and IBM SPSS Statistics v.25 (46). Results of the PCA for covariates gf and general health literacy are presented in supplementary table 7. Descriptive statistics for questionnaire responses were percentages of response relative to number of respondents per questionnaire item (supplementary tables 1-6). An alpha level of .05 was employed for all statistical tests. Welch’s 2-sample t-test, chi-squared tests with Yates’ continuity correction, and Fisher’s exact test were used to compare characteristics of respondents versus non-responders (supplementary table 8). Before undertaking further analysis, respondents who did not attend the most recent wave of LBC1936 testing (wave 5; *n*=8) were excluded, leaving an analytic sample of 182 for inclusion in correlations and regression models. Some outcome measures were recoded from categorical to binary due to low numbers in some response categories; details of outcome measures for correlations and regressions and response coding are included in table 1.

We conducted exploratory bivariate Spearman’s rank correlations to identify relationships between individual differences in previously measured characteristics and 10 COVID-19 questionnaire outcomes from the subthemes: adherence to guidance, impact on day-to-day living, self-reported physical and mental health and loneliness, and lifestyle. We report significant correlations after adjustment for multiple comparisons using Holm-Bonferroni correction (47). Variables that were significantly correlated with COVID-19 questionnaire outcomes were included in binary or ordinal logistic regression models to examine their relative importance and to adjust for potential confounding. All models were adjusted for age and sex. Other covariates varied by outcome but were grouped into blocks by variable type and consistently entered in the following order across models: age and sex, demographics, cognitive ability, health, mood, personality. We report odds ratios (OR) and confidence intervals (CI) for significant associations in final models after adjustment for all covariates. Associations with p-values <.005 remained significant after correction for multiple testing using false discovery rate (FDR) correction (48). Odds ratios reported for continuous independent variables relate to a 1SD increase.

In an additional exploratory step, we conducted Wilcoxon signed rank tests to test for significant changes between ‘before’ and ‘during’ lockdown ratings for self-reported physical and mental health (reported as part of the online questionnaire). We derived physical and mental health change scores by subtracting ‘before’ from ‘during’ scores, then examined possible correlations with previously measured characteristics to explore potential predictors of change.

## RESULTS

### Comparison of responders and non-responders

Background characteristics of respondents (*n*=190) and non-responders (*n=*264) are presented in supplementary table 8. Respondents were less likely to live alone and tended to have had a more professional occupational status; more years of formal education; higher cognitive ability scores; better physical fitness and self-rated general health; fewer symptoms of anxiety and depression; and higher scores for personality traits emotional stability, extraversion, and conscientiousness (all *p-*values ≤.02; Cohen’s d: 0.25 to 0.69).

### Questionnaire responses

#### Experience of COVID-19

Of 190 respondents, 4 (2.1%) reported a self-diagnosis of COVID-19 based on symptoms (see supplementary figure 1); 13.7% were advised to shield due to an underlying health condition; and 12.6% postponed contacting a medical service or attending a medical appointment due to anxiety about COVID-19 (supplementary table 1).

#### Knowledge and adherence to guidance

The majority (94.7%) rated their COVID-19 knowledge extremely or somewhat good, and 86.3% found Scottish Government COVID-19 guidance extremely or somewhat easy to understand. Almost all followed guidance in relation to leaving the home once daily or less (97.9%), social distancing (98.9%), staying at home (96.8%), hand-washing (97.9%), and self-isolating if suffering COVID-19 symptoms (88.6%) all or most of the time. 70.5% said they were unlikely to accidentally come into close contact with someone not in their household (i.e. less than 2 metres) when leaving their home (supplementary table 2). Most (94.1%) followed COVID-19-related news daily; the BBC was the most frequently used source and was rated most helpful (supplementary figures 2 and 3).

#### Living situation and impact on day-to-day living

Over one-third of respondents (38.4%) were living alone and 56.3% were living with a partner during lockdown. 60.0% lived in a suburban area, and almost all had access to a shared or private garden (91.9%). Almost three-quarters (73.8%) reported change in their daily routine during lockdown. Nearly two-thirds (62.6%) received help from others during lockdown, and 64.2% changed their prescription or method of ordering in order to continue to access prescribed medicines during lockdown. Half of respondents were aware of local initiatives to help those self-isolating (51.6%), whereas 42.1% did not know. Nearly two-thirds used more non-cash alternatives during lockdown, and 35.3% said using cash was important. 54.5% used the internet more often during lockdown and 37.1% thought they would continue to do so after the COVID-19 emergency (supplementary table 3).

#### Social contact

Compared to before lockdown, respondents had less face-to-face contact with friends and family members during lockdown, but more regular telephone calls, video calls, and text or instant messages (supplementary figures 4 and 5). Over one-third (33.7%) had more contact with their neighbours during lockdown; 19.5% had less contact; of 101 who reported a change, 62.4% rated this change positively, 31.7% neutral, and 5.9% negatively (supplementary table 4).

#### Self-reported physical and mental health and loneliness

In total, 55.8% rated their physical health before lockdown as being either excellent or very good; this fell to 47.8% during lockdown (figure 1). Before lockdown, 85.1% rated their emotional and mental health as being either excellent or very good; this fell to 68.6% during lockdown (figure 2). Over one-third (36.5%) of respondents felt nervous or stressed because of COVID-19, and less than one quarter (23.8%) felt lonely during lockdown (supplementary table 5).

**Figure 1:**
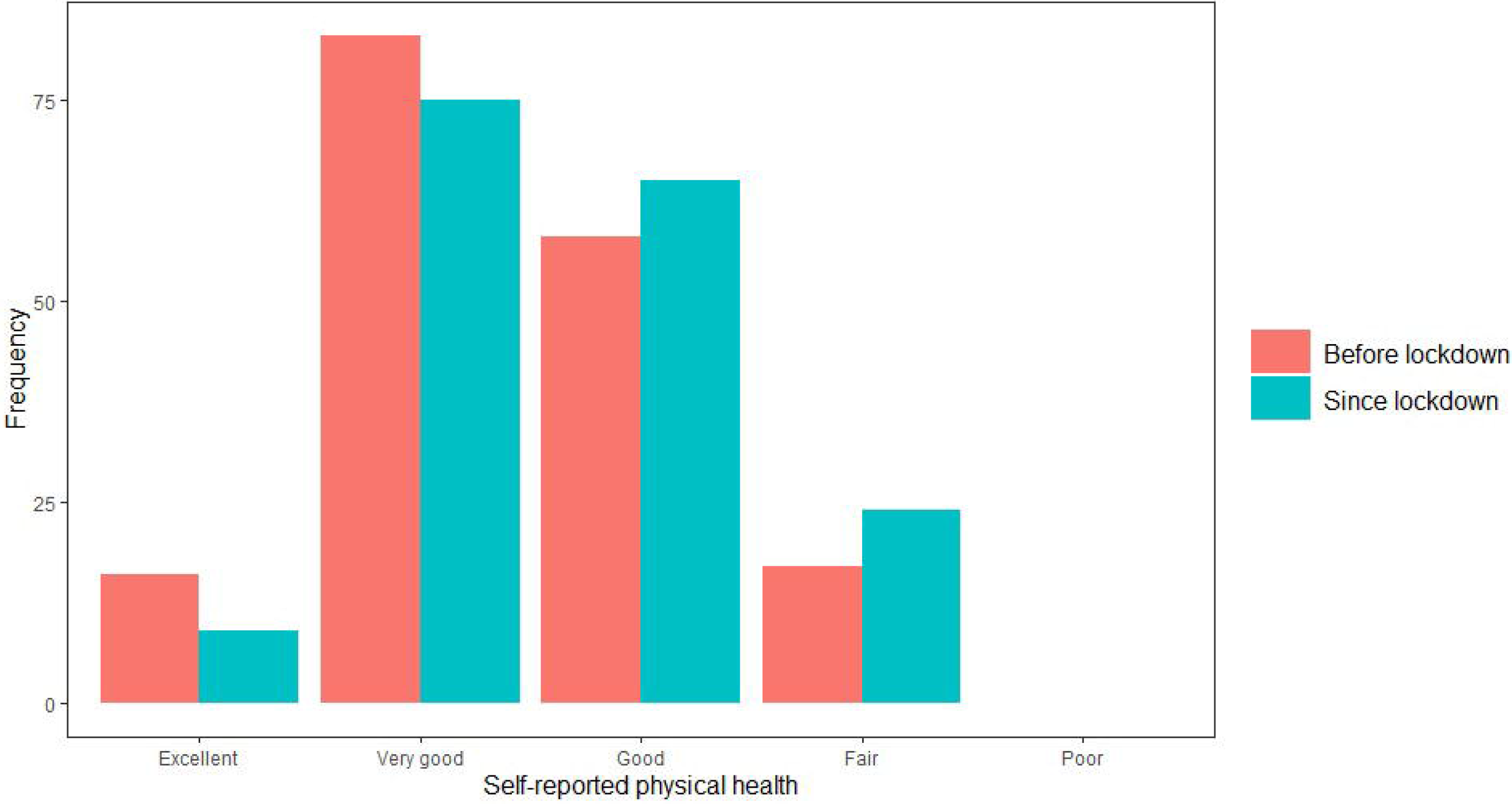
Change in LBC1936 participants’ self-reported physical health after COVID-19 measures introduced.

**Figure 2:**
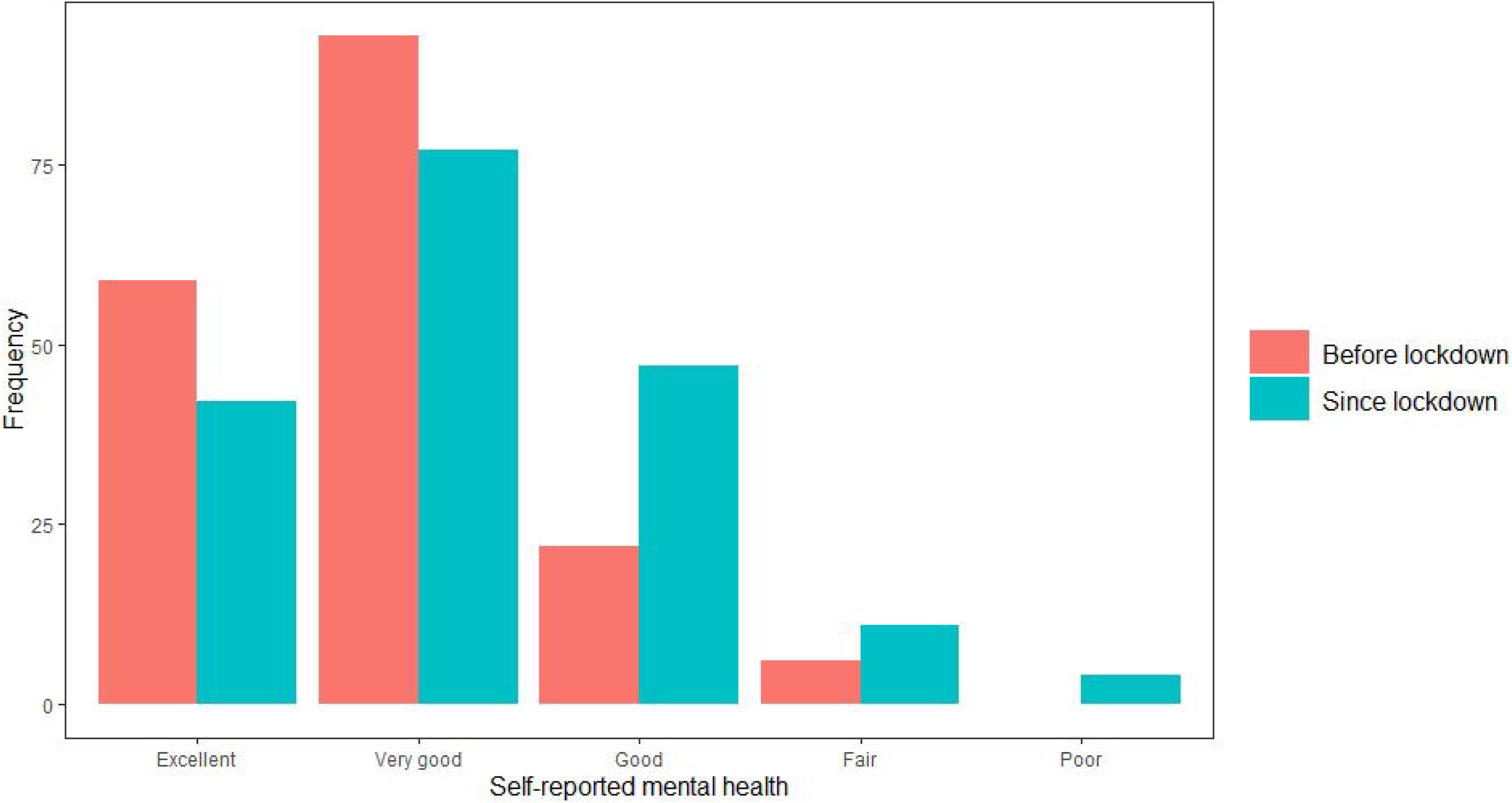
Change in LBC1936 participants’ self-reported emotional and mental health after COVID-19 measures introduced.

#### Lifestyle factors

Of 121 respondents who drink alcohol, 11.7% consumed more alcohol during lockdown; 24.2% consumed less. There were 75 (39.7%) ex-smokers, 2 (1.1%) current smokers, and 112 (59.3%) had never smoked. Few reported a change in diet during lockdown: 18.5% had a healthier diet; 7.9% had a less healthy diet; 10.1% were eating more; and 12.7% were eating less. Almost half of respondents (48.2%) reported doing less physical activity during lockdown, whereas 17.5% did more, and 34.4% did the same amount. Over half of respondents (62.6%) returned to an old pastime or started a new one during lockdown (supplementary table 6). Of 18 pastimes, the most popular were reading (65.3%), watching films or television (63.2%), and gardening (54.0%; figure 3).

**Figure 3:**
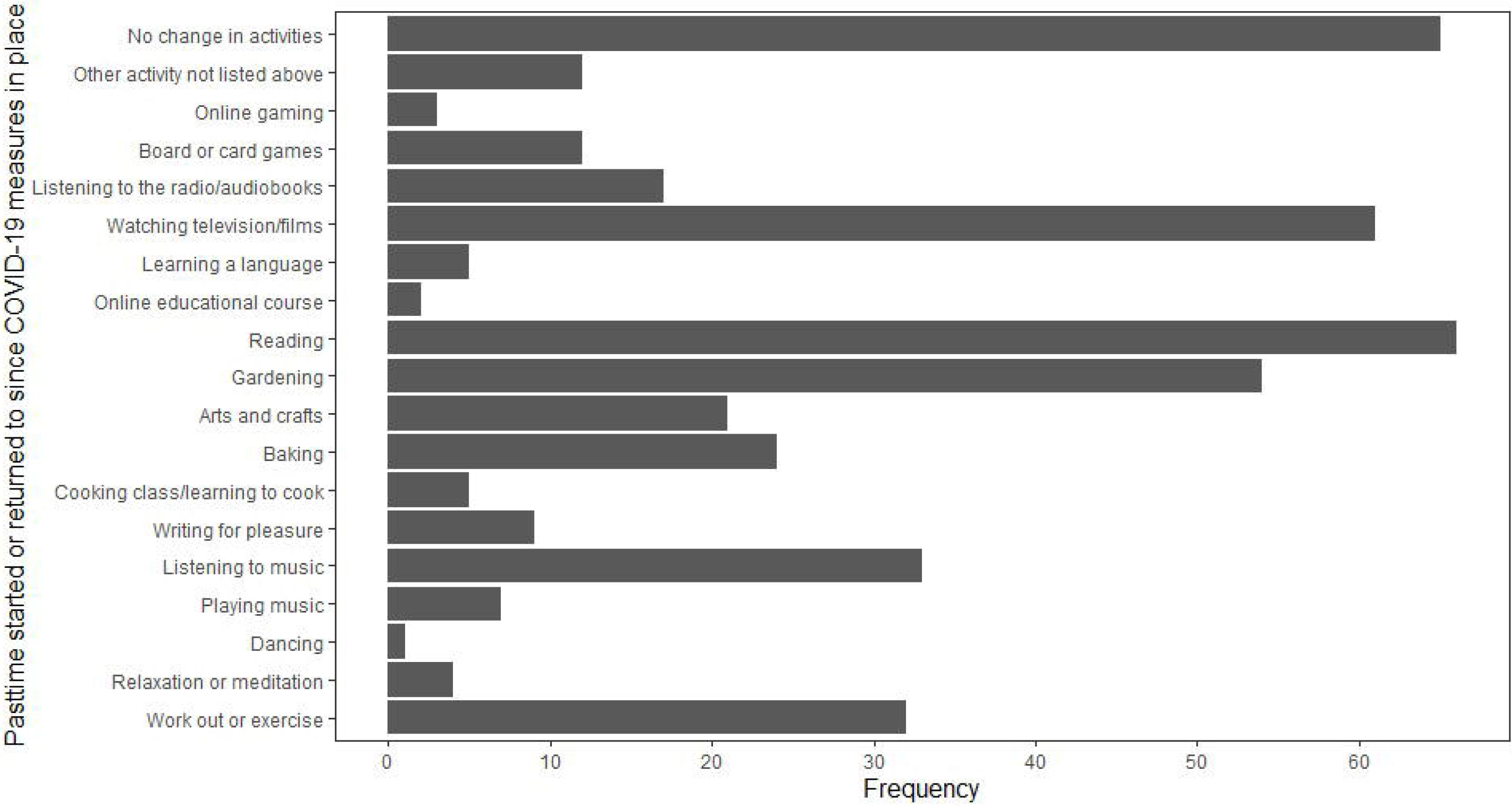
Pastimes returned to or taken up during lockdown.

### Correlations between characteristics at age 82 (or earlier) and COVID-19 outcomes at age 84

Spearman’s rank correlations for the analytical sample (*n*=182) are presented in table 2. *Adherence to guidance*: Leaving home less frequently during lockdown was correlated with more less professional occupational class (r=-0.18), more chronic diseases (r=-0.15), higher Townsend Disability Scale score (r=-0.16), poorer self-rated general health (r=-0.27), and lower gf (r=0.19) at age-82. *Impact on day-to-day living*: Using the internet more often during lockdown was correlated with being female (r=-0.23), currently living alone (r=0.20), higher age-82 gf (r=0.16), and greater anxiety symptoms (r=-0.20). Change in daily routine was correlated with not living alone (r=0.16) and higher age-82 general health literacy (r=-0.20). No variables correlated with getting additional help during lockdown. *Self-reported physical and mental health and loneliness*: poorer self-reported physical health during lockdown was correlated with being older (r=0.18) and male (r=-0.20), lower gf (r=-0.27), more chronic diseases (r=0.22), higher Townsend Disability Scale score (r=0.32), poorer self-rated general health (r=0.52), greater anxiety (r=0.17) and depression symptoms (r=0.33), and lower emotional stability (r=-0.29), conscientiousness (r=-0.20), and extraversion (r=-0.25) at age-82. Poorer self-reported mental health during lockdown was correlated with currently living alone (r=-0.16), more chronic diseases (r=0.15), poorer self-rated general health (r=0.32), greater anxiety (r=0.36) and depression symptoms (r=0.26), and lower emotional stability (r=-0.43), and extraversion (r=-0.22) at age-82. COVID-19 related stress or nervousness during lockdown was correlated with currently living alone (r=-0.19), greater anxiety symptoms (r=0.28), and lower emotional stability at age-82 (r=-0.39). Feeling lonely during lockdown was correlated with living alone (r=-0.40), higher Townsend Disability Scale score (r=0.17), poorer self-rated general health (r=0.16), greater anxiety symptoms (r=0.31), and lower emotional stability at age-82 (r=-0.29). *Lifestyle*: Doing less physical activity during lockdown was correlated with having a less professional occupational class (r=0.22) and lower age-82 gf (r=-0.26). Returning to an old pastime or starting a new one during lockdown was correlated with being female (r=-0.18) and higher age-73 general health literacy (r=-0.16).

**Table 2.** Spearman’s rank correlations between age-82 (or earlier) characteristics and COVID-19 questionnaire outcomes

|  | Adherence to guidance | Impact on day-to-day living |  |  | Self-reported physical and mental health and loneliness |  |  |  | Lifestyle |  |
| --- | --- | --- | --- | --- | --- | --- | --- | --- | --- | --- |
|  | Frequency of leaving home | Change in internet usage | Gets additional help | Change in daily routine | Self-reported physical health | Self-reported mental health | COVID-19-related stress or nervousness | Loneliness | Decrease in physical activity | New pastime |
| Age<sup>a</sup> | -0.01 | 0.03 | 0.00 | 0.05 | 0.18* | 0.14 | -0.02 | 0.08 | 0.03 | 0.06 |
| Sex (*n* male) | -0.12 | -0.23** | 0.03 | -0.05 | -0.20** | 0.03 | 0.13 | 0.06 | 0.00 | -0.18* |
| Demographics |  |  |  |  |  |  |  |  |  |  |
| Adulthood occupational class | -0.18* | 0.09 | 0.14 | -0.15 | 0.10 | -0.03 | -0.01 | 0.11 | 0.22** | -0.02 |
| Living alone<sup>b</sup> (*n* yes) | -0.02 | 0.20* | -0.01 | 0.16* | 0.07 | -0.16* | -0.19* | -0.40*** | -0.03 | 0.14 |
| Cognitive |  |  |  |  |  |  |  |  |  |  |
| Fluid cognitive ability 'gf' | 0.19* | 0.16* | 0.02 | -0.13 | -0.27*** | 0.37 | 0.07 | -0.09 | -0.26*** | -0.09 |
| General health literacy | 0.13 | -0.17 | 0.01 | -0.20* | -0.02 | -0.10 | 0.07 | -0.03 | -0.13 | -0.16* |
| Health |  |  |  |  |  |  |  |  |  |  |
| Number of chronic diseases | -0.15* | 0.03 | 0.03 | 0.03 | 0.22** | 0.15* | 0.01 | 0.12 | 0.12 | 0.06 |
| Townsend disability scale score | -0.16* | -0.04 | 0.00 | 0.11 | 0.32*** | 0.14 | 0.05 | 0.17* | 0.07 | 0.05 |
| Self-rated general health | -0.27*** | -0.07 | -0.02 | 0.00 | 0.52*** | 0.32*** | 0.10 | 0.16* | 0.09 | 0.12 |
| Mood |  |  |  |  |  |  |  |  |  |  |
| Anxiety symptoms | -0.05 | -0.20** | 0.13 | -0.01 | 0.17* | 0.36*** | 0.28*** | 0.31*** | -0.01 | -0.09 |
| Depression symptoms | -0.07 | -0.01 | 0.09 | 0.13 | 0.33*** | 0.26*** | 0.12 | 0.11 | -0.03 | 0.09 |
| Personality |  |  |  |  |  |  |  |  |  |  |
| Emotional stability | 0.04 | 0.07 | -0.10 | -0.04 | -0.29*** | -0.43*** | -0.39*** | -0.29*** | 0.00 | 0.01 |
| Conscientiousness | 0.08 | 0.11 | -0.08 | 0.05 | -0.20** | -0.10 | -0.09 | -0.04 | -0.00 | -0.03 |
| Extraversion | 0.09 | -0.06 | 0.05 | -0.05 | -0.25** | -0.22** | -0.03 | -0.09 | -0.14 | -0.12 |

\* $p < .05$ , \*\* $p < .01$ , \*\*\* $p < .001$ ; Independent variables are from age-82 unless otherwise stated. All p-values corrected using Holm-Bonferroni correction(47).
<sup>a</sup> Age is age in days at time of questionnaire (mean age 84).
<sup>b</sup> Living alone at time of questionnaire (mean age 84).

### Regression analyses with age-82 (or earlier) characteristics as independent variables and age-84 COVID-19 questionnaire responses as outcomes

Results of final regression models for each outcome are displayed in table 3; full results of all individual regression models are provided in supplementary tables 9–17. *Adherence to guidance:* Leaving home less frequently during lockdown was associated with a less professional occupational class (OR=0.71, 95%CI 0.51-0.98) and poorer age-82 self-rated general health (OR=0.62, 95%CI 0.42-0.92). *Impact on day-to-day living:* The odds of using the internet more during lockdown were greater for women (OR=2.32, 95%CI 1.12-4.86) and higher age-82 fluid cognitive ability (gf; OR=1.53, 95%CI 1.03-2.33). No measures were significantly associated with change in daily routine in the final model. *Self-reported physical and mental health and loneliness:* Odds of poorer self-reported physical health during lockdown were increased for those who were older (OR=1.45, 95%CI 1.04-2.04), and had poorer age-82 self-rated general health (OR=3.99, 95%CI 2.31-7.11). Odds of poorer self-reported emotional and mental health during lockdown were lower for those with higher emotional stability (OR=0.54, 95%CI 0.35-0.81). Odds of COVID-19-related stress or nervousness during lockdown were lower for those with higher emotional stability trait scores (OR=0.40, 95%CI 0.24-0.62). Odds of being lonely during lockdown were higher for those with greater age-82 anxiety symptoms (OR=1.76, 95%CI 1.01-3.14) and lower for those not living alone (OR=0.15, 95%CI 0.07-0.31). *Lifestyle*: Decreased physical activity was associated with less professional occupational class (OR=1.43, 95%CI 1.04–1.96), and lower general cognitive ability (OR=0.679, 95%CI 0.491–0.931). There were no significant associations with participation in pastimes in fully adjusted models.

**Table 3.** Odds Ratios (95% Confidence Intervals) for final regression models of COVID-19 outcomes predicted by characteristics at age-82 (or earlier)

|  |  | Adherence to guidance | Impact on day-to-day living |  | Self-reported physical and mental health and loneliness |  |  |  | Lifestyle |  |
| --- | --- | --- | --- | --- | --- | --- | --- | --- | --- | --- |
|  |  | Frequency of leaving home<sup>c</sup> | Change in internet usage<sup>c</sup> | Change in daily routine<sup>d</sup> | Self-reported physical health<sup>d</sup> | Self-reported mental health<sup>d</sup> | COVID-19-related stress or nervousness<sup>c</sup> | Loneliness<sup>c</sup> | Decrease in physical activity<sup>d</sup> | New pastime<sup>c</sup> |
| Age<sup>a</sup> |  | 0.92 (0.69-1.23) | 0.97 (0.68-1.38) | 1.16 (0.86-1.57) | 1.45 (1.04-2.04)* | 1.27 (0.94-1.73) | 0.94 (0.66-1.33) | 1.04 (0.68-1.62) | 1.13 (0.85-1.51) | 0.92 (0.66-1.28) |
| Sex | Male | Reference | Reference | Reference | Reference | Reference | Reference | Reference | Reference | Reference |
|  | Female | 0.56 (0.30-1.02) | 2.32 (1.12-4.86)* | 1.08 (0.55-2.12) | 0.56 (0.28-1.11) | 1.11 (0.57-2.16) | 1.55 (0.73-3.31) | 0.48 (0.17-1.26) | 1.08 (0.61-1.89) | 1.89 (0.96-3.75) |
| Demographics |  |  |  |  |  |  |  |  |  |  |
| Adulthood occupational class |  | 0.71 (0.51-0.98)* | - | - | - | - | - | - | 1.43 (1.04-1.96)* | - |
| Living alone<sup>b</sup> | Alone | Reference | Reference | Reference | Reference | Reference | Reference | Reference | Reference | Reference |
|  | Not alone | - | 0.65 (0.38-1.10) | 1.83 (0.92-3.68) | - | 0.53 (0.28-1.02) | 0.65 (0.38-1.11) | 0.15 (0.07-0.31)***† | - | - |
| Cognitive |  |  |  |  |  |  |  |  |  |  |
| Fluid cognitive ability 'gf' |  | 1.24 (0.88-1.74) | 1.53 (1.03-2.33)* | - | 0.73 (0.50-1.05) | - | - | - | 0.68 (0.49-0.93)* | - |
| General health literacy |  | - | - | 0.69 (0.46-1.01) | - | - | - | - | - | 1.36 (0.92-2.02) |
| Health |  |  |  |  |  |  |  |  |  |  |
| Number of chronic diseases |  | 0.87 (0.60-1.26) | - | - | 0.98 (0.65-1.47) | 1.20 (0.82-1.75) | - | - | - | - |
| Townsend disability scale score |  | 0.73 (0.46-1.13) | - | - | 1.31 (0.74-2.37) | - | - | 1.67 (0.92-3.14) | - | - |
| Self-rated general health<sup>c</sup> |  | 0.62 (0.42-0.92)* | - | - | 3.99 (2.31-7.11)***† | 1.48 (0.99-2.24) | - | - | - | - |
|  | Excellent | - | - | - | - | - | - | 0.17 (0.001-26.69) | - | - |
|  | Very good | - | - | - | - | - | - | 0.64 (0.01-65.46) | - | - |
|  | Good | - | - | - | - | - | - | 0.34 (0.003-33.48) | - | - |
|  | Fair | - | - | - | - | - | - | 3.32 (0.03-399.82) | - | - |
|  | Poor | Reference | Reference | Reference | Reference | Reference | Reference | Reference | Reference | Reference |
| Mood |  |  |  |  |  |  |  |  |  |  |
| Anxiety symptoms |  | - | 1.31 (0.92-1.90) | - | 0.84 (0.54-1.30) | 1.15 (0.76-1.73) | 0.99 (0.63-1.55) | 1.76 (1.01-3.14)* | - | - |
| Depression symptoms |  | - | - | - | 1.17 (0.77-1.78) | 1.03 (0.71-1.50) | - | - | - | - |
| Personality |  |  |  |  |  |  |  |  |  |  |
| Emotional stability |  | - | - | - | 0.81 (0.51-1.26) | 0.54 (0.35-0.81)**† | 0.40 (0.24-0.62)***† | 0.76 (0.45-1.24) | - | - |
| Conscientiousness |  | - | - | - | 0.83 (0.57-1.20) | - | - | - | - | - |
| Extraversion |  | - | - | - | 0.83 (0.58-1.17) | 0.89 (0.64-1.24) | - | - | - | - |

\* $p < .05$ , \*\* $p < .01$ , \*\*\* $p < .001$ ; Independent variables are from age-82 unless otherwise stated: <sup>a</sup> age is age in days at time of questionnaire (mean age 84); <sup>b</sup> living alone at time of questionnaire (mean age 84). †associations remain significant after multiple testing correction via false discovery rate (FDR) estimation. <sup>c</sup> models were binary logistic regression models. <sup>d</sup> models were ordinal logistic regression models.

Wilcoxon signed rank tests indicated significant changes between ‘before’ and ‘during’ lockdown ratings for physical (W=30, Z=0.358, p<.001) and mental health (W=22, Z=0.480, p<0.001) which indicated that fewer participants rated their physical and mental health to be excellent or very good. However, there were no significant correlations between derived change scores and previously measured characteristics.

## DISCUSSION

In a well-characterised sample of community-dwelling 84-year-olds from the LBC1936, we conducted a questionnaire examining the impact of Scottish COVID-19 lockdown guidance on the lives of older people. This is one of the largest studies – exclusively in adults aged over 80 years – of psychosocial factors, health and lifestyle in relation to COVID-19 to-date. This study offers an important snapshot of the impact on octogenarians following two months of stringent lockdown restrictions. By linking questionnaire responses during lockdown with characteristics measured at least two years earlier (age-82), it highlights possible risk and protective factors for health and behaviour during lockdown, and adds to what is known about effects of the COVID-19 lockdown on older people.

Reassuringly, our findings indicate that this group of older adults coped relatively well during lockdown. Respondents had little direct experience of the virus and mostly had good self-reported physical and mental health, but experienced some changes to their routines and activities. However, the lockdown experience was not universally positive. Some experienced modest declines in self-reported mental and physical health; one-third of respondents experienced COVID-19-related stress or nervousness, and 25% felt lonely during lockdown. Results of our regression analyses highlight individual differences that may be associated with increased risk of, or protection against, negative outcomes during the current and future waves of the pandemic.

Being lonely during lockdown was associated with living alone and greater age-82 anxiety symptoms. Evidence on mental health during lockdown is mixed; some studies suggest those over age 70 are less likely to feel stressed or anxious or report a negative effect on their mental health than younger age groups, and others report that odds of reporting high anxiety during COVID-19 was twice as likely in those over age 75 than those under 24 (7,31). Non-responders in the current study were more likely to be living alone and to have greater age-82 anxiety symptoms; therefore, our results might underestimate the proportion of older people experiencing loneliness and the magnitude of the associations between loneliness, living alone, and anxiety in the general population. Given the known negative consequences of loneliness for older adults (9–15), public health measures to counteract loneliness are likely to be increasingly important.

Our finding that almost half of respondents reported decreased physical activity builds upon previous findings of lower levels of vigorous physical activity in adults during lockdown (33). This may be particularly important in the context of previous studies showing associations between physical fitness and cognitive ability (49) and cognitive decline (17) in older age, and on the mediating effect of physical activity on the relationship between stress levels and mental health (50). Individuals who left home less frequently during lockdown had poorer self-rated health and less professional occupational class. This may reflect that those with previous health problems, and those who may face greater material disadvantages (such as having fewer financial resources due to past occupational status) when managing the stress of the virus (51), may take greater precautions to safeguard their health. This complements findings that the threat of COVID-19 is perceived to be lower for those who are healthier and have higher income (22). Healthcare providers considering web-based provision of information and interventions should consider that online campaigns may not reach all parts of the older adult population equally; men and those with lower age-82 gf were less likely to report increased internet usage during lockdown. A more positive outcome is that over 60% of respondents started or returned to a pastime during lockdown. A previous LBC1936 study found that playing analogue games was associated with less cognitive decline in those aged 70-79 (52), however cognitive benefits of different types of pastimes may vary across different age groups (49). Future studies might examine whether there were benefits associated with taking part in specific types of pastime (e.g active versus passive) during lockdown.

### Strengths and Limitations

This was one of the first studies to collect data on the impact of the COVID-19 lockdown on octogenarians. This adds to and strengthens the current COVID-19 literature, specifically in terms of examining outcomes in older adults. Few studies to date have included large samples of older adults; where older adults have been included, sample sizes tend to be low. Even among larger-scale studies, and those which sampled a wider range of older ages (e.g. extending from age 70 into late 80s), few report equivalent sample sizes to that achieved in the current study, with others ranging between only 22 participants over age 80 (23) to 237 when adults aged 60 years and below are included (19). Additionally, LBC1936 members have a narrow age range, which reduces the likelihood that results are confounded by variation in age. Due to the wealth of previously collected data, the current study also had the rare advantage of being able to link COVID-19 questionnaire outcomes with longitudinal characteristics, thereby avoiding problems inherent in retrospective data collection, such as results being affected by poor recall memory or current circumstances. Furthermore, multivariate models were able to include relevant variables to minimise confounding. The questionnaire was distributed at an expedient time, when lockdown guidance was consistent for all respondents, and respondents completed it around two months after the onset of lockdown, so responses were unlikely to be affected by a short-lived peak in anxiety or emotional distress which might have occurred when the pandemic first took hold.

This study has limitations. The LBC1936 is a self-selecting sample consisting of mostly white Scottish participants who are likely to be healthier than the general population (40,41), potentially limiting the generalisability of our results, which may be underestimates of the experiences in the general population. The questionnaire relied on self-report, without objective measures to gauge the accuracy of results. By administering the questionnaire online, willing participants without the means to respond to online formats may have been excluded.

## CONCLUSIONS

In this study, we reported on the impact of COVID-19 lockdown in Scotland on psychosocial factors, health, and lifestyle in members of LBC1936. Results indicated that those with lower cognitive functioning, less professional occupational social class, lower emotional stability, greater anxiety symptoms, and living alone may be particularly at risk of negative lockdown-related outcomes, including loneliness and reduced physical activity, poorer self-reported mental and physical health, and greater stress and nervousness. Older adults with these characteristics may benefit from additional support to reduce the risk of negative outcomes. Additionally, policy makers and healthcare providers might focus on outcomes of loneliness and physical activity, which are widely known to have attendant negative consequences.

## Supporting information

R code; Correlation matrix; STROBE

## Data Availability

The datasets generated and/or analysed during the current study are not publicly available due to them containing information that could compromise participant consent and confidentiality, but are available from the corresponding author on reasonable request.

## ABBREVIATIONS

COVID-19: Coronavirus disease
LBC1936: Lothian Birth Cohort 1936
SMS1947: Scottish Mental Survey 1947
BMI: Body Mass Index
FDR: false discovery rate

## DECLARATIONS

### Ethics approval and consent to participate

Participants gave full and informed written consent before partaking in this study. Ethical approval was obtained from Multi-Centre Research Ethics Committee for Scotland (MREC/01/0/56; Wave 1), the Lothian Research Ethics Committee (LREC/2003/2/29; Wave 1), and the Scotland A Research Ethics Committee (07/MRE00/58; Waves 2-5). The authors assert that all procedures contributing to this work comply with the ethical standards of the relevant national and institutional committees on human experimentation and with the Helsinki Declaration of 1975, as revised in 2008.

### Consent to publish

Not applicable.

### Authors’ contributions

AT and DP contributed equally to this study, conducting all analyses and authoring the paper. AT, DP, JAO and JC conducted cognitive testing appointments at previous waves. All authors contributed to the design of the COVID-19 questionnaire, contributed to the design of the study, and advised paper edits. All authors have agreed with the final version of this paper.

## Acknowledgements

We are grateful to all participants of the Lothian Birth Cohort 1936 study. We thank the LBC1936 research team, and the research nurses at the Wellcome Trust Clinical Research Facility, Western General Hospital, Edinburgh, for their contributions to previous waves of the study. We thank Professor Ian Deary for suggestions on study design and on earlier versions of this paper.

## REFERENCES

1. WHO. WHO Director-General’s opening remarks at the media briefing on COVID-19 [Internet]. 2020 [cited 2020 Jul 8]. Available from: https://www.who.int/dg/speeches/detail/who-director-general-s-opening-remarks-at-the-media-briefing-on-covid-1911-march-2020

2. Scottish Government. Prime Minister’s statement on coronavirus (COVID-19): 23 March 2020 [Internet]. 2020 [cited 2020 Jul 8]. Available from: 2. https://www.gov.uk/government/speeches/pm-address-to-the-nation-on-coronavirus-23-march-2020

3. Bonanad C, García-Blas S, Tarazona-Santabalbina F, Sanchis J, Bertomeu-González V, Fácila L, et al. The effect of age on mortality in patients with Covid-19: a metanalysis with 611,583 subjects. J Am Med Dir Assoc. 2020 May;S1525861020304412.

4. Docherty AB, Harrison EM, Green CA, Hardwick HE, Pius R, Norman L, et al. Features of 20 133 UK patients in hospital with covid-19 using the ISARIC WHO Clinical Characterisation Protocol: prospective observational cohort study. BMJ. 2020 May 22;m1985.

5. NRS. NRS Key Findings [Internet]. 2020 [cited 2020 Jun 12]. Available from: https://www.nrscotland.gov.uk/covid19stats

6. Yang J, Zheng Y, Gou X, Pu K, Chen Z, Guo Q, et al. Prevalence of comorbidities and its effects in patients infected with SARS-CoV-2: a systematic review and meta-analysis. Int J Infect Dis. 2020 May;94:91–5.

7. Office for National Statistics. Coronavirus and the social impacts on older people in Great Britain: 3 April to 10 May 2020 [Internet]. Office for National Statistics; 2020 Jun. Available from: https://www.ons.gov.uk/peoplepopulationandcommunity/birthsdeathsandmarriages/ageing/articles/coronavirusandthesocialimpactsonolderpeopleingreatbritain/3aprilto10may2020

8. Holmes EA, O’Connor RC, Perry VH, Tracey I, Wessely S, Arseneault L, et al. Multidisciplinary research priorities for the COVID-19 pandemic: a call for action for mental health science. Lancet Psychiatry. 2020 Jun;7(6):547–60.

9. Gow AJ, Corley J, Starr JM, Deary IJ. Which Social Network or Support Factors are Associated with Cognitive Abilities in Old Age? Gerontology. 2013;59(5):454–63.

10. Holt-Lunstad J, Smith TB, Baker M, Harris T, Stephenson D. Loneliness and Social Isolation as Risk Factors for Mortality: A Meta-Analytic Review. Perspect Psychol Sci. 2015 Mar;10(2):227–37.

11. Novotney A. The risks of social isolation. Am Psychol Assoc [Internet]. 2019;50(5). Available from: https://www.apa.org/monitor/2019/05/ce-corner-isolation

12. Shankar A, Hamer M, McMunn A, Steptoe A. Social Isolation and Loneliness: Relationships With Cognitive Function During 4 Years of Follow-up in the English Longitudinal Study of Ageing. Psychosom Med. 2013;75(2):161–70.

13. Elovainio M, Hakulinen C, Pulkki-Råback L, Virtanen M, Josefsson K, Jokela M, et al. Contribution of risk factors to excess mortality in isolated and lonely individuals: an analysis of data from the UK Biobank cohort study. Lancet Public Health. 2017 Jun;2(6):e260–6.

14. Alcaraz KI, Eddens KS, Blase JL, Diver WR, Patel AV, Teras LR, et al. Social Isolation and Mortality in US Black and White Men and Women. Am J Epidemiol. 2019 Jan 1;188(1):102–9.

15. Hawkley LC, Capitanio JP. Perceived social isolation, evolutionary fitness and health outcomes: a lifespan approach. Philos Trans R Soc B Biol Sci. 2015 May 26;370(1669):20140114.

16. Lippi G, Henry BM, Sanchis-Gomar F. Physical inactivity and cardiovascular disease at the time of coronavirus disease 2019 (COVID-19). Eur J Prev Cardiol. 2020 Jun;27(9):906–8.

17. Okely JA, Deary IJ. Associations Between Declining Physical and Cognitive Functions in the Lothian Birth Cohort 1936. Newman A, editor. J Gerontol Ser A. 2020 Jun 18;75(7):1393–402.

18. Webb L. COVID-19 lockdown: A perfect storm for older people’s mental health. J Psychiatr Ment Health Nurs. 2020 Jun 28;jpm.12644.

19. González-Sanguino C, Ausín B, Castellanos MÁ, Saiz J, López-Gómez A, Ugidos C, et al. Mental health consequences during the initial stage of the 2020 Coronavirus pandemic (COVID-19) in Spain. Brain Behav Immun. 2020 May;S0889159120308126.

20. McBride O, Murphy J, Shevlin M, Gibson Miller J, Hartman TK, Hyland P, et al. Monitoring the psychological impact of the COVID-19 pandemic in the general population: an overview of the context, design and conduct of the COVID-19 Psychological Research Consortium (C19PRC) Study [Internet]. PsyArXiv; 2020 Apr [cited 2020 Jul 8]. Available from: https://osf.io/wxe2n

21. Nelson LM, Simard JF, Oluyomi A, Nava V, Rosas LG, Bondy M, et al. US Public Concerns About the COVID-19 Pandemic From Results of a Survey Given via Social Media. JAMA Intern Med [Internet]. 2020 Apr 7 [cited 2020 Jun 5]; Available from: https://jamanetwork.com/journals/jamainternalmedicine/fullarticle/2764368

22. Wolf MS, Serper M, Opsasnick L, O’Conor RM, Curtis LM, Benavente JY, et al. Awareness, Attitudes, and Actions Related to COVID-19 Among Adults With Chronic Conditions at the Onset of the U.S. Outbreak: A Cross-sectional Survey. Ann Intern Med. 2020 Apr 9;M20–1239.

23. Cowan K. The Academy of Medical Sciences: Survey results: Understanding people’s concerns about the mental health impacts of the COVID-19 pandemic [Internet]. 2020 [cited 2020 Jun 21]. Available from: http://www.acmedsci.ac.uk/COVIDmentalhealthsurveys

24. Smith L, Jacob L, Yakkundi A, McDermott D, Armstrong NC, Barnett Y, et al. Correlates of symptoms of anxiety and depression and mental wellbeing associated with COVID-19: a cross-sectional study of UK-based respondents. Psychiatry Res. 2020 May;113138.

25. Huang Y, Zhao N. Generalized anxiety disorder, depressive symptoms and sleep quality during COVID-19 outbreak in China: a web-based cross-sectional survey. Psychiatry Res. 2020 Jun;288:112954.

26. Qiu J, Shen B, Zhao M, Wang Z, Xie B, Xu Y. A nationwide survey of psychological distress among Chinese people in the COVID-19 epidemic: implications and policy recommendations. Gen Psychiatry. 2020 Mar;33(2):e100213.

27. Brooks SK, Webster RK, Smith LE, Woodland L, Wessely S, Greenberg N, et al. The psychological impact of quarantine and how to reduce it: rapid review of the evidence. The Lancet. 2020 Mar;395(10227):912–20.

28. Vindegaard N, Eriksen Benros M. COVID-19 pandemic and mental health consequences: Systematic review of the current evidence. Brain Behav Immun. 2020 May;S0889159120309545.

29. Wang C, Pan R, Wan X, Tan Y, Xu L, Ho CS, et al. Immediate Psychological Responses and Associated Factors during the Initial Stage of the 2019 Coronavirus Disease (COVID-19) Epidemic among the General Population in China. Int J Environ Res Public Health. 2020 Mar 6;17(5):1729.

30. Lau ALD, Chi I, Cummins RA, Lee TMC, Chou K-L, Chung LWM. The SARS (Severe Acute Respiratory Syndrome) pandemic in Hong Kong: Effects on the subjective wellbeing of elderly and younger people. Aging Ment Health. 2008 Nov;12(6):746–60.

31. Office for National Statistics. Coronavirus and anxiety, Great Britain: 3 April 2020 to 10 May 2020 [Internet]. Office for National Statistics; 2020 Jun. Available from: https://www.ons.gov.uk/peoplepopulationandcommunity/wellbeing/articles/coronavirusandanxietygreatbritain/3april2020to10may2020

32. Cellini N, Canale N, Mioni G, Costa S. Changes in sleep pattern, sense of time and digital media use during COVID- 19 lockdown in Italy. J Sleep Res [Internet]. 2020 May 15 [cited 2020 Jul 8]; Available from: https://onlinelibrary.wiley.com/doi/abs/10.1111/jsr.13074

33. Cheval B, Sivaramakrishnan H, Maltagliati S, Fessler L, Forestier C, Sarrazin P, et al. Relationships Between Changes in Self-Reported Physical Activity and Sedentary Behaviours and Health During the Coronavirus (COVID-19) Pandemic in France and Switzerland [Internet]. SportRxiv; 2020 Apr [cited 2020 Jul 8]. Available from: https://osf.io/ydv84

34. Bacon AM, Corr PJ. Coronavirus (COVID- 19) in the United Kingdom: A personality-based perspective on concerns and intention to self-isolate. Br J Health Psychol. 2020 Apr 29;bjhp.12423.

35. Wright L, Steptoe A, Fancourt D. Are we all in this together? Longitudinal assessment of cumulative adversities by socioeconomic position in the first 3 weeks of lockdown in the UK. J Epidemiol Community Health. 2020 Jun 5;jech-2020-214475.

36. Gibson Miller J, Hartman TK, Levita L, Martinez AP, Mason L, McBride O, et al. Capability, opportunity, and motivation to enact hygienic practices in the early stages of the COVID- 19 outbreak in the United Kingdom. Br J Health Psychol. 2020 May 16;bjhp.12426.

37. Rimfeld K, Malancini M, Allegrini A, Packer AE, McMillan A, Ogden R, et al. Genetic correlates of psychological responses to the COVID-19 crisis in young adult twins in Great Britain. 2020 May 27 [cited 2020 Jun 5]; Available from: https://www.researchsquare.com/article/rs-31853/v1

38. Scottish Council for Research in Education. The trend of Scottish intelligence: A comparison of the 1947 and 1932 surveys of the intelligence of eleven-year-old pupils. London: University of London Press.; 1949.

39. Deary IJ, Gow AJ, Taylor MD, Corley J, Brett C, Wilson V, et al. The Lothian Birth Cohort 1936: a study to examine influences on cognitive ageing from age 11 to age 70 and beyond. BMC Geriatr. 2007 Dec;7(1):28.

40. Deary IJ, Gow AJ, Pattie A, Starr JM. Cohort profile: the Lothian Birth Cohorts of 1921 and 1936. Int J Epidemiol. 2012;41(6):1576–84.

41. Taylor AM, Pattie A, Deary IJ. Cohort Profile Update: The Lothian Birth Cohorts of 1921 and 1936. Int J Epidemiol. 2018 Aug 1;47(4):1042–1042r.

42. Generation Scotland. CovidLife Survey First Report [Internet]. 2020 [cited 2020 Jul 9]. Available from: https://www.ed.ac.uk/files/atoms/files/2020-05-15_covidlifesurvey_report_final_web.pdf

43. Folstein MF, Folstein SE, McHugh PR. “Mini-mental state”. J Psychiatr Res. 1975 Nov;12(3):189–98.

44. Sainsbury S. Measuring disability. G. Bell; 1973.

45. R Core Team. R: A Language and Environment for Statistical Computing [Internet]. Vienna, Austria: R Foundation for Statistical Computing; 2019. Available from: https://www.R-project.org/

46. IBM Corp. IBM SPSS Statistics for Windows, Version 25.0. Armonk, NY: IBM Corp; 2017.

47. Holm S. A simple sequentially rejective multiple test procedure. Scand J Stat. 1979;65–70.

48. Benjamini Y, Hochberg Y. Controlling the False Discovery Rate: A Practical and Powerful Approach to Multiple Testing. J R Stat Soc Ser B Methodol. 1995 Jan;57(1):289–300.

49. Gow AJ, Corley J, Starr JM, Deary IJ. Reverse causation in activity-cognitive ability associations: The Lothian Birth Cohort 1936. Psychol Aging. 2012;27(1):250–5.

50. Kwag KH, Martin P, Russell D, Franke W, Kohut M. The impact of perceived stress, social support, and home-based physical activity on mental health among older adults. Int J Aging Hum Dev. 2011;72(2):137–54.

51. Hamilton R. Scarcity and Coronavirus. J Public Policy Mark. 2020 May 28;4391562092811.

52. Altschul DM, Deary IJ. Playing Analog Games Is Associated With Reduced Declines in Cognitive Function: A 68-Year Longitudinal Cohort Study. J Gerontol Ser B. 2020;75(3):474–82.

53. General Register Office. Analysis of General Practice Records for April 1952 - March 1954. London:HMSO; 1956.

54. Office of Population Censuses and Surveys. Classification of Occupations 1980. London: Her Majesty’s Stationery Office; 1980.

55. Wechsler D. WAIS-IV Administration and Scoring Manual. San Antonio, TX: The Psychological Corporation.; 2008.

56. Luciano M, Gow AJ, Harris SE, Hayward C, Allerhand M, Starr JM, et al. Cognitive ability at age 11 and 70 years, information processing speed, and APOE variation: The Lothian Birth Cohort 1936 study. Psychol Aging. 2009 Mar;24(1):129–38.

57. Fawns-Ritchie C, Starr JM, Deary IJ. Health literacy, cognitive ability and smoking: a cross-sectional analysis of the English Longitudinal Study of Ageing. BMJ Open. 2018 Oct;8(10):e023929.

58. Zigmond AS, Snaith RP. The hospital anxiety and depression scale. Acta Psychiatr Scand. 1983;67(6):361–70.

59. Goldberg LR. An alternative ‘description of personality’: The Big-Five factor structure. J Pers Soc Psychol. 1990;59(6):1216–29.

