## Supplementary figure 1 for "Impact of COVID-19 lockdown on psychosocial factors, health, and lifestyle in Scottish octogenarians: the Lothian Birth Cohort 1936 Study"

### SUPPLEMENTARY MATERIALS: Supplementary figure 1

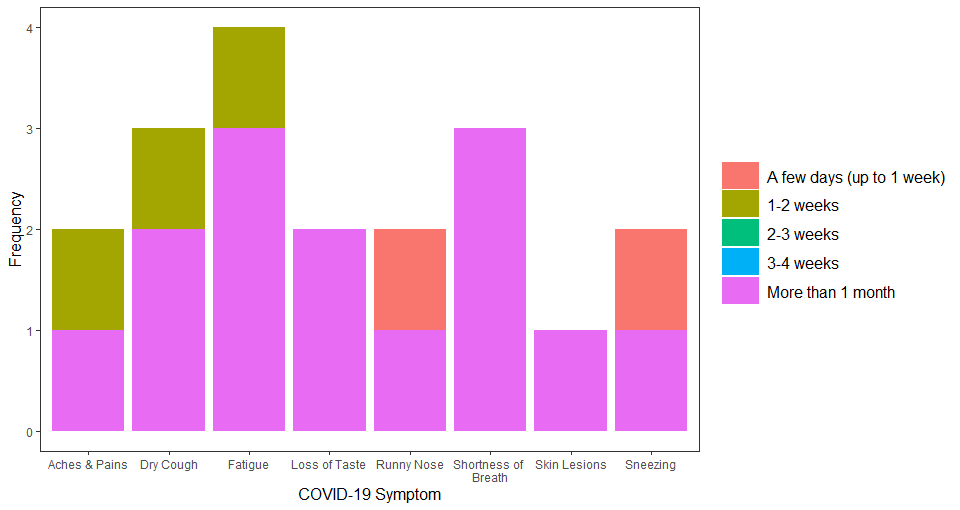

Supplementary figure 1. LBC1936 participants’ (n=4) responses to duration of COVID-19 symptoms
