## Supplementary figure 2 for "Impact of COVID-19 lockdown on psychosocial factors, health, and lifestyle in Scottish octogenarians: the Lothian Birth Cohort 1936 Study"

### SUPPLEMENTARY MATERIALS: Supplementary figure 2

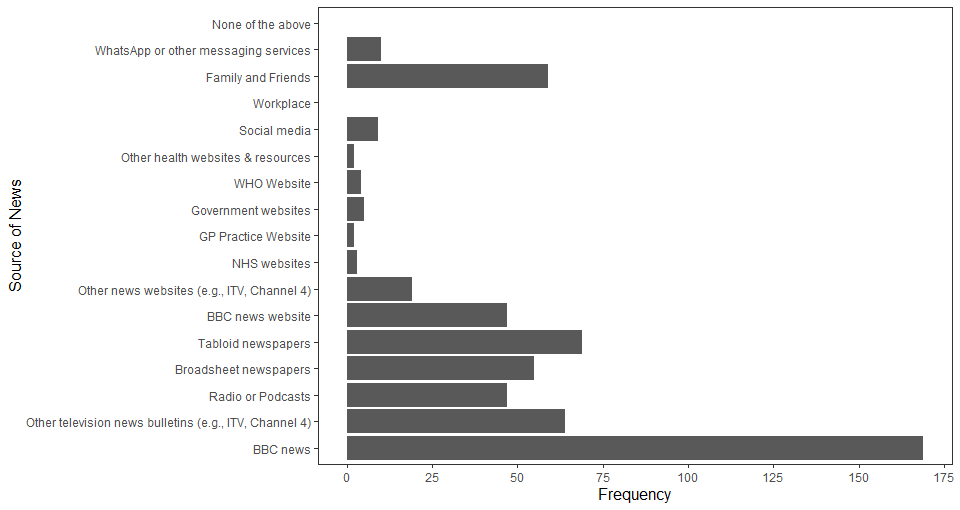

Supplementary figure 2. Types of news source used by LBC1936 participants to stay informed about COVID-19
