## Supplementary figure 3 for "Impact of COVID-19 lockdown on psychosocial factors, health, and lifestyle in Scottish octogenarians: the Lothian Birth Cohort 1936 Study"

### SUPPLEMENTARY MATERIALS: Supplementary figure 3

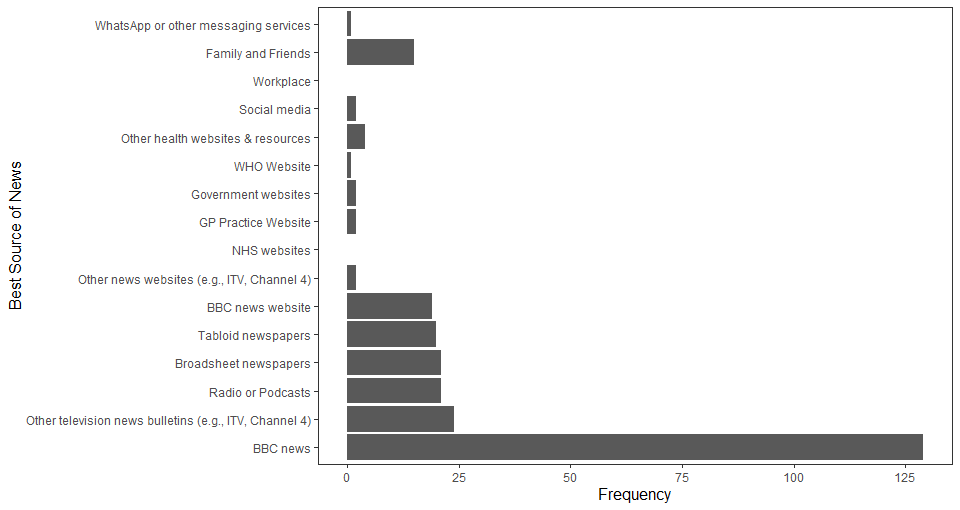

Supplementary figure 3. News sources rated most helpful by LBC1936 participants staying informed about COVID-19
