## Supplementary figure 5 for "Impact of COVID-19 lockdown on psychosocial factors, health, and lifestyle in Scottish octogenarians: the Lothian Birth Cohort 1936 Study"

### SUPPLEMENTARY MATERIALS: Supplementary figure 5

LBC1936 participants’ self-reported methods of contact with family and friends before and during COVID-19 lockdown

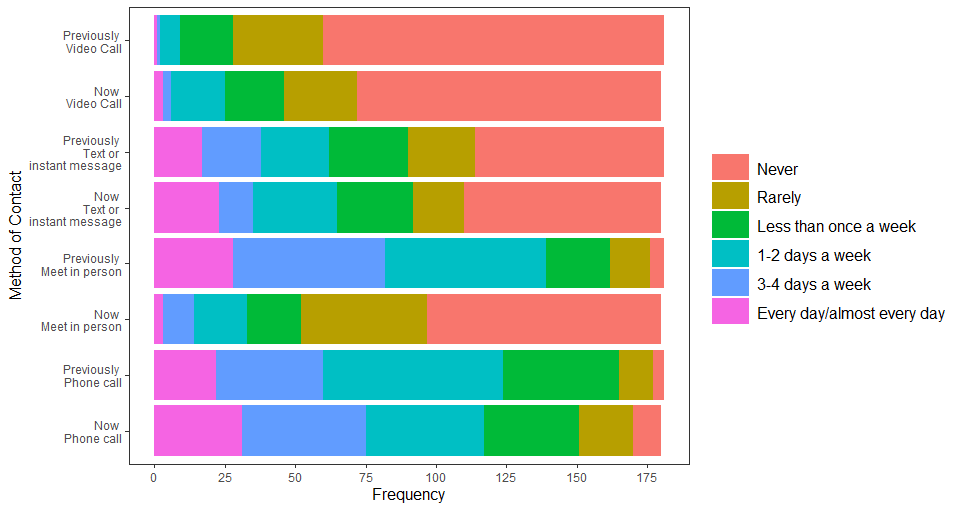

Supplementary figure 5. LBC1936 participants’ self-reported methods of contact with friends before and during lockdown
