## Supplementary table 1 for "Impact of COVID-19 lockdown on psychosocial factors, health, and lifestyle in Scottish octogenarians: the Lothian Birth Cohort 1936 Study"

### SUPPLEMENTARY MATERIALS: Supplementary table 1

LBC1936 participant responses to COVID-19 questionnaire

Supplementary Table 1. LBC1936 participant responses to COVID-19 questionnaire: Experience of COVID-19

| Experience of COVID-19 questions | N | % |
| --- | --- | --- |
| Do you think that you have had, or currently have COVID-19? |  |  |
| No | 186 | 97.89 |
| Yes, suspected but not tested (self-diagnosis based on symptoms) | 4 | 2.11 |
| Yes, suspected but not tested (diagnosed by medical professional based on symptoms) | 0 | - |
| Yes, confirmed by a positive test | 0 | - |
| Which symptoms did you have? (answered only by those who have had COVID-19) |  |  |
| Fatigue | 4 | 100 |
| Dry cough | 3 | 75.0 |
| Shortness of breath | 3 | 75.0 |
| Aches and pains | 2 | 50.0 |
| Runny nose | 2 | 50.0 |
| Sneezing | 2 | 50.0 |
| Loss of taste | 2 | 50.0 |
| Chilblain-like skin lesions | 1 | 25.0 |
| Fever | 0 | - |
| Headaches | 0 | - |
| Sore throat | 0 | - |
| Diarrhoea | 0 | - |
| Loss of smell | 0 | - |
| Other symptoms | 0 | - |
| During the time that you had or suspected you had COVID-19 did you: |  |  |
| Self-isolate of distance from others in the household | 2 | 50.0 |
| Call NHS 111 or GP | 1 | 25.0 |
| Visit a COVID hub to see a doctor | 1 | 25.0 |
| Do nothing, because already shielding | 0 | - |
| Get swabbed for COVID-19 | 0 | - |
| Get admitted to hospital | 0 | - |
| Get admitted to an intensive care unit | 0 | - |
| Need a ventilator to help you breath | 0 | - |
| Do you think anyone else in your household has had or currently has COVID-19? |  |  |
| Yes | 0 |  |
| No | 190 | 100 |
| Have you been contacted by letter or text message to say you are at severe risk from COVID-19 due to an underlying health condition and should be shielding? |  |  |
| Yes | 26 | 13.7 |
| No | 164 | 86.3 |
| Has anxiety about COVID-19 caused you to avoid or postpone contacting or attending a medical service? |  |  |
| No, I have not been anxious about contacting or attending a medical service | 103 | 74.2 |
| Yes, postponed contacting a medical service (such as calling my GP) | 13 | 6.8 |
| Yes, postponed attending a medical appointment that was made previously | 11 | 5.8 |
| I was anxious about COVID-19 but did not avoid or postpone contacting or attending a medical service | 12 | 6.3 |
| Not applicable | 51 | 26.8 |
