## Supplementary table 2 for "Impact of COVID-19 lockdown on psychosocial factors, health, and lifestyle in Scottish octogenarians: the Lothian Birth Cohort 1936 Study"

### SUPPLEMENTARY MATERIALS: Supplementary table 2

LBC1936 participant responses to COVID-19 questionnaire

Supplementary Table 2. LBC1936 participant responses to COVID-19 questionnaire: COVID-19 knowledge and guidance

| COVID-19 knowledge and guidance questions | N | % |
| --- | --- | --- |
| How would you rate your knowledge of COVID-19 (e.g. the symptoms of the disease, how it is transmitted, and how to minimise your exposure to it)? |  |  |
| Extremely good | 85 | 44.7 |
| Somewhat good | 95 | 50.0 |
| Neither good nor bad | 8 | 4.2 |
| Somewhat bad | 2 | 1.1 |
| Extremely bad | 0 | - |
| Do you find the Scottish Government guidance on COVID-19 easy to understand? |  |  |
| Extremely easy | 73 | 38.4 |
| Somewhat easy | 91 | 47.9 |
| Neither easy nor difficult | 18 | 9.5 |
| Somewhat difficult | 7 | 3.7 |
| Extremely difficult | 0 |  |
| Not aware of government guidance | 1 | 0.5 |
| Have you been following the government guidance on social distancing? |  |  |
| Always | 138 | 72.6 |
| Most of the time | 50 | 26.3 |
| Some of the time | 2 | 1.1 |
| Never | 0 | - |
| Have you been following the government guidance on staying at home as much as possible? |  |  |
| Always | 127 | 66.8 |
| Most of the time | 57 | 30.0 |
| Some of the time | 6 | 3.2 |
| Never | 0 | - |
| Have you been following the government guidance on hand washing? |  |  |
| Always | 141 | 75.0 |
| Most of the time | 43 | 22.9 |
| Some of the time | 4 | 2.1 |
| Never | 0 | - |
| Have you been following the government guidance on self-isolating (if suffering COVID-19 symptoms)? |  |  |
| Always | 35 | 79.5 |
| Most of the time | 4 | 9.1 |
| Some of the time | 0 | - |
| Never | 5 | 11.4 |
| Not applicable |  |  |
| How often have you been leaving your home since COVID-19 measures were introduced (23rd March 2020)? |  |  |
| Multiple times a day | 4 | 2.1 |
| Once per day | 80 | 42.1 |
| A few times per week | 49 | 25.8 |
| Once per week | 9 | 4.7 |
| Less than once per week (e.g. once per fortnight) | 21 | 11.1 |
| Never, since COVID-19 measures were introduced | 27 | 14.21 |
| When leaving your home for a daily walk or other exercise, how likely are you to accidentally come into close contact with someone not living in your household (e.g. less than 2 metres)? |  |  |
| I don’t leave my home | 28 | 14.7 |
| Not at all likely | 66 | 34.7 |
| Not that likely | 68 | 35.8 |
| Somewhat likely | 26 | 13.7 |
| Very likely | 2 | 1.1 |
| How often do you keep up to date on COVID-19 related news? |  |  |
| Multiple times per day | 53 | 27.9 |
| Daily | 122 | 64.2 |
| Every few days | 13 | 6.8 |
| Weekly | 0 | - |
| Less than weekly | 2 | 1.1 |
