## Supplementary table 3 for "Impact of COVID-19 lockdown on psychosocial factors, health, and lifestyle in Scottish octogenarians: the Lothian Birth Cohort 1936 Study"

### SUPPLEMENTARY MATERIALS: Supplementary table 3

LBC1936 participant responses to COVID-19 questionnaire

Supplementary Table 3. LBC1936 participant responses to COVID-19 questionnaire: Living situation and impact on day-to-day living.

| Living situation and impact on day-to-day living questions | N | % |
| --- | --- | --- |
| Including yourself, how many people currently live in your household? (Mean/SD) |  |  |
| 1 | 74 | 39.8 |
| 2 | 99 | 53.2 |
| 3 | 8 | 4.3 |
| 4 | 2 | 1.1 |
| 5 | 3 | 1.6 |
| Who currently lives in your household with you? (select all that apply) |  |  |
| Live alone | 73 | 38.4 |
| Spouse/partner | 107 | 56.3 |
| Child/children | 9 | 4.7 |
| Grandchildren/Great grandchildren | 2 | 1.05 |
| Parent(s) or parent(s)-in-law | 0 | - |
| Other family member(s) | 12 | 6.3 |
| Paid carers | 0 | - |
| Friend(s) or other non-family member(s) | 1 | 0.5 |
| What kind of area do you live in? |  |  |
| Rural | 21 | 11.1 |
| Urban | 55 | 28.9 |
| Suburban | 114 | 60.0 |
| Do you have a garden (including access to a shared garden) or allotment? |  |  |
| Garden | 172 | 91.0 |
| Allotment | 0 | - |
| No | 17 | 9.0 |
| How much has COVID-19 changed your daily routine? |  |  |
| A lot | 65 | 34.6 |
| Somewhat | 70 | 37.2 |
| A little | 40 | 21.3 |
| Not at all | 13 | 6.9 |
| Have you received any additional help in your daily life with things such as grocery shopping, errands, or picking up medications since COVID-19 measures were introduced? |  |  |
| Yes | 119 | 62.6 |
| No | 71 | 37.4 |
| If yes, since COVID-19 measures were introduced have you newly received additional help in daily life with: |  |  |
| Grocery shopping | 112 | 59.0 |
| Picking up medicines | 46 | 24.2 |
| Other | 16 | 8.4 |
| No new additional help received | 71 | 37.4 |
| What actions have you taken to continue to get any prescription medicines since COVID-19 measures were introduced (23rd March 2020)? |  |  |
| Switched order to mail order or delivery | 58 | 30.5 |
| Increased the prescription to a longer supply (e.g. 90-day supply) | 12 | 6.3 |
| Other | 52 | 27.4 |
| None | 58 | 38.5 |
| Not applicable | 10 | 5.3 |
| Are there any local initiatives in your neighbourhood or broader community to help those who are self-isolating e.g. getting shopping, medications etc.? |  |  |
| Yes | 98 | 51.6 |
| No | 12 | 6.36 |
| Don’t know | 80 | 42.1 |
| Compared to before COVID-19 measures were introduced, have you found the need to use more non-cash alternatives such as paying for groceries by credit/debit card? |  |  |
| Yes | 123 | 64.7 |
| No | 67 | 35.3 |
| How important is it for you to be able to use cash? |  |  |
| Very important | 26 | 13.7 |
| Quite important | 41 | 21.6 |
| Not very important | 84 | 44.2 |
| Not important at all | 39 | 20.5 |
| How has your internet usage changed since COVID-19 measures were introduced (23rd March 2020)? |  |  |
| I use the internet much more now | 33 | 18.5 |
| I use the internet slightly more now | 64 | 36.0 |
| My internet usage is about the same | 78 | 43.8 |
| I use the internet slightly less now | 1 | 0.6 |
| I use the internet much less now | 2 | 1.1 |
| If you are using the internet more, do you think you will continue to use the internet more often after the COVID-19 emergency has passed? |  |  |
| Yes | 36 | 37.1 |
| No | 28 | 28.9 |
| Don’t know | 33 | 34.0 |
