## Supplementary table 4 for "Impact of COVID-19 lockdown on psychosocial factors, health, and lifestyle in Scottish octogenarians: the Lothian Birth Cohort 1936 Study"

### SUPPLEMENTARY MATERIALS: Supplementary table 4

### LBC1936 participant responses to COVID-19 questionnaire

Supplementary Table 4. LBC1936 participant responses to COVID-19 questionnaire: Social connectedness.

| Social Connectedness questions | N | % |
| --- | --- | --- |
| Since the COVID-19 measures were introduced (23rd March 2020), has contact with your neighbours changed? |  |  |
| More contact now | 64 | 33.7 |
| The same | 89 | 46.8 |
| Less contact now | 37 | 19.4 |
| If contact with your neighbours has changed, how have you found this experience? |  |  |
| Positive | 63 | 62.4 |
| Neutral | 32 | 31.7 |
| Negative | 6 | 5.9 |
