## Supplementary table 5 for "Impact of COVID-19 lockdown on psychosocial factors, health, and lifestyle in Scottish octogenarians: the Lothian Birth Cohort 1936 Study"

### SUPPLEMENTARY MATERIALS: Supplementary table 5

LBC1936 participant responses to COVID-19 questionnaire

Supplementary Table 5. LBC1936 participant responses to COVID-19 questionnaire: Self-reported physical and mental health and loneliness.

| Self-reported physical and mental health and loneliness questions | N | % |
| --- | --- | --- |
| How often have you felt lonely during the past week? |  |  |
| None or almost none of the time | 144 | 76.2 |
| Some of the time | 39 | 20.6 |
| Most of the time | 5 | 2.7 |
| All or almost all of the time | 1 | 0.5 |
| In general, before the COVID-19 measures were introduced (23rd March 2020), would you say your emotional and mental health was: |  |  |
| Excellent | 62 | 33.0 |
| Very good | 98 | 52.1 |
| Good | 22 | 11.7 |
| Fair | 6 | 3.2 |
| Poor | 0 | - |
| In general, since the COVID-19 measures were introduced, would you say your emotional and mental health is: |  |  |
| Excellent | 45 | 23.8 |
| Very good | 82 | 43.4 |
| Good | 47 | 24.9 |
| Fair | 11 | 5.8 |
| Poor | 4 | 2.1 |
| In general, before the COVID-19 measures were introduced (23rd March 2020), would you say your physical health was: |  |  |
| Excellent | 16 | 8.8 |
| Very good | 85 | 47.0 |
| Good | 60 | 33.2 |
| Fair | 20 | 11.1 |
| Poor | 0 | - |
| In general, since the COVID-19 measures were introduced, would you say your physical health is: |  |  |
| Excellent | 9 | 5.0 |
| Very good | 77 | 42.8 |
| Good | 67 | 37.2 |
| Fair | 27 | 15.0 |
| Poor | 0 | - |
| In the last two weeks, how often have you felt nervous or stressed because of COVID-19? |  |  |
| Never | 120 | 63.5 |
| Some of the time | 66 | 34.9 |
| Most of the time | 3 | 1.6 |
| All of the time | 0 | - |
