## Supplementary table 6 for "Impact of COVID-19 lockdown on psychosocial factors, health, and lifestyle in Scottish octogenarians: the Lothian Birth Cohort 1936 Study"

### SUPPLEMENTARY MATERIALS: Supplementary table 6

LBC1936 participant responses to COVID-19 questionnaire

Supplementary table 6. LBC1936 participant responses to COVID-19 questionnaire: Behaviour

| Behaviour questions | N | % |
| --- | --- | --- |
| Do you ever drink alcohol? |  |  |
| Yes | 121 | 64.0 |
| No | 28 | 14.8 |
| Special occasions only (less than once per month) | 40 | 21.2 |
| If yes, compared to before COVID-19 measures were introduced are you: |  |  |
| Drinking more alcohol now | 14 | 11.7 |
| Drinking about the same amount of alcohol now | 77 | 64.2 |
| Drinking less alcohol now | 29 | 24.2 |
| What is your current smoking status (cigarettes only)? |  |  |
| Smoker | 2 | 1.2 |
| Ex-smoker | 75 | 39.7 |
| Never smoked | 112 | 59.3 |
| If you are a current smoker, compared to before COVID-19 measures were introduced are you: |  |  |
| Smoking more now | 1 | 50.0 |
| Smoking about the same now | 1 | 50.0 |
| Smoking less now | 0 | - |
| Compared to before COVID-19 measures were introduced (23rd March 2020), is your diet: |  |  |
| Much healthier now | 7 | 3.7 |
| Slightly healthier now | 28 | 14.8 |
| About the same now | 139 | 73.5 |
| Slightly less healthy now | 14 | 7.4 |
| Much less healthy now | 1 | 0.5 |
| How much are you eating compared to before the COVID-19 measures were introduced? |  |  |
| More now | 19 | 10.1 |
| About the same | 146 | 77.3 |
| Less now | 24 | 12.7 |
| Compared to before COVID-19 measures were introduced (23rd March 2020), how much physical activity are you doing now? This includes activities that make you breathe harder than normal (e.g., brisk walking). |  |  |
| Much more physical activity now | 7 | 3.7 |
| Slightly more physical activity now | 26 | 13.8 |
| About the same physical activity now | 65 | 34.4 |
| Slightly less physical activity now | 50 | 26.5 |
| Much less physical activity now | 41 | 21.7 |
| Since COVID-19 measures have been in place (23rd March 2020), have you returned to or started up a new pastime that you can do from home? (Select all that apply) |  |  |
| Work out or exercise | 32 | - |
| Relaxation or meditation | 4 | - |
| Cooking class / learning to cook | 5 | - |
| Language class / learning a language | 5 | - |
| Arts and crafts | 21 | - |
| Singing | 0 | - |
| Playing music | 7 | - |
| Listening to music | 33 | - |
| Reading | 67 | - |
| Board or card games | 13 | - |
| Online gaming | 3 | - |
| Writing for pleasure | 9 | - |
| Watching television/films | 63 | - |
| Listening to the radio/audiobooks | 18 | - |
| Baking | 24 | - |
| Gardening | 54 | - |
| Dancing | 1 | - |
| Online educational course | 2 | - |
| Other activity not listed above | 13 | - |
| No change in activities | 71 | - |
