## Supplementary table 7 for "Impact of COVID-19 lockdown on psychosocial factors, health, and lifestyle in Scottish octogenarians: the Lothian Birth Cohort 1936 Study"

### SUPPLEMENTARY MATERIALS: Supplementary table 7

Supplementary table 7. Factor loadings and percentage of variance explained by first unrotated component from principal components analysis of general fluid ability and general health literacy items

| Item | Unrotated factor loadings and percentage of variance explained |
| --- | --- |
| General fluid ability (gf) at age 82 |  |
| Matrix Reasoning | .71 |
| Block Design | .72 |
| Digit Symbol Coding | .79 |
| Digit Span Backwards | .62 |
| Letter-number Sequencing | .73 |
| Symbol Search | .82 |
| % of variance | 53.8% |
| General health literacy at age 73 |  |
| Rapid Estimate of Adult Literacy in Medicine (REALM) | .74 |
| Shortened Test of Functional Health Literacy in Adults (S-TOFHLA) | .80 |
| Newest Vital Sign (NVS) | .77 |
| % of variance | 59.7% |
