## Supplementary table 8 for "Impact of COVID-19 lockdown on psychosocial factors, health, and lifestyle in Scottish octogenarians: the Lothian Birth Cohort 1936 Study"

### SUPPLEMENTARY MATERIALS: Supplementary table 8

Supplementary table 8. Comparison of background characteristics for Lothian Birth Cohort 1936 participants who responded to the COVID-19 questionnaire versus those who did not respond.

| Background characteristic | Respondent | | Non-responder | | Difference test | | |
| --- | --- | --- | --- | --- | --- | --- | --- |
|  | Mean/N | SD/% | Mean/N | SD/% | t/χ^2^ | p | Cohen’s *d* |
| Age (years)* | 81.98 | 0.46 | 82.02 | 0.48 | 0.96 | 0.34 | 0.09 |
| Sex (*n* male) | 96 | 52.7 | 113 | 45.4 | 2.00 | 0.16 | - |
| Childhood occupational class | 2.83 | 0.94 | 2.88 | 0.96 | 0.46 | 0.65 | 0.05 |
| Adulthood occupational class | 2.00 | 0.83 | 2.40 | 0.92 | 4.70 | <.001 | 0.46 |
| Years of formal full-time education | 11.21 | 1.17 | 10.7 | 1.12 | -4.83 | <.001 | 0.47 |
| Marital status* |  |  |  |  | 3.41 | 0.06 | - |
| Married | 110 | 60.4 | 127 | 51.0 |  |  |  |
| Not married | 72 | 39.6 | 122 | 49.0 |  |  |  |
| Living alone* (*n* yes) | 60 | 33.0 | 114 | 45.8 | 6.65 | 0.01 | - |
| Moray House test (MHT) score at mean age 11† | 53.33 | 10.63 | 48.65 | 12.30 | -5.99 | <.001 | 0.48 |
| Mini mental state examination (MMSE) score* | 28.77 | 1.82 | 27.85 | 2.55 | -4.40 | <.001 | 0.42 |
| General cognitive ability* | 0.37 | 0.92 | -0.28 | 0.97 | -6.78 | <.001 | 0.69 |
| General health literacy score at mean age 73 | 0.45 | 0.82 | -0.08 | 0.97 | -5.71 | <.001 | 0.59 |
| Body Mass Index (BMI)* | 26.95 | 3.92 | 27.29 | 4.43 | 0.84 | 0.40 | 0.08 |
| Grip strength (kg; max in dominant hand)* | 27.82 | 8.77 | 25.77 | 8.62 | -2.37 | 0.02 | 0.24 |
| Forced expiratory volume in 1s (FEV1)* | 2.12 | 0.64 | 1.97 | 0.61 | -2.51 | 0.01 | 0.25 |
| History of hypertension history (*n* yes)* | 110 | 60.4 | 138 | 56.6 | 0.50 | 0.48 | - |
| History of cardiovascular disease ( *n* yes)* | 75 | 41.4 | 95 | 38.3 | 0.31 | 0.58 | - |
| History of diabetes( *n* yes)* | 19 | 10.4 | 32 | 12.9 | 0.38 | 0.54 | - |
| Glycated haemoglobin (HbA1c)* | 40.02 | 7.44 | 40.58 | 8.29 | 0.71 | 0.48 | 0.07 |
| Townsend disability scale score* | 1.35 | 2.20 | 2.36 | 3.45 | 3.67 | <.001 | 0.35 |
| Self-reported health* |  |  |  |  | 13.61 | 0.008 |  |
| Excellent | 23 | 12.6 | 19 | 7.6 |  |  |  |
| Very good | 86 | 47.3 | 91 | 36.5 |  |  |  |
| Good | 61 | 33.5 | 102 | 41.0 |  |  |  |
| Fair | 10 | 5.5 | 29 | 11.6 |  |  |  |
| Poor | 2 | 1.1 | 8 | 3.2 |  |  |  |
| Anxiety symptoms* | 3.66 | 2.96 | 4.53 | 2.94 | 3.01 | 0.003 | 0.29 |
| Depression symptoms* | 2.72 | 2.22 | 3.43 | 2.50 | 3.09 | 0.002 | 0.30 |
| Emotional stability * | 37.07 | 7.21 | 34.64 | 6.47 | -3.54 | <.001 | 0.36 |
| Extraversion* | 32.63 | 7.41 | 30.64 | 7.07 | -2.75 | 0.006 | 0.28 |
| Conscientiousness* | 38.50 | 5.74 | 36.74 | 6.13 | -2.92 | 0.003 | 0.30 |

For continuous variables, *p*-values are for differences calculated using Welch’s 2-sample *t*-test for continuous variables with Cohen’s *d* for standardised effect size. For categorical variables, *p*-values are for Fisher’s exact test or χ^2^ test with Yates’s continuity correction.

*Measures were recorded at the most recent full wave of data collection when participants were mean age 82 years old.

† Difference test results for Moray House Test scores at mean age 11 are based on age-adjusted values.
