## Supplementary tables 9-17 for "Impact of COVID-19 lockdown on psychosocial factors, health, and lifestyle in Scottish octogenarians: the Lothian Birth Cohort 1936 Study"

### SUPPLEMENTARY MATERIALS: Supplementary tables 9-17

Odds Ratios (95% Confidence Intervals) for all COVID-19 outcome regression models

Supplementary table 9. Odds Ratios (95% Confidence Intervals) for decreased frequency of leaving the home since COVID-19 lockdown

|  | Model 1 | Model 2 | Model 3 | Model 4 |
| --- | --- | --- | --- | --- |
| Age^a^ | 0.941 (0.718 – 1.229) | 0.952 (0.726 – 1.245) | 0.904 (0.682 – 1.194) | 0.923 (0.692 – 1.229) |
| Sex Male | Reference | Reference | Reference | Reference |
| Female | 0.633 (0.368 – 1.087) | 0.598 (0.344 – 1.033) | 0.540 (0.303 – 0.955)* | 0.557 (0.303 – 1.018) |
| Adulthood occupational social class |  | 0.666 (0.495 – 0.893)** | 0.721 (0.524 – 0.989)* | 0.707 (0.508 – 0.978)* |
| General cognitive ability |  |  | 1.437 (1.041 – 1.990)* | 1.236 (0.878 – 1.743) |
| Number of chronic diseases |  |  |  | 0.872 (0.603 – 1.258) |
| Townsend disability scale score |  |  |  | 0.726 (0.459 – 1.130) |
| Self-reported health |  |  |  | 0.621 (0.415 – 0.920)* |

**p*<.05, ***p*<.01, ****p*<.001; Independent variables are from age-82 unless otherwise stated.

**^a^** Age is age in days at time of questionnaire (mean age 84).

Odds ratios for continuous variables based on 1SD change in independent variable.

Supplementary table 10. Odds Ratios (95% Confidence Intervals) for increased internet usage since COVID-19 lockdown

|  | Model 1 | Model 2 | Model 3 | Model 4 |
| --- | --- | --- | --- | --- |
| Age^a^ | 1.02 (0.75 – 1.40) | 1.003(0.730 -1.380) | 1.03(0.73 – 1.45) | 0.97 (0.68 – 1.38) |
| Sex Male | Reference | Reference | Reference | Reference |
| Female | 2.79( 1.50 – 5.30)** | 2.50(1.29 – 4.93)** | 2.34 (1.14 – 4.85)* | 2.32 (1.12 – 4.86)* |
| Living alone^b^ Alone |  | Reference | Reference | Reference |
| Not alone |  | 0.689(0.43 – 1.11) | 0.66 (0.39 – 1.10) | 0.65 (0.38 – 1.10) |
| General cognitive ability |  |  | 1.50 (1.02 -2.24)* | 1.53 (1.03 – 2.33)* |
| Anxiety symptoms |  |  |  | 1.31 (0.92 – 1.90) |

**p*<.05, ***p*<.01, ****p*<.001; Independent variables are from age-82 unless otherwise stated.

**^a^** Age is age in days at time of questionnaire (mean age 84).

**^b^** Living alone at time of questionnaire (mean age 84).

Odds ratios for continuous variables based on 1SD change in independent variable.

Supplementary table 11. Odds Ratios (95% Confidence Intervals) for reporting a greater change in daily routine since COVID-19 lockdown

|  | Model 1 | Model 2 | Model 3 |
| --- | --- | --- | --- |
| Age^a^ | 1.066 (0.818 – 1.392) | 1.125 (0.858 – 1.479) | 1.157 (0.857 – 1.567) |
| Sex Male | Reference | Reference | Reference |
| Female | 0.861 (0.498 – 1.485) | 1.170 (0.645 – 2.135) | 1.075 (0.547 – 2.124) |
| Living alone^b^ Alone |  | Reference | Reference |
| Not alone |  | 2.012 (1.098 – 3.727)* | 1.830 (0.920 – 3.683) |
| General Health Literacy score at mean age 73 |  |  | 0.687 (0.462 – 1.014) |

**p*<.05, ***p*<.01, ****p*<.001; Independent variables are from age-82 unless otherwise stated.

**^a^** Age is age in days at time of questionnaire (mean age 84).

**^b^** Living alone at time of questionnaire (mean age 84).

Odds ratios for continuous variables based on 1SD change in independent variable.

Supplementary table 12. Odds Ratios (95% Confidence Intervals) for reporting poorer self-reported physical health since COVID-19 lockdown measures introduced

|  | Model 1 | Model 2 | Model 3 | Model 4 | Model 5 |
| --- | --- | --- | --- | --- | --- |
| Age^a^ | 1.311 (0.976 – 1.767) | 1.379 (1.018 – 1.878)* | 1.502 (1.086 – 2.095)* | 1.469 (1.058 – 2.055)* | 1.450 (1.040 – 2.036)* |
| Sex Male | Reference | Reference | Reference | Reference | Reference |
| Female | 0.517 (0.287 – 0.925) | 0.544 (0.296 – 0.992)* | 0.522 (0.270 – 0.998) | 0.560 (0.285 – 1.088) | 0.559 (0.277 – 1.113) |
| General intelligence (g) |  | 0.589 (0.422 – 0.814)** | 0.772 (0.535 – 1.108) | 0.751 (0.517 – 1.083) | 0.725 (0.496 – 1.051) |
| Number of chronic diseases |  |  | 0.981 (0.661 – 1.456) | 0.942 (0.632 – 1.402) | 0.978 (0.651 – 1.467) |
| Townsend disability scale score |  |  | 1.480 (0.870 – 2.590) | 1.327 (0.763 – 2.350) | 1.311 (0.735 – 2.368) |
| Self-rated general health |  |  | 4.672 (2.821 – 7.985)*** | 4.492 (2.630 – 7.921)*** | 3.991 (2.310 – 7.110)*** |
| Anxiety symptoms |  |  |  | 0.983 (0.692 – 1.396) | 0.838 (0.536 – 1.301) |
| Depression symptoms |  |  |  | 1.306 (0.879 – 1.957) | 1.166 (0.766 – 1.783) |
| Emotional Stability |  |  |  |  | 0.807 (0.514 – 1.258) |
| Conscientiousness |  |  |  |  | 0.825 (0.566 – 1.198) |
| Extraversion |  |  |  |  | 0.827 (0.583 – 1.172) |

**p*<.05, ***p*<.01, ****p*<.001; Independent variables are from age-82 unless otherwise stated.

**^a^** Age is age in days at time of questionnaire (mean age 84).

Odds ratios for continuous variables based on 1SD change in independent variable.

Supplementary table 13. Odds Ratios (95% Confidence Intervals) for reporting poorer self-reported mental health since COVID-19 lockdown measures introduced

|  | Model 1 | Model 2 | Model 3 | Model 4 | Model 5 |
| --- | --- | --- | --- | --- | --- |
| Age^a^ | 1.343 (1.009 – 1.793)* | 1.308 (0.979 – 1.750) | 1.304 (0.972 – 1.755) | 1.273 (0.945 – 1.718) | 1.271 (0.939 – 1.726) |
| Sex Male | Reference | Reference | Reference | Reference | Reference |
| Female | 1.272 (0.729 – 2.227) | 1.044 (0.572 – 1.903) | 1.220 (0.648 – 2.299) | 1.098 (0.579 – 2.082) | 1.109 (0.570 – 2.159) |
| Living alone^b^ Alone |  | Reference | Reference | Reference | Reference |
| Not alone |  | 0.561 (0.307 – 1.017) | 0.582 (0.313 – 1.074) | 0.609 (0.325 – 1.132) | 0.533 (0.277 – 1.015) |
| Number of chronic diseases |  |  | 1.201 (0.836 – 1.730) | 1.113 (0.771 – 1.607) | 1.201 (0.824 – 1.754) |
| Self-rated general health |  |  | 2.015 (1.389 – 2.973)*** | 1.625 (1.099 – 2.434)* | 1.483 (0.992 – 2.238) |
| Anxiety symptoms* |  |  |  | 1.694 (1.213 – 2.381)** | 1.148 (0.760 – 1.732) |
| Depression symptoms* |  |  |  | 1.174 (0.832 – 1.664) | 1.033 (0.712 – 1.503) |
| Emotional Stability |  |  |  |  | 0.535 (0.351 – 0.806)** |
| Extraversion |  |  |  |  | 0.892 (0.641 – 1.242) |

**p*<.05, ***p*<.01, ****p*<.001; Independent variables are from age-82 unless otherwise stated.

**^a^** Age is age in days at time of questionnaire (mean age 84).

**^b^** Living alone at time of questionnaire (mean age 84).

Odds ratios for continuous variables based on 1SD change in independent variable.

Supplementary table 14. Odds Ratios (95% Confidence Intervals) for experiencing COVID-19 related stress or nervousness during COVID-19 lockdown

|  | Model 1 | Model 2 | Model 3 | Model 4 |
| --- | --- | --- | --- | --- |
| Age^a^ | 0.98 (0.72 – 1.32) | 0.95 (0.70 – 1.29) | 0.92(0.66 – 1.29) | 0.94 (0.66 – 1.33) |
| Sex Male | Reference | Reference | Reference | Reference |
| Female | 1.71 (0.94 – 3.13) | 1.33 (0.69 – 2.53) | 1.24 (0.62 – 2.48) | 1.55 (0.73 – 3.31) |
| Living alone^b^ Alone |  | Reference | Reference | Reference |
| Not alone |  | 0.60 (0.38– 0.95)* | 0.62 (0.38 – 1.01) | 0.65 (0.38 – 1.11) |
| Anxiety symptoms |  |  | 1.75 (1.26 – 2.49)** | 0.99 (0.63 – 1.55) |
| Emotional stability |  |  |  | 0.40 (0.24 – 0.62)*** |

**p*<.05, ***p*<.01, ****p*<.001; Independent variables are from age-82 unless otherwise stated.

**^a^** Age is age in days at time of questionnaire (mean age 84).

**^b^** Living alone at time of questionnaire (mean age 84).

Odds ratios for continuous variables based on 1SD change in independent variable.

Supplementary table 15. Odds Ratios (95% Confidence Intervals) for experiencing loneliness during COVID-19 lockdown

|  | Model 1 | Model 2 | Model 3 | Model 4 | Model 5 |
| --- | --- | --- | --- | --- | --- |
| Age^a^ | 1.20 (0.999 – 1.006) | 1.13 (0.77 – 1.65) | 1.06 (0.71 – 1.61) | 1.06 (0.69 – 1.64) | 1.04 (0.68 – 1.62) |
| Sex Male | Reference | Reference | Reference | Reference | Reference |
| Female | 1.37 (0.71 – 2.84) | 0.66 (0.29 – 1.44) | 0.53 (0.21 – 1.27) | 0.41 (0.15 – 1.06) | 0.48 (0.17 – 1.26) |
| Living alone^b^ Alone |  | Reference | Reference | Reference | Reference |
| Not alone |  | 0.21 (0.11– 0.38)*** | 0.15 (0.07 – 0.30)*** | 0.14 (0.06 – 0.29)*** | 0.15 (0.07 – 0.31)*** |
| Townsend Disability Scale score |  |  | 1.64 (0.95– 2.90) | 1.79 (1.01 – 3.33) | 1.67 (0.92 – 3.14) |
| Self-rated general health Excellent |  |  | 0.10 (0.001 – 7.14) | 0.18 (0.001 – 26.09) | 0.17 (0.001 – 26.69) |
| Very good |  |  | 0.66 (0.01 – 32.39) | 0.76 (0.01 – 73.66) | 0.64 (0.01 – 65.46) |
| Good |  |  | 0.44 (0.01 – 21.26) | 0.42 (0.004 – 38.993) | 0.34 (0.003 – 33.48) |
| Fair |  |  | 5.48 (0.10 – 321.55) | 3.14 (0.03 – 356.73) | 3.32 (0.03 – 399.82) |
| Poor |  |  | Reference | Reference | Reference |
| Anxiety symptoms |  |  |  | 1.99 (1.26 – 3.27)** | 1.76 (1.01 – 3.14)* |
| Emotional stability |  |  |  |  | 0.76 (0.45 – 1.24) |

**p*<.05, ***p*<.01, ****p*<.001; Independent variables are from age-82 unless otherwise stated.

**^a^** Age is age in days at time of questionnaire (mean age 84).

**^b^** Living alone at time of questionnaire (mean age 84).

Odds ratios for continuous variables based on 1SD change in independent variable.

Supplementary table 16. Odds Ratios (95% Confidence Intervals) for a decrease in physical activity since COVID-19 lockdown measures introduced

|  | Model 1 | Model 2 | Model 3 |
| --- | --- | --- | --- |
| Age^a^ | 1.058 (0.807 – 1.389) | 1.054 (0.803 – 1.387) | 1.133 (0.854 – 1.506) |
| Sex Male | Reference | Reference | Reference |
| Female | 1.024 (0.601 – 1.748) | 1.133 (0.660 – 1.950) | 1.076 (0.614 – 1.891) |
| Adulthood occupational class |  | 1.586 (1.185 – 2.134)** | 1.426 (1.043 – 1.961)* |
| General cognitive ability (g) |  |  | 0.679 (0.491 – 0.931)* |

**p*<.05, ***p*<.01, ****p*<.001; Independent variables are from age-82 unless otherwise stated.

**^a^** Age is age in days at time of questionnaire (mean age 84).

Odds ratios for continuous variables based on 1SD change in independent variable.

Supplementary table 17. Odds Ratios (95% Confidence Intervals) for returning to or starting a new pastime since COVID-19 lockdown measures introduced

|  | Model 1 | Model 2 |
| --- | --- | --- |
| Age^a^ | 0.90 (0.67 – 1.22) | 0.92 (0.66 – 1.28) |
| Sex Male | Reference | Reference |
| Female | 2.02 (1.10 – 3.75)* | 1.89 (0.96 – 3.75) |
| General health literacy at age 73 |  | 1.36 (0.92– 2.02) |

**p*<.05, ***p*<.01, ****p*<.001; Independent variables are from age-82 unless otherwise stated.

**^a^** Age is age in days at time of questionnaire (mean age 84).

Odds ratios for continuous variables based on 1SD change in independent variable.
