## Appendix 1 for "Impact of COVID-19 lockdown on psychosocial factors, health, and lifestyle in Scottish octogenarians: the Lothian Birth Cohort 1936 Study"

LBC1936 COVID-19 Questionnaire

Start of Block: Knowledge of COVID-19

First, we would like to ask you about your knowledge and experience of the pandemic.

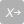

How would you rate your knowledge of COVID-19 (e.g. the symptoms of the disease, how it is transmitted, and how to minimise your exposure to it)?

- Extremely good (1)
- Somewhat good (2)
- Neither good nor bad (3)
- Somewhat bad (4)
- Extremely bad (5)

Do you find the Scottish Government’s guidance on COVID-19 easy to understand?

- Extremely easy (1)
- Somewhat easy (2)
- Neither easy nor difficult (3)
- Somewhat difficult (4)
- Extremely difficult (5)
- I am not aware of the government guidance on COVID-19 (6)

| Page Break |
| --- |

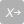

Have you been following the government guidance on:

|  | Always (1) | Most of the time (2) | Some of the time (3) | Never (4) | N/A (0) |
| --- | --- | --- | --- | --- | --- |
| Social distancing (1) |  |  |  |  |  |
| Staying at home as much as possible (2) |  |  |  |  |  |
| Self-isolating (if suffering COVID-19 symptoms) (3) |  |  |  |  |  |
| Hand washing (4) |  |  |  |  |  |

| Page Break |
| --- |

How often do you keep up to date on COVID-19 related news?

- Multiple times per day (1)
- Daily (2)
- Every few days (3)
- Weekly (4)
- Less than weekly (5)

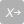

Which of the following sources of information have you used to keep informed about COVID-19? Select all that apply, or select 'None of the above'.

- BBC news television bulletins (1)
- Other television news bulletins (e.g., ITV, Channel 4) (2)
- BBC news website (3)
- Other news websites (e.g., ITV, Channel 4) (4)
- NHS websites (5)
- GP practice website (6)
- Government websites (7)
- World Health Organisation (WHO) website (8)
- Broadsheet newspapers (print or website) (9)
- Tabloid newspapers (print or website) (10)
- Radio or podcasts (11)
- Social media websites and news feeds (e.g., Instagram, Facebook, Twitter) (12)
- Your workplace (13)
- Family and friends (14)
- WhatsApp or other messaging services (15)
- Other health websites and resources (16)
- ⊗None of the above (0)

Display This Question:

If Which of the following sources of information have you used to keep informed about COVID-19? Sele... != None of the above

Carry Forward Selected Choices from 'Which of the following sources of information have you used to keep informed about COVID-19? Select all that apply, or select 'None of the above'.'

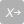

Which of the following sources of information have you found the most helpful for keeping informed about COVID-19?

- BBC news television bulletins (1)
- Other television news bulletins (e.g., ITV, Channel 4) (2)
- BBC news website (3)
- Other news websites (e.g., ITV, Channel 4) (4)
- NHS websites (5)
- GP practice website (6)
- Government websites (7)
- World Health Organisation (WHO) website (8)
- Broadsheet newspapers (print or website) (9)
- Tabloid newspapers (print or website) (10)
- Radio or podcasts (11)
- Social media websites and news feeds (e.g., Instagram, Facebook, Twitter) (12)
- Your workplace (13)
- Family and friends (14)
- WhatsApp or other messaging services (15)
- Other health websites and resources (16)
- ⊗None of the above (0)

End of Block: Knowledge of COVID-19

Start of Block: Experience of COVID-19

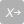

Do you think that you have had, or currently have COVID-19?

- Yes, confirmed by a positive test (2)
- Yes, suspected COVID-19 but was not tested	(1)
- No (0)

Display This Question:

If Do you think that you have had, or currently have COVID-19? = Yes, suspected COVID-19 but was not tested

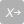

If COVID-19 was suspected but not tested, was it:

- Diagnosed by a medical professional based on symptoms (1)
- Self-diagnosis based on symptoms (2)

Display This Question:

If Do you think that you have had, or currently have COVID-19? != No

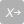

Which symptoms did you have? (select all that apply)

- Fever (1)
- Dry cough (2)
- Shortness of breath (3)
- Headaches (4)
- Aches and pains (5)
- Sore throat (6)
- Fatigue (7)
- Diarrhoea (8)
- Runny nose (9)
- Sneezing (10)
- Loss of taste (11)
- Loss of smell (12)
- Chilblain-like skin lesions (13)
- Other symptoms (0)

Display This Question:

If Which symptoms did you have? (select all that apply) = Other symptoms

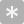

If you responded 'other symptoms', please give details:

________________________________________________________________

Display This Question:

If Do you think that you have had, or currently have COVID-19? != No

Carry Forward Selected Choices from 'Which symptoms did you have? (select all that apply)'

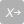

How long did each of your symptoms last?

|  | A few days (up to one week) (1) | One to two weeks (2) | Two to three weeks (3) | Three to four weeks (4) | More than one month (5) |
| --- | --- | --- | --- | --- | --- |
| Fever (x228) |  |  |  |  |  |
| Dry cough (x229) |  |  |  |  |  |
| Shortness of breath (x230) |  |  |  |  |  |
| Headaches (x231) |  |  |  |  |  |
| Aches and pains (x232) |  |  |  |  |  |
| Sore throat (x233) |  |  |  |  |  |
| Fatigue (x234) |  |  |  |  |  |
| Diarrhoea (x235) |  |  |  |  |  |
| Runny nose (x236) |  |  |  |  |  |
| Sneezing (x237) |  |  |  |  |  |
| Loss of taste (x238) |  |  |  |  |  |
| Loss of smell (x239) |  |  |  |  |  |
| Chilblain-like skin lesions (x240) |  |  |  |  |  |
| Other symptoms (x241) |  |  |  |  |  |

| Page Break |
| --- |

Display This Question:

If Do you think that you have had, or currently have COVID-19? != No

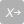

During the time that you had or suspected you had COVID-19 did you: (select all that apply)

- Do nothing, I’m already shielding (0)
- Self-isolate or distance myself from others in the household (1)
- Call NHS 111 or my GP (2)
- Get swabbed for COVID-19 (3)
- Visit a COVID Hub to see a doctor (4)
- Get admitted to hospital	(5)

Display This Question:

If During the time that you had or suspected you had COVID-19 did you: (select all that apply) = Get admitted to hospital

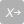

Were you admitted to an intensive care unit?

- Yes (1)
- No (0)
- Don’t know (2)

Display This Question:

If During the time that you had or suspected you had COVID-19 did you: (select all that apply) = Get admitted to hospital

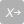

Did you need to have a ventilator to help you breathe?

- Yes (1)
- No (0)
- Don’t know (2)

| Page Break |
| --- |

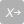

Do you think anyone else in your household has had or currently has COVID-19?

- Yes, confirmed by a positive test (2)
- Yes, suspected COVID-19 but was not tested	(1)
- No (0)

Display This Question:

If Do you think anyone else in your household has had or currently has COVID-19? = Yes, suspected COVID-19 but was not tested

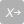

If COVID-19 was suspected but not tested, was it:

- Diagnosed by a medical professional based on symptoms (1)
- Self-diagnosis based on symptoms (2)

| Page Break |
| --- |

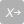

Have you been contacted by letter or text message to say you are at severe risk from COVID-19 due to an underlying health condition and should be shielding?

- Yes (1)
- No (0)

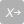

Has **anxiety about COVID-19** caused you to avoid or postpone contacting or attending a medical service (this does not include appointments cancelled by the service provider)?

- Yes, postponed contacting a medical service (such as calling my GP) (4)
- Yes, postponed attending a medical appointment that was made previously (3)
- I was anxious about COVID-19 but did not avoid or postpone contacting or attending a medical service (2)
- ⊗No, I have not been anxious about contacting or attending a medical service (1)
- ⊗Not applicable (0)

End of Block: Experience of COVID-19

Start of Block: Household

We would like to ask you some questions about your household.

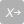

Including yourself, how many people currently live in your household?

- 1 (1)
- 2 (2)
- 3 (3)
- 4 (4)
- 5 (5)
- 6 (6)
- 7 (7)
- 8 (8)
- 9 (9)
- 10 or more (10)

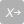

Who currently lives in your household with you? (select all that apply)

- ⊗I live alone (1)
- Spouse/partner (2)
- Child/children (3)
- Grandchildren/Great grandchildren (4)
- Parent(s) or parent(s)-in-law (5)
- Other family member(s) (6)
- Paid caregiver(s) (7)
- Friend(s) or other non-family member(s) (8)

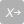

Has your living situation changed because of COVID-19? (e.g., has your place of residence or the number of people living in your household changed?)

- Yes (1)
- No (0)

Display This Question:

If Has your living situation changed because of COVID-19? (e.g., has your place of residence or the... = Yes

| 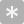 | 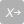 |
| --- | --- |

If yes, has your living situation changed in any of the following ways? (select all that apply)

- Living in a different house (1)
- Living with more people (2)
- Living with fewer people (3)

| Page Break |
| --- |

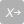

What kind of area do you live in?

- Rural (1)
- Urban (2)
- Suburban (3)

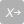

How often have you been leaving your home since COVID-19 measures were introduced (23rd March 2020)?

- Multiple times a day (6)
- Once per day (5)
- A few times per week (4)
- Once per week (3)
- Less than once per week (e.g. once per fortnight) (2)
- Never, since COVID-19 measures were introduced. (1)

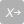

When leaving your home for a daily walk or other exercise, how likely are you to accidentally come into close contact with someone not living in your household (e.g. less than 2 metres)?

- I don’t leave my home (1)
- Not at all likely (2)
- Not that likely (3)
- Somewhat likely (4)
- Very likely (5)

| Page Break |
| --- |

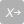

Do you have any pets?

- Yes (1)
- No (0)

Display This Question:

If Do you have any pets? = Yes

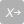

If yes, what kind of pet(s) do you have?

- Dog(s) (1)
- Cat(s) (2)
- Other pet(s) (3)

Display This Question:

If If yes, what kind of pet(s) do you have? = Other pet(s)

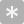

If 'other pet(s)', please give details:

________________________________________________________________

| Page Break |
| --- |

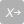

Have you been helping to care for grandchildren or great-grandchildren since COVID-19 measures were introduced (23rd March 2020)? (e.g. while their parents are at work)

- Yes (1)
- No (0)

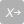

Are you currently a carer (i.e., do you provide regular help and support to someone who is less able to cope alone)?

- Yes, for someone who lives with me (2)
- Yes, for someone who does not live with me (1)
- ⊗No, not a carer (0)

Display This Question:

If Are you currently a carer (i.e., do you provide regular help and support to someone who is less a... != No, not a carer

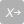

If yes, were you a carer **before** COVID-19 measures were introduced (23rd March 2020)? (i.e., did you provide regular help and support to someone who is less able to cope alone)?

- Yes, for someone who lives with me (2)
- Yes, for someone who does not live with me (1)
- ⊗No, not a carer (0)

Display This Question:

If Are you currently a carer (i.e., do you provide regular help and support to someone who is less a... != No, not a carer

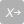

If yes, has your experience of caring changed since COVID-19 measures were introduced?

- More challenging now (4)
- No change (3)
- Less challenging now (2)
- Not possible now (1)

End of Block: Household

Start of Block: Medicine and shopping

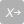

What actions have you taken to continue to get any prescription medicines since COVID-19 measures were introduced (23rd March 2020)?

- ⊗None (1)
- Switched order to mail order or delivery (2)
- Increased the prescription to a longer supply (e.g. 90-day supply) (3)
- Other (4)
- ⊗N/A (0)

Display This Question:

If What actions have you taken to continue to get any prescription medicines since COVID-19 measures... = Other

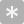

If you responded 'other', please give details about actions you have taken to continue to get any prescription medicines:

________________________________________________________________

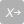

Have you received any additional help in your daily life with things such as grocery shopping, errands, or picking up medications since COVID-19 measures were introduced? (Here, we are referring to new help, not previously provided before the COVID-19 pandemic)

- Yes (1)
- No (0)

Display This Question:

If Have you received any additional help in your daily life with things such as grocery shopping, er... = Yes

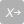

If yes, what additional help have you received?

- Grocery shopping (1)
- Picking up medicines (prescription or over the counter) (2)
- Other (3)

Display This Question:

If If yes, what additional help have you received? = Other

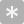

If you answered 'other', please give details about the additional help you have received:

________________________________________________________________

| Page Break |
| --- |

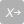

Have you been shopping for others (i.e. anyone who does not live with you)?

- Yes (1)
- No (0)

| Page Break |
| --- |

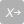

Compared to before COVID-19 measures were introduced (23rd March 2020), have you found the need to use more non-cash alternatives such as paying for groceries by credit/debit card?

- Yes (1)
- No (0)

How important is it for you to be able to use cash?

- Very important (1)
- Quite important (2)
- Not very important (3)
- Not important at all (4)

End of Block: Medicine and shopping

Start of Block: Social contact

We would like to ask some questions about your social contact.

Just before the COVID-19 measures were introduced (23rd March 2020), how regularly did you:

|  | Every day/almost every day (1) | 3-4 days a week (2) | 1-2 days a week (3) | Less than once a week (4) | Rarely (5) | Never (6) |
| --- | --- | --- | --- | --- | --- | --- |
| Meet with family members face-to-face (1) |  |  |  |  |  |  |
| Meet with friends face-to-face (2) |  |  |  |  |  |  |
| Call family members (3) |  |  |  |  |  |  |
| Call friends (4) |  |  |  |  |  |  |
| Video call with family members (e.g., Skype, FaceTime) (5) |  |  |  |  |  |  |
| Video call with friends (e.g., Skype, FaceTime) (6) |  |  |  |  |  |  |
| Text or instant message (e.g., WhatsApp, Facebook Messenger) with family members (7) |  |  |  |  |  |  |
| Text or instant message (e.g., WhatsApp, Facebook Messenger) with friends (8) |  |  |  |  |  |  |

| Page Break |
| --- |

Now that the COVID-19 measures are in place, how regularly do you do these activities:

|  | Every day/almost every day (1) | 3-4 days a week (2) | 1-2 days a week (3) | Less than once a week (4) | Rarely (5) | Never (6) |
| --- | --- | --- | --- | --- | --- | --- |
| Meet with family members face-to-face (1) |  |  |  |  |  |  |
| Meet with friends face-to-face (2) |  |  |  |  |  |  |
| Call family members (3) |  |  |  |  |  |  |
| Call friends (4) |  |  |  |  |  |  |
| Video call with family members (e.g., Skype, FaceTime) (5) |  |  |  |  |  |  |
| Video call with friends (e.g., Skype, FaceTime) (6) |  |  |  |  |  |  |
| Text or instant message (e.g., WhatsApp, Facebook Messenger) with family members (7) |  |  |  |  |  |  |
| Text or instant message (e.g., WhatsApp, Facebook Messenger) with friends (8) |  |  |  |  |  |  |

| Page Break |
| --- |

Since the COVID-19 measures were introduced (23rd March 2020), has contact with your neighbours changed?

- More contact now (1)
- The same (2)
- Less contact now (3)

Display This Question:

If Since the COVID-19 measures were introduced (23rd March 2020), has contact with your neighbours c... != The same

If contact with your neighbours has changed, how have you found this experience?

- Positive (1)
- Neutral (2)
- Negative	(3)

| Page Break |
| --- |

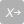

Are there any local initiatives in your neighbourhood or broader community to help those who are self-isolating e.g. getting shopping, medications etc.?

- Yes (1)
- No (0)
- Don't know (2)

Display This Question:

If Are there any local initiatives in your neighbourhood or broader community to help those who are... = Yes

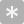

If yes, please give details about local initiatives in your community to help those who are self-isolating:

________________________________________________________________

End of Block: Social contact

Start of Block: Mental health

We would like to ask some questions about your mental health.

In general, **before** the COVID-19 measures were introduced (23rd March 2020), would you say your emotional and mental health was:

- Excellent (1)
- Very good (2)
- Good (3)
- Fair (4)
- Poor	(5)

In general, **since** the COVID-19 measures were introduced, would you say your emotional and mental health is:

- Excellent (1)
- Very good (2)
- Good (3)
- Fair (4)
- Poor	(5)

In the last **two weeks**, how often have you felt nervous or stressed because of COVID-19?

- Never (1)
- Some of the time (2)
- Most of the time (3)
- All of the time (4)

End of Block: Mental health

Start of Block: Health & Health Behaviours

We would like to ask some questions about your health.

In general, **before** the COVID-19 measures were introduced (23rd March 2020), would you say your physical health was:

- Excellent (1)
- Very good (2)
- Good (3)
- Fair (4)
- Poor	(5)

In general, **since** the COVID-19 measures were introduced, would you say your physical health is:

- Excellent (1)
- Very good (2)
- Good (3)
- Fair (4)
- Poor	(5)

End of Block: Health & Health Behaviours

Start of Block: Alcohol

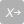

Do you ever drink alcohol?

- Yes (1)
- No (0)
- Special occasions only (less than once per month) (99)

Display This Question:

If Do you ever drink alcohol? = Yes

If yes, compared to before COVID-19 measures were introduced are you:

- Drinking more alcohol now (1)
- Drinking about the same amount of alcohol now (2)
- Drinking less alcohol now (3)

End of Block: Alcohol

Start of Block: Smoking

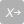

What is your current smoking status (**cigarettes only**)?

- Smoker (2)
- Ex-smoker (1)
- Never smoker (0)

Display This Question:

If What is your current smoking status (cigarettes only)? = Smoker

If you are a current smoker, compared to before COVID-19 measures were introduced are you:

- Smoking more now (1)
- Smoking about the same now	(2)
- Smoking less now	(3)

End of Block: Smoking

Start of Block: Diet

Compared to before COVID-19 measures were introduced (23rd March 2020), is your diet:

- Much healthier now (1)
- Slightly healthier now (2)
- About the same now (3)
- Slightly less healthy now (4)
- Much less healthy now (5)

How much are you eating compared to before the COVID-19 measures were introduced?

- More now (1)
- About the same (2)
- Less now (3)

End of Block: Diet

Start of Block: Exercise

Compared to before COVID-19 measures were introduced (23rd March 2020), how much physical activity are you doing now? This includes activities that make you breathe harder than normal (e.g., brisk walking).

- Much more physical activity now (1)
- Slightly more physical activity now (2)
- About the same physical activity now (3)
- Slightly less physical activity now (4)
- Much less physical activity now (5)

End of Block: Exercise

Start of Block: Activities

How much has COVID-19 changed your daily routine?

- A lot (1)
- Somewhat (2)
- A little (3)
- Not at all (4)

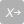

Since COVID-19 measures have been in place (23rd March 2020), have you returned to or started up a new pastime that you can do from home? (Select all that apply, or select 'No change in activities')

- Work out or exercise	(1)
- Relaxation or meditation	(2)
- Cooking class/learning to cook	(3)
- Language class/learning a language	(4)
- Arts and crafts	(5)
- Singing (6)
- Playing music (7)
- Listening to music	(8)
- Reading	(9)
- Board or card games	(10)
- Online gaming	(11)
- Writing for pleasure (12)
- Watching television/films (13)
- Listening to the radio/audiobooks (14)
- Baking (15)
- Gardening (16)
- Dancing (17)
- Online educational course (18)
- Other activity not listed above	(19)
- ⊗No change in activities (0)

Display This Question:

If Since COVID-19 measures have been in place (23rd March 2020), have you returned to or started up... = Other activity not listed above

If you responded 'Other activity not listed above', please give details about your new pastime(s):

________________________________________________________________

| Page Break |
| --- |

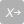

Do you have a garden (including access to a shared garden) or allotment?

- Yes, garden (1)
- Yes, allotment (2)
- ⊗No (0)

Display This Question:

If Do you have a garden (including access to a shared garden) or allotment? != No

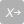

If yes, what do you do in the garden/allotment? (Select all that apply).

- Gardening (1)
- Exercise (2)
- Relax (3)
- Other activity (6)
- ⊗Not using currently (4)

Display This Question:

If If yes, what do you do in the garden/allotment? (Select all that apply). = Other activity

If you responded 'Other activity', please give details about what you do in the garden/allotment:

________________________________________________________________

Display This Question:

If Do you have a garden (including access to a shared garden) or allotment? != No

Compared to before COVID-19 measures were introduced, how often do you currently use your garden/allotment?

- Much more often now (1)
- Slightly more often now (2)
- About the same now (3)
- Slightly less now (4)
- Much less now (5)

| Page Break |
| --- |

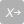

How has your internet usage changed since COVID-19 measures were introduced (23rd March 2020)?

- I use the internet much more now (1)
- I use the internet slightly more now (2)
- My internet usage is about the same (3)
- I use the internet slightly less now (4)
- I use the internet much less now (5)
- Not applicable (0)

Display This Question:

If How has your internet usage changed since COVID-19 measures were introduced (23rd March 2020)?  = I use the internet much more now

Or How has your internet usage changed since COVID-19 measures were introduced (23rd March 2020)?  = I use the internet slightly more now

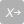

If you are using the internet more, do you think you will continue to use the internet more often after the COVID-19 emergency has passed?

- Yes (1)
- No (0)
- Don’t know (2)

End of Block: Activities

Start of Block: Repeated measures from the LBC1936 questionnaire booklet

Finally, we’d like you to complete some questionnaires that you usually complete as part of the LBC1936 questionnaire booklet and that you last completed at Wave 5 of the study.
 We understand that some of these will be difficult to answer in the current circumstances, and that your answers to some of the questions might be restricted for the moment due to the current guidelines limiting opportunities for social contact and to be outdoors, but please try to answer the questions as best as you can.
Please respond to each question in a way that best describes your current situation or experience, now that the COVID-19 measures are in place.

End of Block: Repeated measures from the LBC1936 questionnaire booklet

Start of Block: Physical activity

You are going to be asked to rate how physically active you generally are at present. Please select the response which best indicates the level of sport or exercise you have been mainly participating in since COVID-19 measures were introduced (23rd March 2020).

What level of physical activity do you mainly do? (Select only one)

- Moving only in connection with necessary (household) chores (1)
- Walking or other outdoor activities 1-2 times per week (2)
- Walking or other outdoor activities several times per week (3)
- Exercising 1-2 times per week to the point of perspiring and heavy breathing (4)
- Exercising several times per week to the point of perspiring and heavy breathing (5)
- Keep-fit/heavy exercise or competitive sport several times per week (6)

End of Block: Physical activity

Start of Block: The Warwick-Edinburgh Mental Well-being Scale

Below are some statements about feelings and thoughts. Please tick the box that best describes your experience of each over the last 2 weeks.

I’ve been feeling optimistic about the future

- None of the time (1)
- Rarely (2)
- Some of the time (3)
- Often (4)
- All of the time (5)

I’ve been feeling useful

- None of the time (1)
- Rarely (2)
- Some of the time (3)
- Often (4)
- All of the time (5)

I’ve been feeling relaxed

- None of the time (1)
- Rarely (2)
- Some of the time (3)
- Often (4)
- All of the time (5)

I’ve been feeling interested in other people

- None of the time (1)
- Rarely (2)
- Some of the time (3)
- Often (4)
- All of the time (5)

I’ve had energy to spare

- None of the time (1)
- Rarely (2)
- Some of the time (3)
- Often (4)
- All of the time (5)

I’ve been dealing with problems well

- None of the time (1)
- Rarely (2)
- Some of the time (3)
- Often (4)
- All of the time (5)

I’ve been thinking clearly

- None of the time (1)
- Rarely (2)
- Some of the time (3)
- Often (4)
- All of the time (5)

I’ve been feeling good about myself

- None of the time (1)
- Rarely (2)
- Some of the time (3)
- Often (4)
- All of the time (5)

I’ve been feeling close to other people

- None of the time (1)
- Rarely (2)
- Some of the time (3)
- Often (4)
- All of the time (5)

I’ve been feeling confident

- None of the time (1)
- Rarely (2)
- Some of the time (3)
- Often (4)
- All of the time (5)

I’ve been able to make up my own mind about things

- None of the time (1)
- Rarely (2)
- Some of the time (3)
- Often (4)
- All of the time (5)

I’ve been feeling loved

- None of the time (1)
- Rarely (2)
- Some of the time (3)
- Often (4)
- All of the time (5)

I’ve been interested in new things

- None of the time (1)
- Rarely (2)
- Some of the time (3)
- Often (4)
- All of the time (5)

I’ve been feeling cheerful

- None of the time (1)
- Rarely (2)
- Some of the time (3)
- Often (4)
- All of the time (5)

End of Block: The Warwick-Edinburgh Mental Well-being Scale

Start of Block: Perceived Social Support Scale

We would now like you to think about your family and friends. By family we mean those you live with as well as those elsewhere. Here are some comments people have made about their family and friends. We’d like you to indicate how far each statement is true for you since COVID-19 measures were introduced (23 March 2020).

There are people I know amongst my family or friends who do things to make me happy

- Not true (1)
- Partly true (2)
- Certainly true (3)

There are people I know amongst my family and friends who make me feel loved

- Not true (1)
- Partly true (2)
- Certainly true (3)

There are people I know amongst my family and friends who can be relied upon no matter what happens

- Not true (1)
- Partly true (2)
- Certainly true (3)

There are people I know amongst my family and friends who would see that I am taken care of if I needed to be

- Not true (1)
- Partly true (2)
- Certainly true (3)

There are people I know amongst my family and friends who accept me just as I am

- Not true (1)
- Partly true (2)
- Certainly true (3)

There are people I know amongst my family and friends who make me feel an important part of their lives

- Not true (1)
- Partly true (2)
- Certainly true (3)

There are people I know amongst my family and friends who give me support and encouragement

- Not true (1)
- Partly true (2)
- Certainly true (3)

End of Block: Perceived Social Support Scale

Start of Block: Loneliness

How often have you felt lonely **during the past week**?

- None or almost none of the time (1)
- Some of the time (2)
- Most of the time (3)
- All or almost all of the time (4)

End of Block: Loneliness

Start of Block: Neighbourhood questions

The following statements are about neighbourhoods. Please indicate how strongly you agree or disagree with each statement.

I feel like I belong to this neighbourhood

- Strongly Agree (1)
- Agree (2)
- Neither Agree or Disagree (3)
- Disagree (4)
- Strongly Disagree (5)

The friendships and associations I have with other people in my neighbourhood mean a lot to me

- Strongly Agree (1)
- Agree (2)
- Neither Agree or Disagree (3)
- Disagree (4)
- Strongly Disagree (5)

If I needed advice about something I could go to someone in my neighbourhood

- Strongly Agree (1)
- Agree (2)
- Neither Agree or Disagree (3)
- Disagree (4)
- Strongly Disagree (5)

I borrow things and exchange favours with my neighbours

- Strongly Agree (1)
- Agree (2)
- Neither Agree or Disagree (3)
- Disagree (4)
- Strongly Disagree (5)

I would be willing to work together with others on something to improve my neighbourhood

- Strongly Agree (1)
- Agree (2)
- Neither Agree or Disagree (3)
- Disagree (4)
- Strongly Disagree (5)

I plan to remain a resident of this neighbourhood for a number of years

- Strongly Agree (1)
- Agree (2)
- Neither Agree or Disagree (3)
- Disagree (4)
- Strongly Disagree (5)

I like to think of myself as similar to the people who live in this neighbourhood

- Strongly Agree (1)
- Agree (2)
- Neither Agree or Disagree (3)
- Disagree (4)
- Strongly Disagree (5)

I regularly stop and talk with people in my neighbourhood

- Strongly Agree (1)
- Agree (2)
- Neither Agree or Disagree (3)
- Disagree (4)
- Strongly Disagree (5)

End of Block: Neighbourhood questions

Start of Block: Sleep Questionnaire

We would like to ask you some questions about your sleeping behaviour.  Please tick one of the following boxes to indicate your response.
During the past month, how would you rate your sleep quality overall?

- Very good (1)
- Fairly good (2)
- Fairly bad (3)
- Very bad (4)

Compared to before COVID-19 measures were introduced, is the quality of your sleep:

- Much better now (1)
- Somewhat better now (2)
- About the same now (3)
- Somewhat worse now (4)
- Much worse now (5)

| Page Break |
| --- |

For the following questions, please provide an answer in relation to **weeknights**.

On weeknights, what time do you usually go to bed at night?*Please give your answers in* ***24 hour time*** *format, for example: 23:30*

________________________________________________________________

On weeknights, how long does it take you to fall asleep at night? *Please answer in number of* ***hours and minutes****, for example: 0 hours, 45 minutes****.***

- Number of hours: (4) ________________________________________________
- Number of minutes: (5) ________________________________________________

On weekdays, What time do you usually get up in the morning?*Please give your answers in* ***24 hour time*** *format, for example: 07:45*

________________________________________________________________

How many hours of actual sleep do you usually get at night on **weeknights**? (This may be different than the number of hours you spend in bed)*Please answer in number of hours and minutes, for example: 8 hours, 30 minutes*

- Number of hours: (4) ________________________________________________
- Number of minutes: (5) ________________________________________________

How long do you usually spend asleep during the day and evening on **weekdays**?
*Please answer in number of hours and minutes, for example: 2 hours 30 minutes*

- Number of hours: (4) ________________________________________________
- Number of minutes: (5) ________________________________________________

| Page Break |
| --- |

For the following questions, please provide an answer in relation to **weekends.**

On **weekends,** what time do you usually go to bed at night?*Please give your answers in* ***24 hour time*** *format, for example: 23:30*

________________________________________________________________

On **weekends,** how long does it take you to fall asleep at night? *Please answer in number of hours and minutes, for example: 0 hours 45 minutes*

- Number of hours: (4) ________________________________________________
- Number of minutes: (5) ________________________________________________

On **weekends,** what time do you usually get up in the morning?*Please give your answers in* ***24 hour time*** *format, for example: 07:45*

________________________________________________________________

How many hours of actual sleep do you usually get at night on **weekends**? (This may be different than the number of hours you spend in bed)*Please answer in number of hours and minutes, for example: 8 hours 30 minutes*

- Number of hours: (4) ________________________________________________
- Number of minutes: (5) ________________________________________________

How long do you usually spend asleep during the day and evening on **weekends**?
*Please answer in number of hours and minutes, for example: 2 hours 30 minutes*

- Number of hours: (4) ________________________________________________
- Number of minutes: (5) ________________________________________________

End of Block: Sleep Questionnaire

Start of Block: Experiences and attitudes to ageing questionnaire

The following statements are about attitude to and experience of getting older. Please indicate how strongly you agree or disagree with each statement.

It is a privilege to grow old.

- Strongly disagree (1)
- Disagree (2)
- Uncertain (3)
- Agree (4)
- Strongly agree (5)

There are many pleasant things about growing older.

- Strongly disagree (1)
- Disagree (2)
- Uncertain (3)
- Agree (4)
- Strongly agree (5)

Old age is a depressing time of life.

- Strongly disagree (1)
- Disagree (2)
- Uncertain (3)
- Agree (4)
- Strongly agree (5)

I don’t feel old.

- Strongly disagree (1)
- Disagree (2)
- Uncertain (3)
- Agree (4)
- Strongly agree (5)

I see old age mainly as a time of loss.

- Strongly disagree (1)
- Disagree (2)
- Uncertain (3)
- Agree (4)
- Strongly agree (5)

I have more energy now than I expected for my age.

- Strongly disagree (1)
- Disagree (2)
- Uncertain (3)
- Agree (4)
- Strongly agree (5)

As I get older, I find it more difficult to make new friends.

- Strongly disagree (1)
- Disagree (2)
- Uncertain (3)
- Agree (4)
- Strongly agree (5)

It is very important to pass on the benefits of my experiences to younger people.

- Strongly disagree (1)
- Disagree (2)
- Uncertain (3)
- Agree (4)
- Strongly agree (5)

I want to give a good example to younger people.

- Strongly disagree (1)
- Disagree (2)
- Uncertain (3)
- Agree (4)
- Strongly agree (5)

I feel excluded from things because of my age.

- Strongly disagree (1)
- Disagree (2)
- Uncertain (3)
- Agree (4)
- Strongly agree (5)

My health is better than I expected for my age.

- Strongly disagree (1)
- Disagree (2)
- Uncertain (3)
- Agree (4)
- Strongly agree (5)

I keep myself as fit and active as possible by exercising.

- Strongly disagree (1)
- Disagree (2)
- Uncertain (3)
- Agree (4)
- Strongly agree (5)

End of Block: Experiences and attitudes to ageing questionnaire

Start of Block: Self-reported memory

We would like to ask some questions about your memory.

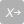

Do you currently have any problems with your memory?

- Yes (1)
- No (0)

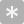

Do you have any comments on your response?

________________________________________________________________

| Page Break |
| --- |

Display This Question:

If Do you currently have any problems with your memory? = Yes

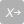

If yes, are these problems interfering with your normal life?

- Yes (1)
- No (0)

Display This Question:

If Do you currently have any problems with your memory? = Yes

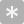

Do you have any comments on your response?

________________________________________________________________

| Page Break |
| --- |

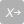

Do you forget where you have left things more than you used to?

- Yes (1)
- No (0)

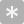

Do you have any comments on your response?

________________________________________________________________

| Page Break |
| --- |

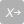

Do you forget the names of close friends or relatives?

- Yes (1)
- No (0)

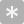

Do you have any comments on your response?

________________________________________________________________

| Page Break |
| --- |

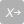

Have you ever been in your own neighbourhood and forgotten your way?

- Yes (1)
- No (0)

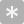

Do you have any comments on your response?

________________________________________________________________

End of Block: Self-reported memory

Start of Block: Final thoughts

The COVID-19 pandemic is an unprecedented global health and societal emergency. We are interested in your experience of the COVID-19 pandemic and thoughts about the future.

If there is anything you would like to mention regarding your experience of the COVID-19 pandemic, please use the box below:

________________________________________________________________

________________________________________________________________

________________________________________________________________

________________________________________________________________

________________________________________________________________

End of Block: Final thoughts
