## Supplementary material for "Impact of COVID-19 lockdown on psychosocial factors, health, and lifestyle in Scottish octogenarians: the Lothian Birth Cohort 1936 Study": R code; Correlation matrix; STROBE: Descriptives code.R

```
# Load packages
library(psych)
library(foreign)
library(gplots)
library(ggplot2)
library(dplyr)
library(lsr)
library(tidyverse)
library(GPArotation)
library(car)
library(GGally)
library(psy)
library(Hmisc)
library(MASS)
library(reshape2)
library(jtools)
library(RColorBrewer)
library(brant)
library(lmtest)
library(ordinal)


# LBC1936 -----------------------------------------------------------------


# Load data
Descriptives <- data.frame (read.spss ("Descriptives.sav", to.data.frame = TRUE))

describe(Descriptives) 
summary(Descriptives)

# Data checking and cleaning ----------------------------------------------

#Attend Wave 5 
class (Descriptives$attendw5)
table(Descriptives$attendw5)
contrasts(Descriptives$attendw5)
summary(Descriptives$attendw5)

#Responeded to covid 19 questionnaire
class (Descriptives$responder)
table(Descriptives$responder)
contrasts(Descriptives$responder)
summary(Descriptives$responder)

# Sex (Male = 0 [level 1], Female = 1 [level 2])
class(Descriptives$sex)
contrasts(Descriptives$sex)
summary(Descriptives$sex)
proportions <- table(Descriptives$sex)/length(Descriptives$sex)
percentages <- proportions*100
proportions
percentages

# Age in days at SMS 1947
class(Descriptives$agedays_SMS)
describe(Descriptives$agedays_SMS)
describe(Descriptives$agedays_SMS/365.25)
hist(Descriptives$agedays_SMS/365.25)
plot(Descriptives$agedays_SMS/365.25)

#Age in years at SMS 1947
Descriptives$age11 <- Descriptives$agedays_SMS/365.25
describe(Descriptives$age11)
hist(Descriptives$age11)
plot(Descriptives$age11)

#Age in days at MHT wave 1
describe(Descriptives$agedays_w1)
hist(Descriptives$agedays_w1)
plot(Descriptives$agedays_w1)

#Age in years at MHT wave 1
Descriptives$age70 <- Descriptives$agedays_w1/365.25
describe(Descriptives$age70)
hist(Descriptives$age70)
plot(Descriptives$age70)

#Age in days at MHT wave 4
describe(Descriptives$agedays_w4)
hist(Descriptives$agedays_w4)
plot(Descriptives$agedays_w4)

#Age in years at MHT wave 4
Descriptives$age79 <- Descriptives$agedays_w4/365.25
describe(Descriptives$age79)
hist(Descriptives$age79)
plot(Descriptives$age79)

# Age 82 age in years
describe(Descriptives$agedays_w5)
Descriptives$age82 <- Descriptives$agedays_w5/365.25
class(Descriptives$age82)
describe(Descriptives$age82)
hist(Descriptives$age82)
plot(Descriptives$age82)

# Age in days when completed COVID-19 questionnaire
describe(Descriptives$agedays_CIQ)
hist(Descriptives$agedays_CIQ)
plot(Descriptives$agedays_CIQ)

#Age in years when completed COVID-19 questionnaire
Descriptives$ageyearsCIQ <- Descriptives$agedays_CIQ/365.25
class(Descriptives$ageyearsCIQ)
describe(Descriptives$ageyearsCIQ)
hist(Descriptives$ageyearsCIQ)
plot(Descriptives$ageyearsCIQ)

# Father's SES (General Register Office's Census 1951 Classification of Occupations. Five categories from 1 (professional) to 5(unskilled))
class(Descriptives$fathclass)
describe(Descriptives$fathclass)
hist(Descriptives$fathclass)
plot(Descriptives$fathclass)

# Participant's SES (Office of Population's Censuses and Survey's Classification of Occupations 1980. Six categories from 1 (professional) to 5(unskilled))
class(Descriptives$pptclass)
Descriptives$pptclass <- Descriptives$hmsonum_w1
which(Descriptives$pptclass == 3.5)
LBC1936$pptclass[Descriptives$pptclass == 3.5] <- 3.0 # Combine sES categories 3 and 3.5 
describe(Descriptives$pptclass)
hist(Descriptives$pptclass)
plot(Descriptives$pptclass)

# Participant's education (years of full-time education)
Descriptives$educ <- Descriptives$yrsedu
class(Descriptives$educ)
describe(Descriptives$educ)
hist(Descriptives$educ)
plot(Descriptives$educ)

# Marital status
class(Descriptives$marital)
summary(Descriptives$marital)
which (Descriptives$marital == "other")
#check comments for 'other' and see if they can be re-categorised
#824: divorced, in a relationship of 10 years with man in England, stays regularly at his home but not cohabiting -- recode divorced
#1050: divorced, was cohabiting with new partner but he died very recently – recode divorced
Descriptives$marital [Descriptives$marital == "other"] <- "divorced"
#combine separated and divorced due to low numbers
Descriptives$marital [Descriptives$marital == "separated"] <- "divorced"
Descriptives$marital <- droplevels (Descriptives$marital)
summary(Descriptives$marital)

Descriptives$binarymarital <- Descriptives$marital
summary(Descriptives$binarymarital)
which (Descriptives$binarymarital == "single")
Descriptives$binarymarital <- as.numeric(Descriptives$binarymarital)
Descriptives$binarymarital[Descriptives$binarymarital == 3] <- 1
Descriptives$binarymarital[Descriptives$binarymarital == 4] <- 1
Descriptives$binarymarital[Descriptives$binarymarital == 5] <- 1

Descriptives$binarymarital <-ordered(Descriptives$binarymarital,
                          levels = c(1, 2),
                          labels = c("not married", "married"))

summary(Descriptives$binarymarital)

# LIVE ALONE (YES alone (0) /NO not alone (1))
Descriptives$alone <- Descriptives$livealn_w5
class(Descriptives$alone)
summary(Descriptives$alone)
contrasts(Descriptives$alone)
levels(Descriptives$alone)
plot(Descriptives$alone)

proportions <- table(Descriptives$alone)/length(Descriptives$alone)
percentages <- proportions*100
proportions
percentages

# RESIDENCE 
Descriptives$resdnc <- Descriptives$resdnc_w5
class(Descriptives$resdnc)
summary(Descriptives$resdnc)
levels(Descriptives$resdnc)

#drop levels of unused categories
Descriptives$resdnc <- droplevels(Descriptives$resdnc)
summary(Descriptives$resdnc)


which(Descriptives$resdnc == "other")
## look up residence comments for 239, 614, 648, 658, 866 and see if any can be put into other categories
#239: property owned by daughter, previously owned by ppt but transferred ownership   --  NA
#614: ‘retirement flat’ -- NA
#648: sheltered housing – residential home
#658: property owned by granddaughter now, previously owned by ppt -- NA
#866: sheltered housing – residential home
Descriptives$resdnc [239] <- NA
Descriptives$resdnc [614] <- NA
Descriptives$resdnc [648] <- "residential home"
Descriptives$resdnc [658] <- NA
Descriptives$resdnc [866] <- "residential home"
Descriptives$resdnc <- droplevels (Descriptives$resdnc)
summary(Descriptives$resdnc)


# MMSE age 82
class(Descriptives$mmse_w5)
describe(Descriptives$mmse_w5)
Descriptives$mmse_w5 [Descriptives$mmse_w5 == -999] <- NA
Descriptives$mmse82 <- Descriptives$mmse_w5
describe(Descriptives$mmse82)
hist(Descriptives$mmse82)
plot(Descriptives$mmse82)

# Age 11 MHT score
Descriptives$mht11 <- Descriptives$mht1947
class(Descriptives$mht11)
describe(Descriptives$mht11)
hist(Descriptives$mht11)
plot(Descriptives$mht11)
which(Descriptives$mht11<10)

# Age adjusted age-79 MHT (standardised residuals from MHT score regressed on age in days in SPSS)
class(Descriptives$adjustedmht11)
describe(Descriptives$adjustedmht11)
hist(Descriptives$adjustedmht11)
plot(Descriptives$adjustedmht11)

# Age 79 MHT score
class(Descriptives$mht_w4)
Descriptives$mht79 <- Descriptives$mht_w4
Descriptives$mht79 [Descriptives$mht79 == 999] <- NA
Descriptives$mht79 [Descriptives$mht79 == -777] <- NA
describe(Descriptives$mht79)
hist(Descriptives$mht79)
plot(Descriptives$mht79)
which(Descriptives$mht79 <20)

# Age adjusted age-79 MHT (standardised residuals from MHT score regressed on age in days in SPSS)
class(Descriptives$adjustedmht79)
describe(Descriptives$adjustedmht79)
hist(Descriptives$adjustedmht79)
plot(Descriptives$adjustedmht79)

#HEALTH LITERACY (general factor derived in SPSS from 3 ranked health literacy totals)
describe(Descriptives$GHL)
plot(Descriptives$GHL)
hist(Descriptives$GHL)

#g (general cognitive ability factor derived in SPSS from digit symbol coding, digit span backwards, letter number sequencing, block design, matrix reasoning, symbol search)
describe(Descriptives$g)
plot(Descriptives$g)
hist(Descriptives$g)

# Self-reported health
class(Descriptives$hlthcat_w5)
Descriptives$srh <- Descriptives$hlthcat_w5
summary(Descriptives$srh)
plot(Descriptives$srh)

# Hypertension
class(Descriptives$hibp_w5)
Descriptives$hibp <- Descriptives$hibp_w5
summary(Descriptives$hibp)
which(Descriptives$hibp == "Unsure / Ambiguous")
Descriptives$hibp[230] <- "Yes"
Descriptives$hibp[463]<- NA
Descriptives$hibp[466]<- NA
Descriptives$hibp[601]<- NA
Descriptives$hibp[766]<- NA
Descriptives$hibp[867]<- NA
Descriptives$hibp [Descriptives$hibp == -888] <- NA
Descriptives$hibp <- droplevels(Descriptives$hibp)
summary(Descriptives$hibp)
# notes checked and all except 230 unclear. 230 recoded 'yes' and others recoded 'NA'

# CVD
class(Descriptives$cvdhist_w5)
summary(Descriptives$cvdhist_w5)
which(Descriptives$cvdhist_w5 == "Unsure / Ambiguous")
Descriptives$cvdhistcom_w5 [556]
Descriptives$cvdhist_w5 [746]
Descriptives$cvdhist_w5[556] <- NA
Descriptives$cvdhist_w5[746] <- NA
Descriptives$cvdhist_w5<- droplevels (Descriptives$cvdhist_w5)
Descriptives$cvd<-Descriptives$cvdhist_w5
summary(Descriptives$cvd)
plot(Descriptives$cvd)

# Number of chronic diseases
describe(Descriptives$disease)
plot(Descriptives$disease)

# Diabetes
class(Descriptives$diab_w5)
Descriptives$diab <- Descriptives$diab_w5
summary(Descriptives$diab)
Descriptives$diab <- droplevels(Descriptives$diab)
plot(Descriptives$diab)

# BMI
# height (cm)
class(Descriptives$height_w5)
describe(Descriptives$height_w5)
Descriptives$height_w5 [Descriptives$height_w5 == -999] <- NA
plot(Descriptives$height_w5)
hist(Descriptives$height_w5)
# weight (kg)
class(Descriptives$weight_w5)
describe(Descriptives$weight_w5)
Descriptives$weight_w5 [Descriptives$weight_w5 == -999] <- NA
plot(Descriptives$weight_w5)
hist(Descriptives$weight_w5)
#BMI (weight divided by height squared (kg/m2))
Descriptives$height_met <- Descriptives$height_w5/100
Descriptives$height_metsq <- Descriptives$height_met^2
Descriptives$bmi <- Descriptives$weight_w5/Descriptives$height_metsq
describe(Descriptives$bmi)
which(Descriptives$bmi <17)

# Six meter walk (in seconds)
class(Descriptives$sixmwk_w5)
Descriptives$sixm <- Descriptives$sixmwk_w5
describe(Descriptives$sixm)
hist(Descriptives$sixm)
plot(Descriptives$sixm)
which (Descriptives$sixm <3)

# GRIP STRENGTH
#Recorded as max of 3 measurements in dominant hand.
class(Descriptives$griprh_1_w5)
class(Descriptives$griprh_2_w5)
class(Descriptives$griprh_3_w5)
class(Descriptives$griplh_1_w5)
class(Descriptives$griplh_2_w5)
class(Descriptives$griplh_3_w5)

Descriptives$grip <- pmax(Descriptives$griplh_1_w5, Descriptives$griplh_2_w5, Descriptives$griplh_3_w5, Descriptives$griprh_1_w5, Descriptives$griprh_2_w5, Descriptives$griprh_3_w5)

describe(Descriptives$grip)
head(Descriptives$grip)
head(Descriptives$griprh_1_w5)
head(Descriptives$griprh_2_w5)
head(Descriptives$griprh_3_w5)
head(Descriptives$griplh_1_w5)
head(Descriptives$griplh_2_w5)
head(Descriptives$griplh_3_w5)
plot(Descriptives$grip)
hist(Descriptives$grip)

#LUNG FUNCTION (FEV1)
#recorded as max of 3 values
class(Descriptives$fev_1_w5)
class(Descriptives$fev_2_w5)
class(Descriptives$fev_3_w5)
Descriptives$lung <- pmax (Descriptives$fev_1_w5, Descriptives$fev_2_w5, Descriptives$fev_3_w5)
describe(Descriptives$lung)
head(Descriptives$lung)
head(Descriptives$fev_1_w5)
head(Descriptives$fev_2_w5)
head(Descriptives$fev_3_w5)
plot(Descriptives$lung)
hist(Descriptives$lung)

# Townsend Disability Scale

Descriptives$townsend <- Descriptives$adl_w5
describe(Descriptives$townsend)
plot(Descriptives$townsend)
hist(Descriptives$townsend)

# Hba1c 
class(Descriptives$bld_hba1c_IFFC_w5)
Descriptives$hba1c <- Descriptives$bld_hba1c_IFFC_w5
describe(Descriptives$hba1c)
plot(Descriptives$hba1c)
hist(Descriptives$hba1c)

# HADS anxiety
class (Descriptives$HADS_A_w5)
Descriptives$HADSA <- Descriptives$HADS_A_w5
describe (Descriptives$HADSA)
plot (Descriptives$HADSA)
hist(Descriptives$HADSA)

# HADS depression
class (Descriptives$HADS_D_w5)
Descriptives$HADSD <- Descriptives$HADS_D_w5
describe (Descriptives$HADSD)
plot (Descriptives$HADSD)
hist(Descriptives$HADSD)

#IPIP Emotional Stability (using recalculated ES variable which had 10 added to it to put on proper IPIP scoring scale with max score 50 in SPSS)
class (Descriptives$ES)
Descriptives$emo <- Descriptives$ES
describe (Descriptives$emo)
Descriptives$emo [Descriptives$emo == -54] <- NA
plot (Descriptives$emo)
hist(Descriptives$emo)

#IPIP Extraversion  (using recalculated EX variable which had 10 added to it to put on proper IPIP scoring scale with max score 50 in SPSS)
class (Descriptives$EX)
Descriptives$ex <- Descriptives$EX
describe (Descriptives$ex)
Descriptives$ex [Descriptives$ex == 111] <- NA
plot (Descriptives$ex)
hist(Descriptives$ex)

#IPIP Conscientiousness (using recalculated CON variable which had 10 added to it to put on proper IPIP scoring scale with max score 50 in SPSS)
class (Descriptives$CON)
Descriptives$con <- Descriptives$CON
describe (Descriptives$con)
plot (Descriptives$con)
hist(Descriptives$con)


# Questionnaire Cleaning --------------------------------------------------

#CIQ_ThinksCovid & CIQ_WhyThinksCovid 
class(Descriptives$CIQ_ThinksCovid)     ##Whether ppt had covid (test or suspected)
summary(Descriptives$CIQ_ThinksCovid)

class(Descriptives$CIQ_WhyThinksCovidNoTest)    #why ppt suspected, if suspected
summary(Descriptives$CIQ_WhyThinksCovidNoTest)


#CIQ_Infected variables 
class(Descriptives$CIQ_Infected_Shielded)
summary(Descriptives$CIQ_Infected_Shielded)
class(Descriptives$CIQ_Infected_IsolatedDistanced)
summary(Descriptives$CIQ_Infected_IsolatedDistanced) 
class(Descriptives$CIQ_Infected_NHS111_GP)
summary(Descriptives$CIQ_Infected_NHS111_GP) 
class(Descriptives$CIQ_Infected_Swabbed)
summary(Descriptives$CIQ_Infected_Swabbed) 
class(Descriptives$CIQ_Infected_HubDoctor)
summary(Descriptives$CIQ_Infected_HubDoctor) 
class(Descriptives$CIQ_Infected_AdmittedHospital)
summary(Descriptives$CIQ_Infected_AdmittedHospital) 

class(Descriptives$CIQ_Infected_ICU)
summary(Descriptives$CIQ_Infected_ICU) #none admitted to hosp - therefore all NA
class(Descriptives$CIQ_Infected_Ventilator)
summary(Descriptives$CIQ_Infected_Ventilator) #all NA

#CIQ_Infected_OtherInHousehold
class(Descriptives$CIQ_Infected_OtherInHousehold)
summary(Descriptives$CIQ_Infected_OtherInHousehold) #NONE
#CIQ_Infected_OtherHowDiagnosed no household cases so all NA

#CIQ_Knowledge
class(Descriptives$CIQ_Knowledge) 
Descriptives$CIQ_Knowledge <- factor(Descriptives$CIQ_Knowledge, order=T, 
                                     levels = c('Extremely good',
                                                'Somewhat good',
                                                'Neither good nor bad',
                                                'Somewhat bad',
                                                'Extremely bad'))

summary(Descriptives$CIQ_Knowledge)
plot(Descriptives$CIQ_Knowledge) #skewed
Descriptives$knowledge <- Descriptives$CIQ_Knowledge
Descriptives$knowledge <- droplevels(Descriptives$knowledge)
summary(Descriptives$knowledge)


#CIQ_GovtGuidance
class(Descriptives$CIQ_GovtGuidance) 
levels(Descriptives$CIQ_GovtGuidance)
Descriptives$CIQ_GovtGuidance <- factor(Descriptives$CIQ_GovtGuidance, order=T, 
                                        levels = c("Extremely easy", 
                                                   "Somewhat easy", 
                                                   "Neither easy nor difficult", 
                                                   "Somewhat difficult", 
                                                   "Extremely difficult", 
                                                   "I am not aware of the government guidance on COVID-19"))
summary(Descriptives$CIQ_GovtGuidance)
plot(Descriptives$CIQ_GovtGuidance)

Descriptives$guidance <- Descriptives$CIQ_GovtGuidance
Descriptives$guidance <- droplevels(Descriptives$guidance)
summary(Descriptives$guidance)

Descriptives$numguidance <- as.numeric(Descriptives$CIQ_GovtGuidance)

#CIQ_SocialDistance
class(Descriptives$CIQ_SocialDistance) 
levels(Descriptives$CIQ_SocialDistance)
Descriptives$CIQ_SocialDistance[Descriptives$CIQ_SocialDistance == 'N/A'] <- NA
Descriptives$CIQ_SocialDistance <- factor(Descriptives$CIQ_SocialDistance, order=T, 
                                          levels = c("Always", 
                                                     "Most of the time", 
                                                     "Some of the time", 
                                                     "Never") )
summary(Descriptives$CIQ_SocialDistance)
plot(Descriptives$CIQ_SocialDistance) 

#CIQ_StayHome
class(Descriptives$CIQ_StayHome) 
levels(Descriptives$CIQ_StayHome)
Descriptives$CIQ_StayHome[Descriptives$CIQ_StayHome == 'N/A'] <- NA
Descriptives$CIQ_StayHome <- factor(Descriptives$CIQ_StayHome, order=T, 
                                    levels = c("Always", 
                                               "Most of the time", 
                                               "Some of the time", 
                                               "Never") )
summary(Descriptives$CIQ_StayHome)
plot(Descriptives$CIQ_StayHome) 

#CIQ_SelfIsolating
class(Descriptives$CIQ_SelfIsolating) 
levels(Descriptives$CIQ_SelfIsolating)
Descriptives$CIQ_SelfIsolating[Descriptives$CIQ_SelfIsolating == 'N/A'] <- NA
Descriptives$CIQ_SelfIsolating <- factor(Descriptives$CIQ_SelfIsolating, order=T, 
                                         levels = c("Always", 
                                                    "Most of the time", 
                                                    "Some of the time", 
                                                    "Never") )
summary(Descriptives$CIQ_SelfIsolating)
plot(Descriptives$CIQ_SelfIsolating) 

#CIQ_HandWash
class(Descriptives$CIQ_HandWash) 
levels(Descriptives$CIQ_HandWash)
Descriptives$CIQ_HandWash[Descriptives$CIQ_HandWash == 'N/A'] <- NA
Descriptives$CIQ_HandWash <- factor(Descriptives$CIQ_HandWash, order=T, 
                                    levels = c("Always", 
                                               "Most of the time", 
                                               "Some of the time", 
                                               "Never") )
summary(Descriptives$CIQ_HandWash)
plot(Descriptives$CIQ_HandWash) 

# Adhere to guidance variable
# PCA of 4 adherence variables (stay home, self-isolate, wash hands, social distancing) conducted in SPSS and scores saved.
class(Descriptives$adherence)
describe(Descriptives$adherence)
plot(Descriptives$adherence)
hist(Descriptives$adherence)

#CIQ_StaysUpdated
class(Descriptives$CIQ_StaysUpdated) 
levels(Descriptives$CIQ_StaysUpdated)
Descriptives$CIQ_StaysUpdated <- factor(Descriptives$CIQ_StaysUpdated, order=T, 
                                        levels = c("Multiple times per day",
                                                   "Daily",
                                                   "Every few days",
                                                   "Weekly",
                                                   "Less than weekly"  ) )
summary(Descriptives$CIQ_StaysUpdated)
plot(Descriptives$CIQ_StaysUpdated) 

#CIQ_Source variables
summary(Descriptives[ ,160:176])
summary(Descriptives[ ,176:193])

#CIQ_SeriousRiskWarning
class(Descriptives$CIQ_SeriousRiskWarning)
summary(Descriptives$CIQ_SeriousRiskWarning)
contrasts(Descriptives$CIQ_SeriousRiskWarning)

#CIQ_AvoidedMedicalCare (avoid or postpone contacting medical services)
class(Descriptives$CIQ_AvoidedMedicalCare)
summary(Descriptives$CIQ_AvoidedMedicalCare)
Descriptives$CIQ_AvoidedMedicalCare[Descriptives$CIQ_AvoidedMedicalCare == 'Not applicable'] <- NA
droplevels(Descriptives$CIQ_AvoidedMedicalCare) -> Descriptives$CIQ_AvoidedMedicalCare
plot(Descriptives$CIQ_AvoidedMedicalCare)

Descriptives$avoid <- Descriptives$CIQ_AvoidedMedicalCare
table(Descriptives$avoid)

#CIQ_LivesWith variables (number & identify who)
class(Descriptives$CIQ_LivesWith_HowMany)
summary(Descriptives$CIQ_LivesWith_HowMany)
Descriptives$covidalone <- Descriptives$CIQ_LivesWith_HowMany
class(Descriptives$covidalone)
Descriptives$covidalone[Descriptives$covidalone == 3] <- 2
Descriptives$covidalone[Descriptives$covidalone == 4] <- 2
Descriptives$covidalone[Descriptives$covidalone == 5] <- 2
Descriptives$covidalone[Descriptives$covidalone == 6] <- 2
Descriptives$covidalone[Descriptives$covidalone == 7] <- 2
Descriptives$covidalone[Descriptives$covidalone == 8] <- 2
Descriptives$covidalone[Descriptives$covidalone == 9] <- 2
Descriptives$covidalone[Descriptives$covidalone == "10 or more"] <- 2
summary(Descriptives$covidalone)
Descriptives$covidalone <- droplevels(Descriptives$covidalone)
Descriptives$covidalone <-ordered(Descriptives$covidalone,
                                     levels = c(1, 2),
                                     labels = c("covidalone", "covidnotalone"))
summary(Descriptives$covidalone)

Descriptives$CIQ_LivesWith_HowMany <- as.numeric(Descriptives$CIQ_LivesWith_HowMany)
describe(Descriptives$CIQ_LivesWith_HowMany)
plot(Descriptives$CIQ_LivesWith_HowMany)
summary(Descriptives[ ,197:204]) #who lives with

#CIQ_LivSituChange_Check
class(Descriptives$CIQ_LivSituChange_Check)
summary(Descriptives$CIQ_LivSituChange_Check)
contrasts(Descriptives$CIQ_LivSituChange_Check)
summary(Descriptives[206:208]) #how situation changed

#CIQ_ResidenceAreaType
class(Descriptives$CIQ_ResidenceAreaType)
summary(Descriptives$CIQ_ResidenceAreaType)
plot(Descriptives$CIQ_ResidenceAreaType)
Descriptives$geog <-  Descriptives$CIQ_ResidenceAreaType

#CIQ_SinceMarch23_TimesLeaveHouse
class(Descriptives$CIQ_SinceMarch23_TimesLeaveHouse)
summary(Descriptives$CIQ_SinceMarch23_TimesLeaveHouse)
levels(Descriptives$CIQ_SinceMarch23_TimesLeaveHouse)
Descriptives$CIQ_SinceMarch23_TimesLeaveHouse <- factor(Descriptives$CIQ_SinceMarch23_TimesLeaveHouse,
                                                        order=T, 
                                                        levels = c("Never, since COVID-19 measures were introduced.",
                                                                   "Less than once per week (e.g. once per fortnight)",
                                                                   "Once per week",
                                                                   "A few times per week",
                                                                   "Once per day",
                                                                   "Multiple times a day"))
plot(Descriptives$CIQ_SinceMarch23_TimesLeaveHouse)

Descriptives$leavehouse <- Descriptives$CIQ_SinceMarch23_TimesLeaveHouse
summary(Descriptives$leavehouse)


class(Descriptives$CIQ_LikelyProximity)
summary(Descriptives$CIQ_LikelyProximity)
levels(Descriptives$CIQ_LikelyProximity)
Descriptives$CIQ_LikelyProximity <- factor(Descriptives$CIQ_LikelyProximity, order=T, 
                                           levels = c("I don't leave my home",
                                                      "Not at all likely",
                                                      "Not that likely", 
                                                      "Somewhat likely", 
                                                      "Very likely"  ))
plot(Descriptives$CIQ_LikelyProximity)

#CIQ_Carer variables
class(Descriptives$CIQ_CarerGChildren)
contrasts(Descriptives$CIQ_CarerGChildren)
summary(Descriptives$CIQ_CarerGChildren)
summary(Descriptives[240:243]) #current carer status (CIQ_CarerGChildren;CIQ_CarerLiveWith;CIQ_CarerNotLivesWith;CIQ_NotCarer            )
summary(Descriptives[244:246]) #previous carer status (CIQ_CarerLivesWithBeforeMarch23;CIQ_CarerNotLivesWithBeforeMarch23;CIQ_NotCarerBeforeMarch23)
summary(Descriptives$CIQ_CarerExpChangeSinceMarch23)

#CIQ_Prescriptions variables
summary(Descriptives$CIQ_Prescriptions_NoChange)
summary(Descriptives$CIQ_Prescriptions_SwitchToDelivered)
summary(Descriptives$CIQ_Prescriptions_IncreasedSupply)
summary(Descriptives$CIQ_Prescriptions_Other)
summary(Descriptives$CIQ_Prescriptions_NA) 

summary(Descriptives$CIQ_Prescriptions_OtherDetails)

#CIQ_GetsHelp
class(Descriptives$CIQ_GetsHelp)
summary(Descriptives$CIQ_GetsHelp)
contrasts(Descriptives$CIQ_GetsHelp)
summary(Descriptives[278:280]) #gets help groceries : help medicines : other
summary(Descriptives$CIQ_GetsHelpOtherDetails)

Descriptives$help <- Descriptives$CIQ_GetsHelp
summary(Descriptives$help)

#CIQ_HelpsOthers
summary(Descriptives$CIQ_HelpsOthers)
class(Descriptives$CIQ_HelpsOthers)
contrasts(Descriptives$CIQ_HelpsOthers)

#CIQ_Cash variables 
summary(Descriptives$CIQ_Cash_ChangeToCards)
contrasts(Descriptives$CIQ_Cash_ChangeToCards)
summary(Descriptives$CIQ_Cash_CashImportance)
levels(Descriptives$CIQ_Cash_CashImportance)
Descriptives$CIQ_Cash_CashImportance <- factor(Descriptives$CIQ_Cash_CashImportance, order=T, 
                                               levels = c("Very important",
                                                          "Quite important",
                                                          "Not very important",
                                                          "Not important at all"))
class(Descriptives$CIQ_Cash_CashImportance)

#CIQ_PriorFreq & CIQ_NowFreq variables (change in freq of contact with family/friends)
summary(Descriptives$CIQ_PriorFreq_MeetFamily)
summary(Descriptives$CIQ_PriorFreq_MeetFriends)
summary(Descriptives$CIQ_PriorFreq_CallFamily)
summary(Descriptives$CIQ_PriorFreq_CallFriends)
summary(Descriptives$CIQ_PriorFreq_VidCall_Family)
summary(Descriptives$CIQ_PriorFreq_VidCall_Friends)
summary(Descriptives$CIQ_PriorFreq_MsgFamily)
summary(Descriptives$CIQ_PriorFreq_MsgFriends)
summary(Descriptives$CIQ_NowFreq_MeetFamily)
summary(Descriptives$CIQ_NowFreq_MeetFriends)
summary(Descriptives$CIQ_NowFreq_CallFamily)
summary(Descriptives$CIQ_NowFreq_CallFriends)
summary(Descriptives$CIQ_NowFreq_VidCall_Family)
summary(Descriptives$CIQ_NowFreq_VidCall_Friends)
summary(Descriptives$CIQ_NowFreq_MsgFamily)
summary(Descriptives$CIQ_NowFreq_MsgFriends)  

#CIQ_NeighbourContact
class(Descriptives$CIQ_NeighbourContactMoreLess)
summary(Descriptives$CIQ_NeighbourContactMoreLess)
class(Descriptives$CIQ_NeighbourContactPosNeg)
summary(Descriptives$CIQ_NeighbourContactPosNeg)

#CIQ_LocalInitiatives
class(Descriptives$CIQ_LocalInitiatives)
summary(Descriptives$CIQ_LocalInitiatives)
summary(Descriptives$CIQ_LocalInitiativesDetails)

#CIQ_MentalHealth
class(Descriptives$CIQ_BeforeMarch23_MentalHealth)
summary(Descriptives$CIQ_BeforeMarch23_MentalHealth)
Descriptives$CIQ_BeforeMarch23_MentalHealth <- factor(Descriptives$CIQ_BeforeMarch23_MentalHealth, ordered=T, 
                                                      levels= c("Excellent", 
                                                                "Very good", 
                                                                "Good", 
                                                                "Fair", 
                                                                "Poor"))
plot(Descriptives$CIQ_BeforeMarch23_MentalHealth)
Descriptives$CIQ_SinceMarch23_MentaHealth -> Descriptives$CIQ_SinceMarch23_MentalHealth
class(Descriptives$CIQ_SinceMarch23_MentalHealth)
summary(Descriptives$CIQ_SinceMarch23_MentalHealth)
Descriptives$CIQ_SinceMarch23_MentalHealth <- factor(Descriptives$CIQ_SinceMarch23_MentalHealth, ordered=T, 
                                                     levels= c("Excellent", 
                                                               "Very good", 
                                                               "Good", 
                                                               "Fair", 
                                                               "Poor"))
plot(Descriptives$CIQ_SinceMarch23_MentalHealth)

Descriptives$mentalhealth <-Descriptives$CIQ_SinceMarch23_MentalHealth
summary(Descriptives$mentalhealth)

#CIQ_LastTwoWeeks_Stress
class(Descriptives$CIQ_LastTwoWeeks_Stress)
summary(Descriptives$CIQ_LastTwoWeeks_Stress)
Descriptives$CIQ_LastTwoWeeks_Stress <- factor(Descriptives$CIQ_LastTwoWeeks_Stress, ordered=T, 
                                               levels= c("Never",
                                                         "Some of the time",
                                                         "Most of the time",
                                                         "All of the time"))
plot(Descriptives$CIQ_LastTwoWeeks_Stress)

Descriptives$covidstress <- Descriptives$CIQ_LastTwoWeeks_Stress
Descriptives$covidstress <- droplevels(Descriptives$covidstress)
summary(Descriptives$covidstress)

#CIQ_PhysicalHealth 
class(Descriptives$CIQ_BeforeMarch23_PhysicalHealth)
summary(Descriptives$CIQ_BeforeMarch23_PhysicalHealth)
Descriptives$CIQ_BeforeMarch23_PhysicalHealth <- factor(Descriptives$CIQ_BeforeMarch23_PhysicalHealth, ordered=T, 
                                                        levels= c("Excellent", 
                                                                  "Very good", 
                                                                  "Good", 
                                                                  "Fair",
                                                                  "Poor"))
plot(Descriptives$CIQ_BeforeMarch23_PhysicalHealth)
class(Descriptives$CIQ_SinceMarch23_PhysicalHealth)
summary(Descriptives$CIQ_SinceMarch23_PhysicalHealth)
Descriptives$CIQ_SinceMarch23_PhysicalHealth <- factor(Descriptives$CIQ_SinceMarch23_PhysicalHealth, ordered=T, 
                                                       levels= c("Excellent", 
                                                                 "Very good", 
                                                                 "Good", 
                                                                 "Fair",
                                                                 "Poor"))

plot(Descriptives$CIQ_SinceMarch23_PhysicalHealth)

Descriptives$physicalhealth <- Descriptives$CIQ_SinceMarch23_PhysicalHealth
Descriptives$physicalhealth <- droplevels(Descriptives$physicalhealth)
summary(Descriptives$physicalhealth)
    
     #CIQ_Alc
     class(Descriptives$CIQ_Alc_DrinksAlc)
     summary(Descriptives$CIQ_Alc_DrinksAlc)
     class(Descriptives$CIQ_Alc_Change)
     summary(Descriptives$CIQ_Alc_Change)
     Descriptives$CIQ_Alc_Change <- factor(Descriptives$CIQ_Alc_Change, ordered=T, 
                                           levels= c("Drinking more alcohol now", 
                                                     "Drinking about the same amount of alcohol now",
                                                     "Drinking less alcohol now"))
     
Descriptives$alcohol <- Descriptives$CIQ_Alc_Change
summary(Descriptives$alcohol)
plot(Descriptives$alcohol)  

     #CIQ_Smoker   
     class(Descriptives$CIQ_Smok_Smoker)
     summary(Descriptives$CIQ_Smok_Smoker)
     class(Descriptives$CIQ_Smok_Change)
     summary(Descriptives$CIQ_Smok_Change)
     Descriptives$CIQ_Smok_Change <- factor(Descriptives$CIQ_Smok_Change, ordered=T, 
                                            levels= c("Smoking more now", 
                                                      "Smoking about the same now",
                                                      "Smoking less now" ))

Descriptives$smoking <- Descriptives$CIQ_Smok_Change   
summary(Descriptives$smoking)
plot(Descriptives$smoking) 

     #CIQ_Diet
     class(Descriptives$CIQ_Diet_ChangeHeathly)
     summary(Descriptives$CIQ_Diet_ChangeHeathly)
     Descriptives$CIQ_Diet_ChangeHeathly <- factor(Descriptives$CIQ_Diet_ChangeHeathly, ordered=T, 
                                                   levels= c("Much healthier now",
                                                             "Slightly healthier now",
                                                             "About the same now",
                                                             "Slightly less healthy now",
                                                             "Much less healthy now"))
     plot(Descriptives$CIQ_Diet_ChangeHeathly)
     
Descriptives$healthydiet <- Descriptives$CIQ_Diet_ChangeHeathly
summary(Descriptives$healthydiet)
     
     class(Descriptives$CIQ_Diet_ChangeIntake)
     summary(Descriptives$CIQ_Diet_ChangeIntake)
     Descriptives$CIQ_Diet_ChangeIntake <- factor(Descriptives$CIQ_Diet_ChangeIntake, ordered=T, 
                                                  levels= c("More now",
                                                            "About the same",
                                                            "Less now"))
     plot(Descriptives$CIQ_Diet_ChangeIntake)
     
     
Descriptives$dietintake <- Descriptives$CIQ_Diet_ChangeIntake 
summary(Descriptives$dietintake)


     #CIQ_PhysActivChange
     class(Descriptives$CIQ_PhysActivChange)
     summary(Descriptives$CIQ_PhysActivChange)
     Descriptives$physactivity <- Descriptives$CIQ_PhysActivChange
     plot(Descriptives$physactivity)
     
     #CIQ_DailyRoutineChange
     class(Descriptives$CIQ_DailyRoutineChange)
     summary(Descriptives$CIQ_DailyRoutineChange)
     Descriptives$CIQ_DailyRoutineChange <- factor(Descriptives$CIQ_DailyRoutineChange, ordered=T, 
                                                   levels= c("A lot",
                                                             "Somewhat",   
                                                             "A little",
                                                             "Not at all"))
     plot(Descriptives$CIQ_DailyRoutineChange)
     
     #CIQ_Pastime
     summary(Descriptives[364:383])
     summary(Descriptives$CIQ_Pastime_OtherDetails)
     
Descriptives$pasttime <- as.factor(Descriptives$pasttime)
summary(Descriptives$pasttime)
Descriptives$pasttime <-ordered(Descriptives$pasttime,
                                     levels = c(0, 1),
                                     labels = c("new hobby", "no new hobby"))
summary(Descriptives$pasttime)
     
     #CIQ_GdnAllot
     class(Descriptives$CIQ_GdnAllot_Garden)
     summary(Descriptives$CIQ_GdnAllot_Garden)
     class(Descriptives$CIQ_GdnAllot_Allotment)
     summary(Descriptives$CIQ_GdnAllot_Allotment)
     class(Descriptives$CIQ_GdnAllot_NoAccess)
     summary(Descriptives$CIQ_GdnAllot_NoAccess)
     summary(Descriptives$CIQ_GdnAllot_UsageChange)
     Descriptives$CIQ_GdnAllot_UsageChange <- factor(Descriptives$CIQ_GdnAllot_UsageChange, ordered=T,
                                                     levels= c("Much more often now",
                                                               "Slightly more often now", 
                                                               "About the same now", 
                                                               "Slightly less now", 
                                                               "Much less now"))
     plot(Descriptives$CIQ_GdnAllot_UsageChange)
     
     #CIQ_InternetUse
     class(Descriptives$CIQ_InternetUseChange)
     summary(Descriptives$CIQ_InternetUseChange)
     Descriptives$CIQ_InternetUseChange[Descriptives$CIQ_InternetUseChange=="Not applicable"] <- NA
     Descriptives$CIQ_InternetUseChange <- droplevels(Descriptives$CIQ_InternetUseChange)
     Descriptives$CIQ_InternetUseChange <- factor(Descriptives$CIQ_InternetUseChange, ordered=T,
                                                  levels= c("I use the internet much more now",
                                                            "I use the internet slightly more now",
                                                            "My internet usage is about the same",
                                                            "I use the internet slightly less now",
                                                            "I use the internet much less now"))
     plot(Descriptives$CIQ_InternetUseChange)
    
Descriptives$internet <- Descriptives$CIQ_InternetUseChange
summary(Descriptives$internet)
plot(Descriptives$internet)
     
class(Descriptives$CIQ_InternetUseMoreInFuture)
summary(Descriptives$CIQ_InternetUseMoreInFuture)
     
     #CIQ_loneliness
     class(Descriptives$CIQ_lonelines)
     summary(Descriptives$CIQ_lonelines)
     Descriptives$CIQ_lonelines <- factor(Descriptives$CIQ_lonelines, ordered=T,
                                          levels= c("None or almost none of the time",
                                                    "Some of the time",
                                                    "Most of the time",
                                                    "All or almost all of the time"))
     plot(Descriptives$CIQ_lonelines)

     
# Creating variables for Regressions -------------------------------------------------------------
     
     # Recode Outcome variables into suitable format for regressions
     
     # Binary logistic regression variables ------------------------------------
     
     
     # How has your internet usage changed since COVID-19 measures were introduced
     # Recode as internet used has stayed the same vs use internet more
     
     summary(Descriptives$CIQ_InternetUseChange)
     plot(Descriptives$CIQ_InternetUseChange)
     Descriptives$binaryinternet <- as.numeric(Descriptives$CIQ_InternetUseChange)
     describe(Descriptives$binaryinternet)
     Descriptives$binaryinternet [Descriptives$binaryinternet == 2] <- 1
     Descriptives$binaryinternet [Descriptives$binaryinternet == 4] <- NA
     Descriptives$binaryinternet [Descriptives$binaryinternet == 5] <- NA
     Descriptives$binaryinternet [Descriptives$binaryinternet == 3] <- 2
     
     Descriptives$binaryinternet <- ordered(Descriptives$binaryinternet,
                                            levels = c(1, 2),
                                            labels = c("more internet", "same internet"))
     summary(Descriptives$binaryinternet)
     plot(Descriptives$binaryinternet)
     
     ##In the last two weeks, how often have you felt nervous or stressed because of COVID
     ## Recode as never nervous vs sometimes nervous
     summary(Descriptives$CIQ_LastTwoWeeks_Stress)
     plot(Descriptives$CIQ_LastTwoWeeks_Stress)
     Descriptives$CIQ_LastTwoWeeks_Stress <- droplevels(Descriptives$CIQ_LastTwoWeeks_Stress)
     Descriptives$binarynervous <- as.numeric(Descriptives$CIQ_LastTwoWeeks_Stress)
     describe(Descriptives$binarynervous)
     Descriptives$binarynervous [Descriptives$binarynervous == 3] <- 2
     describe(Descriptives$binarynervous)
     Descriptives$binarynervous <- ordered(Descriptives$binarynervous,
                                           levels = c(1, 2),
                                           labels = c("never nervous", "sometimes nervous"))
     summary(Descriptives$binarynervous)
     plot(Descriptives$binarynervous)
     
#Have you received any additional help in your daily life with things such as grocery shopping, errands, or picking up medications since COVID-19 measures were introduced?
     
     #Already binary, just relabel
     summary(Descriptives$CIQ_GetsHelp)
     plot(Descriptives$CIQ_GetsHelp)
     Descriptives$getshelp <- Descriptives$CIQ_GetsHelp
     summary(Descriptives$getshelp)
     plot(Descriptives$getshelp)
     
 ##Since COVID-19 measures have been in place (23rd March 2020), have you returned to or started up a new pastime that you can do from home? 
     ## already coded as binary in SPSS 
     ## new hobby vs no new hobby
     summary(Descriptives$pasttime)
     plot(Descriptives$pasttime)
     
     ## How often have you felt lonely during the past week? 
     ## recoded to never versus sometimes
     
     summary(Descriptives$CIQ_lonelines)
     plot(Descriptives$CIQ_lonelines)
     Descriptives$lonely <- as.numeric(Descriptives$CIQ_lonelines)
     describe(Descriptives$lonely)
     Descriptives$lonely [Descriptives$lonely == 3] <- 2
     Descriptives$lonely [Descriptives$lonely == 4] <- 2
     Descriptives$lonely <-ordered(Descriptives$lonely,
                                   levels = c(1, 2),
                                   labels = c("none of the time", "some of the time"))
     summary(Descriptives$lonely)
     plot(Descriptives$lonely)
     
     
     
     ##
 # Ordinal logistic regression variables -----------------------------------
     
##Compared to before COVID-19 measures were introduced (23rd March 2020), how much physical activity are you doing now? This includes activities that make you breathe harder than normal (e.g., brisk walking)
# already coded, just relabelled
     
     summary(Descriptives$CIQ_PhysActivChange)
     Descriptives$changephys <- Descriptives$CIQ_PhysActivChange
     summary(Descriptives$changephys)
     plot(Descriptives$changephys)
          
     #How much has COVID-19 changed your daily routine?
     #recode as a lot, somewhat, a little or not at all - combined a little and not at all
     
     summary(Descriptives$CIQ_DailyRoutineChange)
     Descriptives$routine <- Descriptives$CIQ_DailyRoutineChange
     Descriptives$routine <- as.numeric(Descriptives$routine)
     Descriptives$routine [Descriptives$routine == 4] <- 3
     Descriptives$routine <-ordered(Descriptives$routine,
                                   levels = c(1, 2, 3),
                                   labels = c("A lot", "Somewhat", "A little or not at all"))
     summary(Descriptives$routine)
     plot(Descriptives$routine)
     
#How often have you been leaving your home since COVID-19 measures were introduced (23rd March 2020)?
#already coded, just relabelled
     
     summary(Descriptives$CIQ_SinceMarch23_TimesLeaveHouse)
     Descriptives$leavehouse <- Descriptives$CIQ_SinceMarch23_TimesLeaveHouse
     summary(Descriptives$leavehouse)
     plot(Descriptives$leavehouse)
     
     
# Wilcoxon repeated measures variables ------------------------------------
     
#In general, before the COVID-19 measures were introduced (23rd March 2020), would you say your emotional and mental health was:
     
#In general, after the COVID-19 measures were introduced (23rd March 2020), would you say your emotional and mental health was:
     #already coded, just relabelled
     
     summary(Descriptives$CIQ_BeforeMarch23_MentalHealth)
     Descriptives$mentalhealthbefore <- Descriptives$CIQ_BeforeMarch23_MentalHealth
     summary(Descriptives$mentalhealthbefore)
     plot(Descriptives$mentalhealthbefore)
     
#In general, before the COVID-19 measures were introduced (23rd March 2020), would you say your physical health was:
     
     summary(Descriptives$CIQ_SinceMarch23_MentalHealth)
     Descriptives$mentalhealthafter <- Descriptives$CIQ_SinceMarch23_MentalHealth
     summary(Descriptives$mentalhealthafter)
     plot(Descriptives$mentalhealthafter)
     
#In general, before the COVID-19 measures were introduced (23rd March 2020), would you say your physical health was:
     
     summary(Descriptives$CIQ_BeforeMarch23_PhysicalHealth)
     Descriptives$physhealthbefore <- Descriptives$CIQ_BeforeMarch23_PhysicalHealth
     summary(Descriptives$physhealthbefore)
     plot(Descriptives$physhealthbefore)
     
#In general, after the COVID-19 measures were introduced (23rd March 2020), would you say your physical health was:
     
     summary(Descriptives$CIQ_SinceMarch23_PhysicalHealth)
     Descriptives$physhealthafter <- Descriptives$CIQ_SinceMarch23_PhysicalHealth
     summary(Descriptives$physhealthafter)
     plot(Descriptives$physhealthafter)
    
     

# Save dataframe ----------------------------------------------------------
    
# Save Descriptives data frame
     
save(Descriptives,file="Descriptives.Rda")     

     # Table 2: Experience of COVID-19  ------------------------------

#CIQ_ThinksCovid Ppt thinks they had COVID-19
summary(Descriptives$CIQ_ThinksCovid) #N
round(table(na.omit(Descriptives$CIQ_ThinksCovid))/
          length(na.omit(Descriptives$CIQ_ThinksCovid))*100, 2) #%

#CIQ_WhyThinksCovidNoTest
summary(Descriptives$CIQ_WhyThinksCovidNoTest)

#Symptoms
summary(Descriptives[,91:104])
symptoms <- as.data.frame(summary(Descriptives[,91:104]))
symptoms

#CIQ_Infected variables 
summary(Descriptives$CIQ_Infected_Shielded)
summary(Descriptives$CIQ_Infected_IsolatedDistanced) 
summary(Descriptives$CIQ_Infected_NHS111_GP) 
summary(Descriptives$CIQ_Infected_Swabbed) 
summary(Descriptives$CIQ_Infected_HubDoctor) 
summary(Descriptives$CIQ_Infected_AdmittedHospital) 

summary(Descriptives$CIQ_Infected_ICU) #none admitted to hosp - therefore all NA
summary(Descriptives$CIQ_Infected_Ventilator) #all NA

#CIQ_Infected_OtherInHousehold
summary(Descriptives$CIQ_Infected_OtherInHousehold) #NONE
#CIQ_Infected_OtherHowDiagnosed no household cases so all NA

#CIQ_SeriousRiskWarning (contacted by letter about shielding)
summary(Descriptives$CIQ_SeriousRiskWarning)
round(table(na.omit(Descriptives$CIQ_SeriousRiskWarning))/
          length(na.omit(Descriptives$CIQ_SeriousRiskWarning))*100, 2) #%

#CIQ_AvoidedMedicalCare (avoided contacting medical service)
summary(Descriptives$CIQ_AvoidedMedicalCare)
round(table(na.omit(Descriptives$CIQ_AvoidedMedicalCare))/
          length(na.omit(Descriptives$CIQ_AvoidedMedicalCare))*100, 2) #%

#CIQ_TimesLeaveHouse number of times leave house since lockdown
summary(Descriptives$CIQ_SinceMarch23_TimesLeaveHouse)
round(table(na.omit(Descriptives$CIQ_SinceMarch23_TimesLeaveHouse))/
          length(na.omit(Descriptives$CIQ_SinceMarch23_TimesLeaveHouse))*100, 2) #%

summary(Descriptives$CIQ_LikelyProximity)
round(table(na.omit(Descriptives$CIQ_LikelyProximity))/
          length(na.omit(Descriptives$CIQ_LikelyProximity))*100, 2) #%


# Table 3: knowledge and awareness of COVID-19 ----------------------------

#CIQ_Knowledge Rate knowledge of COVID
summary(Descriptives$CIQ_Knowledge)
plot(Descriptives$CIQ_Knowledge) #skewed
round(table(na.omit(Descriptives$CIQ_Knowledge))/
          length(na.omit(Descriptives$CIQ_Knowledge))*100, 2) #%

#CIQ_GovtGuidance How easy govt guidance to follow
summary(Descriptives$CIQ_GovtGuidance)
plot(Descriptives$CIQ_GovtGuidance) 
round(table(na.omit(Descriptives$CIQ_GovtGuidance))/
          length(na.omit(Descriptives$CIQ_GovtGuidance))*100, 2) #%

#CIQ_SocialDistance How often adhere to social distancing
summary(Descriptives$CIQ_SocialDistance)
round(table(na.omit(Descriptives$CIQ_SocialDistance))/
          length(na.omit(Descriptives$CIQ_SocialDistance))*100, 2) #%

#CIQ_StayHome How often adhere to staying home
summary(Descriptives$CIQ_StayHome)
round(table(na.omit(Descriptives$CIQ_StayHome))/
          length(na.omit(Descriptives$CIQ_StayHome))*100, 2) #%

#CIQ_StayHome How often adhere to self isolating
summary(Descriptives$CIQ_SelfIsolating)
round(table(na.omit(Descriptives$CIQ_SelfIsolating))/
          length(na.omit(Descriptives$CIQ_SelfIsolating))*100, 2) #%

#CIQ_StayHome How often adhere to hand washing
summary(Descriptives$CIQ_HandWash)
round(table(na.omit(Descriptives$CIQ_HandWash))/
          length(na.omit(Descriptives$CIQ_HandWash))*100, 2) #%

#CIQ_StayHome How often adhere to hand washing
summary(Descriptives$CIQ_StaysUpdated)
round(table(na.omit(Descriptives$CIQ_StaysUpdated))/
          length(na.omit(Descriptives$CIQ_StaysUpdated))*100, 2) #%

# Table 4: Living situation and behaviours and impact on day-to-da --------

#CIQ_LivesWith_HowMany (how many people in household)
describe(Descriptives$CIQ_LivesWith_HowMany)

#CID_LivesWith variables (who lives in household)
summary(Descriptives$CIQ_LivesWith_Alone)
summary(Descriptives$CIQ_LivesWith_Partner)
summary(Descriptives$CIQ_LivesWith_Child)
summary(Descriptives$CIQ_LivesWith_GrandChildren)
summary(Descriptives$CIQ_LivesWith_Parents)
summary(Descriptives$CIQ_LivesWith_OtherFam)
summary(Descriptives$CIQ_LivesWith_Carer)
summary(Descriptives$CIQ_LivesWith_Friends)

#CID_LivSituChange Has living situation changed due to covid
summary(Descriptives$CIQ_LivSituChange_Check)
round(table(na.omit(Descriptives$CIQ_LivSituChange_Check))/
          length(na.omit(Descriptives$CIQ_LivSituChange_Check))*100, 2) #%
summary(Descriptives$CIQ_LivSituChange_Residence)
summary(Descriptives$CIQ_LivSituChange_MorePeople)
summary(Descriptives$CIQ_LivSituChange_LessPeople)

which(Descriptives$CIQ_LivSituChange_Check == "Yes")
Descriptives$CIQ_LivSituChange_MorePeople[c(175,196,203,403,557,598,771,819,1063)]
Descriptives$CIQ_LivSituChange_Residence[c(175,196,203,403,557,598,771,819,1063)]

#CIQ_CarerGChildren Caring for grandchildren since covid
summary(Descriptives$CIQ_CarerGChildren)
round(table(na.omit(Descriptives$CIQ_CarerGChildren))/
          length(na.omit(Descriptives$CIQ_CarerGChildren))*100, 2) #%

#CIQ_Carer variable (making 1 variable of carer items)
Descriptives$Carer <- Descriptives$CIQ_CarerLiveWith
levels(Descriptives$Carer) <- c('Yes, for someone who lives with me', 
                                'Yes, for someone who does not live with me', 
                                'No, not a carer')
Descriptives$Carer[which(Descriptives$CIQ_CarerNotLivesWith == 'Yes, for someone who does not live with me')] <- 'Yes, for someone who does not live with me'
Descriptives$Carer[which(Descriptives$CIQ_NotCarer == 'No, not a carer')] <- 'No, not a carer'
levels(Descriptives$Carer)
summary(Descriptives$Carer)
round(table(na.omit(Descriptives$Carer))/
          length(na.omit(Descriptives$Carer))*100, 2) #%

Descriptives$CarerPrev <- Descriptives$CIQ_CarerLivesWithBeforeMarch23
levels(Descriptives$CarerPrev) <- c('Yes, for someone who lives with me', 
                                    'Yes, for someone who does not live with me', 
                                    'No, not a carer')
Descriptives$CarerPrev[which(Descriptives$CIQ_CarerNotLivesWithBeforeMarch23 == 'Yes, for someone who does not live with me')] <- 'Yes, for someone who does not live with me'
Descriptives$CarerPrev[which(Descriptives$CIQ_NotCarerBeforeMarch23 == 'No, not a carer')] <- 'No, not a carer'
levels(Descriptives$CarerPrev)
summary(Descriptives$CarerPrev)
round(table(na.omit(Descriptives$CarerPrev))/
          length(na.omit(Descriptives$CarerPrev))*100, 2) #%

summary(Descriptives$CIQ_CarerExpChangeSinceMarch23)
round(table(na.omit(Descriptives$CIQ_CarerExpChangeSinceMarch23))/
          length(na.omit(Descriptives$CarerPrev))*100, 2) #%

#CIQ_Prescriptions (creating 1 variable)
Descriptives$CIQ_Prescriptions <- Descriptives$CIQ_Prescriptions_NoChange
levels(Descriptives$CIQ_Prescriptions) <- c('None', 'Increased Supply', 'N/A', 'Switched to delivery', 'Other')
Descriptives$CIQ_Prescriptions[which(Descriptives$CIQ_Prescriptions_SwitchToDelivered == 'Switched order to mail order or delivery')] <- 'Switched to delivery'
Descriptives$CIQ_Prescriptions[which(Descriptives$CIQ_Prescriptions_IncreasedSupply == 'Increased the prescription to a longer supply (e.g. 90-day supply)')] <- 'Increased Supply'
Descriptives$CIQ_Prescriptions[which(Descriptives$CIQ_Prescriptions_Other == 'Other')] <- 'Other'
Descriptives$CIQ_Prescriptions[which(Descriptives$CIQ_Prescriptions_NA == 'N/A')] <- 'N/A'
levels(Descriptives$CIQ_Prescriptions)
summary(Descriptives$CIQ_Prescriptions)
round(table(na.omit(Descriptives$CIQ_Prescriptions))/
          length(na.omit(Descriptives$CIQ_Prescriptions))*100, 2) #%
plot(Descriptives$CIQ_Prescriptions)

#CIQ_GetsHelp Daily help with tasks
summary(Descriptives$CIQ_GetsHelp)
round(table(na.omit(Descriptives$CIQ_GetsHelp))/
          length(na.omit(Descriptives$CIQ_GetsHelp))*100, 2) #%
summary(Descriptives$CIQ_GetsHelpGroceries)
round(table(na.omit(Descriptives$CIQ_GetsHelpGroceries))/
          length(na.omit(Descriptives$CIQ_GetsHelp))*100, 2) #%
summary(Descriptives$CIQ_GetsHelpMedicines)
round(table(na.omit(Descriptives$CIQ_GetsHelpMedicines))/
          length(na.omit(Descriptives$CIQ_GetsHelp))*100, 2) #%
summary(Descriptives$CIQ_GetsHelpOther)
round(table(na.omit(Descriptives$CIQ_GetsHelpOther))/
          length(na.omit(Descriptives$CIQ_GetsHelp))*100, 2) #%

#CIQ_LocalInitiatives local initiatives in area
summary(Descriptives$CIQ_LocalInitiatives)
round(table(na.omit(Descriptives$CIQ_LocalInitiatives))/
          length(na.omit(Descriptives$CIQ_LocalInitiatives))*100, 2) #%

#CIQ_HelpsOthers Assists others with daily tasks
summary(Descriptives$CIQ_HelpsOthers)
round(table(na.omit(Descriptives$CIQ_HelpsOthers))/
          length(na.omit(Descriptives$CIQ_HelpsOthers))*100, 2) #%

#CIQ_Cash variables (using cash alternatives, importance of using cash)
summary(Descriptives$CIQ_Cash_ChangeToCards)
round(table(na.omit(Descriptives$CIQ_Cash_ChangeToCards))/
          length(na.omit(Descriptives$CIQ_Cash_ChangeToCards))*100, 2) #%
summary(Descriptives$CIQ_Cash_CashImportance)
round(table(na.omit(Descriptives$CIQ_Cash_CashImportance))/
          length(na.omit(Descriptives$CIQ_Cash_CashImportance))*100, 2) #%

#CIQ_GdnAllot Garden/Allotment Access
Descriptives$CIQ_GdnAllot <- Descriptives$CIQ_GdnAllot_Garden
levels(Descriptives$CIQ_GdnAllot) <- c('Garden', 'Allotment', 'No')
Descriptives$CIQ_GdnAllot[which(Descriptives$CIQ_GdnAllot_Allotment == 'Yes, Allotment')] <- 'Allotment'
Descriptives$CIQ_GdnAllot[which(Descriptives$CIQ_GdnAllot_NoAccess == 'No')] <- 'No'
Descriptives$CIQ_GdnAllot[which(Descriptives$CIQ_GdnAllot_Garden == 'Yes, Garden')] <- 'Garden'
levels(Descriptives$CIQ_GdnAllot)
summary(Descriptives$CIQ_GdnAllot)
round(table(na.omit(Descriptives$CIQ_GdnAllot))/
          length(na.omit(Descriptives$CIQ_GdnAllot))*100, 2) #%
plot(Descriptives$CIQ_GdnAllot)

#CIQ_InternetUse
summary(Descriptives$CIQ_InternetUseChange)
round(table(na.omit(Descriptives$CIQ_InternetUseChange))/
          length(na.omit(Descriptives$CIQ_InternetUseChange))*100, 2) #%
summary(Descriptives$CIQ_InternetUseMoreInFuture)
round(table(na.omit(Descriptives$CIQ_InternetUseMoreInFuture))/
          length(na.omit(Descriptives$CIQ_InternetUseMoreInFuture))*100, 2) #%


# Table 5: Social connectedness  ------------------------------------------

#CIQ_Neighbour neighbour variables (more or less contact, positive or negative)
summary(Descriptives$CIQ_NeighbourContactMoreLess)
round(table(na.omit(Descriptives$CIQ_NeighbourContactMoreLess))/
          length(na.omit(Descriptives$CIQ_NeighbourContactMoreLess))*100, 2) #%
summary(Descriptives$CIQ_NeighbourContactPosNeg)
round(table(na.omit(Descriptives$CIQ_NeighbourContactPosNeg))/
          length(na.omit(Descriptives$CIQ_NeighbourContactPosNeg))*100, 2) #%

#CIQ_lonelines loneliness variable
summary(Descriptives$CIQ_lonelines)
round(table(na.omit(Descriptives$CIQ_lonelines))/
          length(na.omit(Descriptives$CIQ_lonelines))*100, 2) #%

# Table 6: Health and lifestyle -------------------------------------------

#CIQ_MentalHealth
summary(Descriptives$CIQ_BeforeMarch23_MentalHealth)
round(table(na.omit(Descriptives$CIQ_BeforeMarch23_MentalHealth))/
          length(na.omit(Descriptives$CIQ_BeforeMarch23_MentalHealth))*100, 2) #%
summary(Descriptives$CIQ_SinceMarch23_MentalHealth)
round(table(na.omit(Descriptives$CIQ_SinceMarch23_MentalHealth))/
          length(na.omit(Descriptives$CIQ_SinceMarch23_MentalHealth))*100, 2) #%
summary(Descriptives$CIQ_LastTwoWeeks_Stress)
round(table(na.omit(Descriptives$CIQ_LastTwoWeeks_Stress))/
          length(na.omit(Descriptives$CIQ_LastTwoWeeks_Stress))*100, 2) #%

#CIQ_PhysicalHealth
summary(Descriptives$CIQ_BeforeMarch23_PhysicalHealth)
round(table(na.omit(Descriptives$CIQ_BeforeMarch23_PhysicalHealth))/
          length(na.omit(Descriptives$CIQ_BeforeMarch23_PhysicalHealth))*100, 2) #%
summary(Descriptives$CIQ_SinceMarch23_PhysicalHealth)
round(table(na.omit(Descriptives$CIQ_SinceMarch23_PhysicalHealth))/
          length(na.omit(Descriptives$CIQ_SinceMarch23_PhysicalHealth))*100, 2) #%

#CIQ_Alcohol
summary(Descriptives$CIQ_Alc_DrinksAlc)
round(table(na.omit(Descriptives$CIQ_Alc_DrinksAlc))/
          length(na.omit(Descriptives$CIQ_Alc_DrinksAlc))*100, 2) #%
summary(Descriptives$CIQ_Alc_Change)
round(table(na.omit(Descriptives$CIQ_Alc_Change))/
          length(na.omit(Descriptives$CIQ_Alc_Change))*100, 2) #%

#CIQ_Smoker
summary(Descriptives$CIQ_Smok_Smoker)
round(table(na.omit(Descriptives$CIQ_Smok_Smoker))/
          length(na.omit(Descriptives$CIQ_Smok_Smoker))*100, 2) #%
summary(Descriptives$CIQ_Smok_Change)
round(table(na.omit(Descriptives$CIQ_Smok_Change))/
          length(na.omit(Descriptives$CIQ_Smok_Change))*100, 2) #%

#CIQ_diet
summary(Descriptives$CIQ_Diet_ChangeHeathly)
round(table(na.omit(Descriptives$CIQ_Diet_ChangeHeathly))/
          length(na.omit(Descriptives$CIQ_Diet_ChangeHeathly))*100, 2) #%
summary(Descriptives$CIQ_Diet_ChangeIntake)
round(table(na.omit(Descriptives$CIQ_Diet_ChangeIntake))/
          length(na.omit(Descriptives$CIQ_Diet_ChangeIntake))*100, 2) #%

#CIQ_PhysActivChange Physical activity change
summary(Descriptives$CIQ_PhysActivChange)
round(table(na.omit(Descriptives$CIQ_PhysActivChange))/
          length(na.omit(Descriptives$CIQ_PhysActivChange))*100, 2) #%

#CIQ_DailyRoutineChange
summary(Descriptives$CIQ_DailyRoutineChange)
round(table(na.omit(Descriptives$CIQ_DailyRoutineChange))/
          length(na.omit(Descriptives$CIQ_DailyRoutineChange))*100, 2) #%

#CIQ_Pastime new pasttimes
summary(Descriptives$CIQ_Pastime_Exercise)
summary(Descriptives$CIQ_Pastime_Relax)
summary(Descriptives$CIQ_Pastime_Cooking)
summary(Descriptives$CIQ_Pastime_Language)
summary(Descriptives$CIQ_Pastime_ArtsCrafts)
summary(Descriptives$CIQ_Pastime_Sing)
summary(Descriptives$CIQ_Pastime_PlayingMusic)
summary(Descriptives$CIQ_Pastime_ListeningMusic)
summary(Descriptives$CIQ_Pastime_Reading)
summary(Descriptives$CIQ_Pastime_BoardGames)
summary(Descriptives$CIQ_Pastime_OnlineGames)
summary(Descriptives$CIQ_Pastime_Writing)
summary(Descriptives$CIQ_Pastime_TvFilms)
summary(Descriptives$CIQ_Pastime_RadioAudioBks)
summary(Descriptives$CIQ_Pastime_Baking)
summary(Descriptives$CIQ_Pastime_Gardening)
summary(Descriptives$CIQ_Pastime_Dancing)
summary(Descriptives$CIQ_Pastime_OnlineLearning)
summary(Descriptives$CIQ_Pastime_OtherActivity)
summary(Descriptives$CIQ_Pastime_NoChange)

# Save Descriptives data frame

save(Descriptives,file="Descriptives.Rda")
# Regression Ns filtered for non-wave 5s ----------------------------------
     
load("C:/Users/ataylo13/OneDrive - University of Edinburgh/COVID papers/Descriptives paper/New plan/New plan/Descriptives.Rda")

#filter data to only those who attended wave 5 before reporting Ns
 Descriptives <- filter(Descriptives, attendw5 == "Yes")
     
     summary(Descriptives$binaryinternet)
     summary(Descriptives$binarynervous)
     summary(Descriptives$getshelp)
     summary(Descriptives$pasttime)
     summary(Descriptives$lonely)
     
     summary(Descriptives$changephys)
     summary(Descriptives$routine)
     summary(Descriptives$leavehouse)
     
     summary(Descriptives$mentalhealthbefore)
     summary(Descriptives$mentalhealthafter)
     summary(Descriptives$physhealthbefore)
     summary(Descriptives$physhealthafter)
     

# Create working data set for comparisons in table 1 -------------------------------------------------

comparison <- data.frame(Descriptives$responder, Descriptives$attendw5, Descriptives$lbc36no, Descriptives$sex, Descriptives$agedays_SMS, Descriptives$age11, Descriptives$agedays_w1, Descriptives$age70, Descriptives$agedays_w4, Descriptives$age79, Descriptives$agedays_w5, Descriptives$age82, Descriptives$fathclass, Descriptives$pptclass, Descriptives$educ, Descriptives$marital, Descriptives$alone, Descriptives$resdnc, Descriptives$mmse82, Descriptives$mht11, Descriptives$adjustedmht11, Descriptives$mht79, Descriptives$adjustedmht79, Descriptives$srh, Descriptives$hibp, Descriptives$cvd, Descriptives$diab, Descriptives$sixm, Descriptives$grip, Descriptives$lung, Descriptives$townsend, Descriptives$hba1c, Descriptives$HADSD, Descriptives$emo, Descriptives$ex, Descriptives$con, Descriptives$healthlit, Descriptives$GHL, Descriptives$bmi, Descriptives$HADSA)

names(comparison) <- c('responder', 'attendw5', 'lbc36no', 'sex', 'agedays_SMS', 'age11', 'agedays_w1', 'age70', 'agedays_w4', 'age79', 'agedays_w5', 'age82', 'fathclass', 'pptclass', 'educ', 'marital', 'alone', 'resdnc', 'mmse82', 'mht11', 'adjustedmht11', 'mht79', 'adjustedmht79', 'srh', 'hibp', 'cvd', 'diab', 'sixm', 'grip', 'lung', 'townsend', 'hba1c', 'HADSD', 'emo', 'ex', 'con', 'healthlit', 'GHL', 'bmi', 'HADSA')
   
#save data files with only variables needed
save(comparison,file="comparison.Rda")


# Table 1 COMPARISONS ANALYSIS ----------------------------------------------------------------

load("C:/Users/ataylo13/OneDrive - University of Edinburgh/COVID papers/Descriptives paper/Data/comparison.Rda")

#filter data so results only for those who attended wave 5
comparison <- filter(comparison, attendw5 == "Yes")

## COMPARISON OF CONTINUOUS VARIABLES

# Age in years (mean age 82)
aggregate(x = comparison$age82,                
          by = list(comparison$responder),              
          FUN = mean, na.rm = T) 

group_by(comparison, responder) %>% summarise(mean = mean(age82), sd = sd(age82))

#t-test
t.test(age82 ~ responder, comparison, na.rm = T)
# t.test(age82 ~ responder, comparison, na.rm = T, var.equal = T) # assume equal variances

#cohens d
cohensD(age82 ~ responder, comparison, method = "unequal")

#ggboxplot
ggplot(data = comparison, mapping = aes(x = responder, y = age82)) + 
  geom_boxplot()

#boxplot
plot(comparison$age82 ~ comparison$responder)

# Childhood SES
aggregate(x = comparison$fathclass,                
          by = list(comparison$responder),              
          FUN = mean, na.rm = T) 

aggregate(x = comparison$fathclass,                
          by = list(comparison$responder),              
          FUN = sd, na.rm = T) 

group_by(comparison, responder) %>% summarise(mean = mean(fathclass), sd = sd(fathclass))

#t-test
t.test(fathclass ~ responder, comparison, na.rm = T)
# t.test(age82 ~ responder, comparison, na.rm = T, var.equal = T) # assume equal variances

#cohens d
cohensD(fathclass ~ responder, comparison, method = "unequal")

#ggboxplot
ggplot(data = comparison, mapping = aes(x = responder, y = fathclass)) + 
  geom_boxplot()

#boxplot
plot(comparison$fathclass ~ comparison$responder)

# Adulthood SES
aggregate(x = comparison$pptclass,                
          by = list(comparison$responder),              
          FUN = mean, na.rm = T) 

aggregate(x = comparison$pptclass,                
          by = list(comparison$responder),              
          FUN = sd, na.rm = T) 
group_by(comparison, responder) %>% summarise(mean = mean(pptclass), sd = sd(pptclass))

#t-test
t.test(pptclass ~ responder, comparison, na.rm = T)
# t.test(age82 ~ responder, comparison, na.rm = T, var.equal = T) # assume equal variances

#cohens d
cohensD(pptclass ~ responder, comparison, method = "unequal")

#ggboxplot
ggplot(data = comparison, mapping = aes(x = responder, y = pptclass)) + 
  geom_boxplot()

#boxplot
plot(comparison$pptclass ~ comparison$responder)

# Years of Education
aggregate(x = comparison$educ,                
          by = list(comparison$responder),              
          FUN = mean, na.rm = T) 

aggregate(x = comparison$educ,                
          by = list(comparison$responder),              
          FUN = sd, na.rm = T) 

group_by(comparison, responder) %>% summarise(mean = mean(educ), sd = sd(educ))

#t-test
t.test(educ ~ responder, comparison, na.rm = T)
# t.test(age82 ~ responder, comparison, na.rm = T, var.equal = T) # assume equal variances

#cohens d
cohensD(educ ~ responder, comparison, method = "unequal")

#ggboxplot
ggplot(data = comparison, mapping = aes(x = responder, y = educ)) + 
  geom_boxplot()

#boxplot
plot(comparison$educ ~ comparison$responder)

# MMSE
aggregate(x = comparison$mmse82,                
          by = list(comparison$responder),              
          FUN = mean, na.rm = T) 

aggregate(x = comparison$mmse82,                
          by = list(comparison$responder),              
          FUN = sd, na.rm = T) 

group_by(comparison, responder) %>% summarise(mean = mean(mmse82), sd = sd(mmse82))

#t-test
t.test(mmse82 ~ responder, comparison, na.rm = T)
# t.test(age82 ~ responder, comparison, na.rm = T, var.equal = T) # assume equal variances

#cohens d
cohensD(mmse82 ~ responder, comparison, method = "unequal")

#ggboxplot
ggplot(data = comparison, mapping = aes(x = responder, y = mmse82)) + 
  geom_boxplot()

#boxplot
plot(comparison$mmse82 ~ comparison$responder)

# MHT age 11
aggregate(x = comparison$mht11,                
          by = list(comparison$responder),              
          FUN = mean, na.rm = T) 

aggregate(x = comparison$mht11,                
          by = list(comparison$responder),              
          FUN = sd, na.rm = T) 

group_by(comparison, responder) %>% summarise(mean = mean(mht11), sd = sd(mht11))

#t-test
t.test(mht11 ~ responder, comparison, na.rm = T)
# t.test(age82 ~ responder, comparison, na.rm = T, var.equal = T) # assume equal variances

#cohens d
cohensD(mht11 ~ responder, comparison, method = "unequal")

#ggboxplot
ggplot(data = comparison, mapping = aes(x = responder, y = mht11)) + 
  geom_boxplot()

#boxplot
plot(comparison$mht11 ~ comparison$responder)

#Age-adjusted MHT 11
aggregate(x = comparison$adjustedmht11,                
          by = list(comparison$responder),              
          FUN = mean, na.rm = T) 

aggregate(x = comparison$adjustedmht11,                
          by = list(comparison$responder),              
          FUN = sd, na.rm = T) 

#t-test
t.test(adjustedmht11 ~ responder, comparison, na.rm = T)

#cohens d
cohensD(adjustedmht11 ~ responder, comparison, method = "unequal")

# Adjusted MHT 79
aggregate(x = comparison$adjustedmht79,                
          by = list(comparison$responder),              
          FUN = mean, na.rm = T) 

aggregate(x = comparison$adjustedmht79,                
          by = list(comparison$responder),              
          FUN = sd, na.rm = T) 

#t-test
t.test(adjustedmht79 ~ responder, comparison, na.rm = T)

#cohens d
cohensD(adjustedmht79 ~ responder, comparison, method = "unequal")

# MHT age 79
aggregate(x = comparison$mht79,                
          by = list(comparison$responder),              
          FUN = mean, na.rm = T) 

aggregate(x = comparison$mht79,                
          by = list(comparison$responder),              
          FUN = sd, na.rm = T) 

group_by(comparison, responder) %>% summarise(mean = mean(mht79), sd = sd(mht79))

#t-test
t.test(mht79 ~ responder, comparison, na.rm = T)
# t.test(age82 ~ responder, comparison, na.rm = T, var.equal = T) # assume equal variances

#cohens d
cohensD(mht79 ~ responder, comparison, method = "unequal")

#ggboxplot
ggplot(data = comparison, mapping = aes(x = responder, y = mht79)) + 
  geom_boxplot()

#boxplot
plot(comparison$mht79 ~ comparison$responder)

# General Health Literacy score (ranked score from spss)
aggregate(x = comparison$GHL,                
          by = list(comparison$responder),              
          FUN = mean, na.rm = T) 

aggregate(x = comparison$GHL,                
          by = list(comparison$responder),              
          FUN = sd, na.rm = T) 

group_by(comparison, responder) %>% summarise(mean = mean(GHL), sd = sd(GHL))

#t-test
t.test(GHL ~ responder, comparison, na.rm = T)
# t.test(age82 ~ responder, comparison, na.rm = T, var.equal = T) # assume equal variances

#cohens d
cohensD(GHL ~ responder, comparison, method = "unequal")

#ggboxplot
ggplot(data = comparison, mapping = aes(x = responder, y = healthlit)) + 
  geom_boxplot()

#boxplot
plot(comparison$healthlit ~ comparison$responder)

# BMI
aggregate(x = comparison$bmi,                
          by = list(comparison$responder),              
          FUN = mean, na.rm = T) 

aggregate(x = comparison$bmi,                
          by = list(comparison$responder),              
          FUN = sd, na.rm = T) 

group_by(comparison, responder) %>% summarise(mean = mean(bmi), sd = sd(bmi))

#t-test
t.test(bmi ~ responder, comparison, na.rm = T)
# t.test(age82 ~ responder, comparison, na.rm = T, var.equal = T) # assume equal variances

#cohens d
cohensD(bmi ~ responder, comparison, method = "unequal")

#ggboxplot
ggplot(data = comparison, mapping = aes(x = responder, y = bmi)) + 
  geom_boxplot()

#boxplot
plot(comparison$bmi ~ comparison$responder)

# Grip strength
aggregate(x = comparison$grip,                
          by = list(comparison$responder),              
          FUN = mean, na.rm = T) 

aggregate(x = comparison$grip,                
          by = list(comparison$responder),              
          FUN = sd, na.rm = T) 

group_by(comparison, responder) %>% summarise(mean = mean(grip), sd = sd(grip))

#t-test
t.test(grip ~ responder, comparison, na.rm = T)
# t.test(age82 ~ responder, comparison, na.rm = T, var.equal = T) # assume equal variances

#cohens d
cohensD(grip ~ responder, comparison, method = "unequal")

#ggboxplot
ggplot(data = comparison, mapping = aes(x = responder, y = grip)) + 
  geom_boxplot()

#boxplot
plot(comparison$grip ~ comparison$responder)

# Lung function (FEV1)
aggregate(x = comparison$lung,                
          by = list(comparison$responder),              
          FUN = mean, na.rm = T) 

aggregate(x = comparison$lung,                
          by = list(comparison$responder),              
          FUN = sd, na.rm = T) 

group_by(comparison, responder) %>% summarise(mean = mean(lung), sd = sd(lung))

#t-test
t.test(lung ~ responder, comparison, na.rm = T)
# t.test(age82 ~ responder, comparison, na.rm = T, var.equal = T) # assume equal variances

#cohens d
cohensD(lung ~ responder, comparison, method = "unequal")

#ggboxplot
ggplot(data = comparison, mapping = aes(x = responder, y = lung)) + 
  geom_boxplot()

#boxplot
plot(comparison$lung ~ comparison$responder)

# HbA1c
aggregate(x = comparison$hba1c,                
          by = list(comparison$responder),              
          FUN = mean, na.rm = T) 

aggregate(x = comparison$hba1c,                
          by = list(comparison$responder),              
          FUN = sd, na.rm = T) 

group_by(comparison, responder) %>% summarise(mean = mean(hba1c), sd = sd(hba1c))

#t-test
t.test(hba1c ~ responder, comparison, na.rm = T)
# t.test(age82 ~ responder, comparison, na.rm = T, var.equal = T) # assume equal variances

#cohens d
cohensD(hba1c ~ responder, comparison, method = "unequal")

#ggboxplot
ggplot(data = comparison, mapping = aes(x = responder, y = hba1c)) + 
  geom_boxplot()

#boxplot
plot(comparison$hba1c ~ comparison$responder)

# Townsend Disability Scale 
aggregate(x = comparison$townsend,                
          by = list(comparison$responder),              
          FUN = mean, na.rm = T) 

aggregate(x = comparison$townsend,                
          by = list(comparison$responder),              
          FUN = sd, na.rm = T) 

group_by(comparison, responder) %>% summarise(mean = mean(townsend), sd = sd(townsend))

#t-test
t.test(townsend ~ responder, comparison, na.rm = T)
# t.test(age82 ~ responder, comparison, na.rm = T, var.equal = T) # assume equal variances

#cohens d
cohensD(townsend ~ responder, comparison, method = "unequal")

#ggboxplot
ggplot(data = comparison, mapping = aes(x = responder, y = townsend)) + 
  geom_boxplot()

#boxplot
plot(comparison$townsend ~ comparison$responder)

# HADS Anxiety
aggregate(x = comparison$HADSA,                
          by = list(comparison$responder),              
          FUN = mean, na.rm = T) 

aggregate(x = comparison$HADSA,                
          by = list(comparison$responder),              
          FUN = sd, na.rm = T) 

group_by(comparison, responder) %>% summarise(mean = mean(HADSA), sd = sd(HADSA))

#t-test
t.test(HADSA ~ responder, comparison, na.rm = T)
# t.test(age82 ~ responder, comparison, na.rm = T, var.equal = T) # assume equal variances

#cohens d
cohensD(HADSA ~ responder, comparison, method = "unequal")

#ggboxplot
ggplot(data = comparison, mapping = aes(x = responder, y = HADSA)) + 
  geom_boxplot()

#boxplot
plot(comparison$HADSA ~ comparison$responder)

# HADSD
aggregate(x = comparison$HADSD,                
          by = list(comparison$responder),              
          FUN = mean, na.rm = T) 

aggregate(x = comparison$HADSD,                
          by = list(comparison$responder),              
          FUN = sd, na.rm = T) 

group_by(comparison, responder) %>% summarise(mean = mean(HADSD), sd = sd(HADSD))

#t-test
t.test(HADSD ~ responder, comparison, na.rm = T)
# t.test(age82 ~ responder, comparison, na.rm = T, var.equal = T) # assume equal variances

#cohens d
cohensD(HADSD ~ responder, comparison, method = "unequal")

#ggboxplot
ggplot(data = comparison, mapping = aes(x = responder, y = HADSD)) + 
  geom_boxplot()

#boxplot
plot(comparison$HADSD ~ comparison$responder)

# Emotional Stability
aggregate(x = comparison$emo,                
          by = list(comparison$responder),              
          FUN = mean, na.rm = T) 

aggregate(x = comparison$emo,                
          by = list(comparison$responder),              
          FUN = sd, na.rm = T) 

group_by(comparison, responder) %>% summarise(mean = mean(emo), sd = sd(emo))

#t-test
t.test(emo ~ responder, comparison, na.rm = T)
# t.test(age82 ~ responder, comparison, na.rm = T, var.equal = T) # assume equal variances

#cohens d
cohensD(emo ~ responder, comparison, method = "unequal")

#ggboxplot
ggplot(data = comparison, mapping = aes(x = responder, y = emo)) + 
  geom_boxplot()

#boxplot
plot(comparison$emo ~ comparison$responder)


# Extraversion
aggregate(x = comparison$ex,                
          by = list(comparison$responder),              
          FUN = mean, na.rm = T) 

aggregate(x = comparison$ex,                
          by = list(comparison$responder),              
          FUN = sd, na.rm = T) 

group_by(comparison, responder) %>% summarise(mean = mean(ex), sd = sd(ex))

#t-test
t.test(ex ~ responder, comparison, na.rm = T)
# t.test(age82 ~ responder, comparison, na.rm = T, var.equal = T) # assume equal variances

#cohens d
cohensD(ex ~ responder, comparison, method = "unequal")

#ggboxplot
ggplot(data = comparison, mapping = aes(x = responder, y = ex)) + 
  geom_boxplot()

#boxplot
plot(comparison$ex ~ comparison$responder)

# Conscientiousness
aggregate(x = comparison$con,                
          by = list(comparison$responder),              
          FUN = mean, na.rm = T) 

aggregate(x = comparison$con,                
          by = list(comparison$responder),              
          FUN = sd, na.rm = T) 

group_by(comparison, responder) %>% summarise(mean = mean(con), sd = sd(con))

#t-test
t.test(con ~ responder, comparison, na.rm = T)
# t.test(age82 ~ responder, comparison, na.rm = T, var.equal = T) # assume equal variances

#cohens d
cohensD(con ~ responder, comparison, method = "unequal")

#ggboxplot
ggplot(data = comparison, mapping = aes(x = responder, y = con)) + 
  geom_boxplot()

#boxplot
plot(comparison$con ~ comparison$responder)

# cohort differences in descriptives (chi-squared tests for Categorical differences) 

# SEX
# build a contingency table for sex ~ cohort

sex_table <- xtabs(~ sex + responder, comparison)
sex_table

#get observed counts, the expected counts and the degrees of freedom.

sex_table <- addmargins (sex_table)

# run chi-squared test of independence for sex
chisq.test(comparison$sex, comparison$responder)

# Marital status
marital_table <- xtabs(~ marital + responder, comparison)
marital_table
plot(marital_table)
chisq <- chisq.test(comparison$marital, comparison$responder)
chisq
chisq$observed
chisq$expected
fisher.test(marital_table) # because expected counts too low

marital_table2 <- xtabs(~ binarymarital + responder, Descriptives)
marital_table2
plot(marital_table2)
chisq <- chisq.test(Descriptives$binarymarital, Descriptives$responder)
chisq
chisq$observed
chisq$expected
fisher.test(marital_table) # because expected counts too low


# Live alone
alone_table <- xtabs(~ alone + responder, comparison)
alone_table
plot(alone_table)
chisq <- chisq.test(comparison$alone, comparison$responder)
chisq
chisq$observed
chisq$expected
fisher.test(alone_table) # because expected counts too low

# Hypertension
hibp_table <- xtabs(~ hibp + responder, comparison)
hibp_table
plot(hibp_table)
chisq <- chisq.test(comparison$hibp, comparison$responder)
chisq
chisq$observed
chisq$expected
fisher.test(hibp_table) # because expected counts too low

# CVD
cvd_table <- xtabs(~ cvd + responder, comparison)
cvd_table
plot(cvd_table)
chisq <- chisq.test(comparison$cvd, comparison$responder)
chisq
chisq$observed
chisq$expected
fisher.test(cvd_table) # because expected counts too low

# Diabetes
diab_table <- xtabs(~ diab + responder, comparison)
diab_table
plot(diab_table)
chisq <- chisq.test(comparison$diab, comparison$responder)
chisq
chisq$observed
chisq$expected
fisher.test(diab_table) # because expected counts too low

# Self-reported health
srh_table <- xtabs(~ srh + responder, comparison)
srh_table
plot(srh_table)
chisq <- chisq.test(comparison$srh, comparison$responder)
chisq
chisq$observed
chisq$expected
fisher.test(srh_table) # because expected counts too low


# Correlations ------------------------------------------------------------

#create numeric variables:
Descriptives$knowledge <- as.numeric(Descriptives$knowledge)
describe(Descriptives$knowledge)

Descriptives$guidance <- as.numeric(Descriptives$guidance)
describe(Descriptives$guidance)

Descriptives$avoid <- as.numeric(Descriptives$avoid)
describe(Descriptives$avoid)

Descriptives$leavehouse <- as.numeric(Descriptives$leavehouse)
describe(Descriptives$leavehouse)

Descriptives$help <- as.numeric(Descriptives$help)
describe(Descriptives$help)

Descriptives$internet <- as.numeric(Descriptives$internet)
describe(Descriptives$internet)

Descriptives$covidstress <- as.numeric(Descriptives$covidstress)
describe(Descriptives$covidstress)

Descriptives$mentalhealth <- as.numeric(Descriptives$mentalhealth)
describe(Descriptives$mentalhealth)

Descriptives$physicalhealth <- as.numeric(Descriptives$physicalhealth)
describe(Descriptives$physicalhealth)

Descriptives$alcohol <- as.numeric(Descriptives$alcohol)
describe(Descriptives$alcohol)

Descriptives$dietintake <- as.numeric(Descriptives$dietintake)
describe(Descriptives$dietintake)

Descriptives$healthydiet <- as.numeric(Descriptives$healthydiet)
describe(Descriptives$healthydiet)

Descriptives$physactivity <- as.numeric(Descriptives$physactivity)
describe(Descriptives$physactivity)

Descriptives$sex <- as.numeric(Descriptives$sex)
describe(Descriptives$sex)

Descriptives$marital <- as.numeric(Descriptives$marital)
describe(Descriptives$marital)

Descriptives$binarymarital <- as.numeric(Descriptives$binarymarital)
describe(Descriptives$binarymarital)

Descriptives$alone <- as.numeric(Descriptives$alone)
describe(Descriptives$alone)

Descriptives$covidalone <- as.numeric(Descriptives$covidalone)
describe(Descriptives$covidalone)

Descriptives$geog <- as.numeric(Descriptives$geog)
describe(Descriptives$geog)

Descriptives$hibp <- as.numeric(Descriptives$hibp)
describe(Descriptives$hibp)

Descriptives$cvd <- as.numeric(Descriptives$cvd)
describe(Descriptives$cvd)

Descriptives$diab <- as.numeric(Descriptives$diab)
describe(Descriptives$diab)

Descriptives$srh <- as.numeric(Descriptives$srh)
describe(Descriptives$srh)

#correlations

#filter data so results only for those who attended wave 5
Descriptives <- filter (Descriptives, attendw5 == "Yes")

# Age in days 82 ----------------------------------------------------------
with(Descriptives, corr.test(agedays_w5, knowledge, use = "pairwise.complete.obs", method = "spearman"))

with(Descriptives, corr.test(agedays_w5, guidance, use = "pairwise.complete.obs", method = "spearman"))

with(Descriptives, corr.test(agedays_w5, adherence, use = "pairwise.complete.obs", method = "spearman"))

with(Descriptives, corr.test(agedays_w5, avoid, use = "pairwise.complete.obs", method = "spearman"))

with(Descriptives, corr.test(agedays_w5, leavehouse, use = "pairwise.complete.obs", method = "spearman"))

with(Descriptives, corr.test(agedays_w5, help, use = "pairwise.complete.obs", method = "spearman"))

with(Descriptives, corr.test(agedays_w5, internet, use = "pairwise.complete.obs", method = "spearman"))

with(Descriptives, corr.test(agedays_w5, covidstress, use = "pairwise.complete.obs", method = "spearman"))

with(Descriptives, corr.test(agedays_w5, mentalhealth, use = "pairwise.complete.obs", method = "spearman"))

with(Descriptives, corr.test(agedays_w5, physicalhealth, use = "pairwise.complete.obs", method = "spearman"))

with(Descriptives, corr.test(agedays_w5, alcohol, use = "pairwise.complete.obs", method = "spearman"))

with(Descriptives, corr.test(agedays_w5, dietintake, use = "pairwise.complete.obs", method = "spearman"))

with(Descriptives, corr.test(agedays_w5, healthydiet, use = "pairwise.complete.obs", method = "spearman"))

with(Descriptives, corr.test(agedays_w5, physactivity, use = "pairwise.complete.obs", method = "spearman"))


# sex ---------------------------------------------------------------------

with(Descriptives, corr.test(sex, knowledge, use = "pairwise.complete.obs", method = "spearman"))

with(Descriptives, corr.test(sex, guidance, use = "pairwise.complete.obs", method = "spearman"))

print(with(Descriptives, corr.test(sex, adherence, use = "pairwise.complete.obs", method = "spearman")), digits=3)

with(Descriptives, corr.test(sex, avoid, use = "pairwise.complete.obs", method = "spearman"))

with(Descriptives, corr.test(sex, leavehouse, use = "pairwise.complete.obs", method = "spearman"))

with(Descriptives, corr.test(sex, help, use = "pairwise.complete.obs", method = "spearman"))

with(Descriptives, corr.test(sex, internet, use = "pairwise.complete.obs", method = "spearman"))

with(Descriptives, corr.test(sex, covidstress, use = "pairwise.complete.obs", method = "spearman"))

with(Descriptives, corr.test(sex, mentalhealth, use = "pairwise.complete.obs", method = "spearman"))

with(Descriptives, corr.test(sex, physicalhealth, use = "pairwise.complete.obs", method = "spearman"))

with(Descriptives, corr.test(sex, alcohol, use = "pairwise.complete.obs", method = "spearman"))

with(Descriptives, corr.test(sex, dietintake, use = "pairwise.complete.obs", method = "spearman"))

with(Descriptives, corr.test(sex, healthydiet, use = "pairwise.complete.obs", method = "spearman"))

with(Descriptives, corr.test(sex, physactivity, use = "pairwise.complete.obs", method = "spearman"))


# SES ---------------------------------------------------------------------

with(Descriptives, corr.test(pptclass, knowledge, use = "pairwise.complete.obs", method = "spearman"))

with(Descriptives, corr.test(pptclass, guidance, use = "pairwise.complete.obs", method = "spearman"))

with(Descriptives, corr.test(pptclass, adherence, use = "pairwise.complete.obs", method = "spearman"))

with(Descriptives, corr.test(pptclass, avoid, use = "pairwise.complete.obs", method = "spearman"))

with(Descriptives, corr.test(pptclass, leavehouse, use = "pairwise.complete.obs", method = "spearman"))

with(Descriptives, corr.test(pptclass, help, use = "pairwise.complete.obs", method = "spearman"))

with(Descriptives, corr.test(pptclass, internet, use = "pairwise.complete.obs", method = "spearman"))

with(Descriptives, corr.test(pptclass, covidstress, use = "pairwise.complete.obs", method = "spearman"))

with(Descriptives, corr.test(pptclass, mentalhealth, use = "pairwise.complete.obs", method = "spearman"))

with(Descriptives, corr.test(pptclass, physicalhealth, use = "pairwise.complete.obs", method = "spearman"))

with(Descriptives, corr.test(pptclass, alcohol, use = "pairwise.complete.obs", method = "spearman"))

with(Descriptives, corr.test(pptclass, dietintake, use = "pairwise.complete.obs", method = "spearman"))

with(Descriptives, corr.test(pptclass, healthydiet, use = "pairwise.complete.obs", method = "spearman"))

with(Descriptives, corr.test(pptclass, physactivity, use = "pairwise.complete.obs", method = "spearman"))


# Marital status ----------------------------------------------------------

with(Descriptives, corr.test(marital, knowledge, use = "pairwise.complete.obs", method = "spearman"))

with(Descriptives, corr.test(marital, guidance, use = "pairwise.complete.obs", method = "spearman"))

#with(Descriptives, corr.test(marital, adherence, use = "pairwise.complete.obs", #method = "spearman"))

with(Descriptives, corr.test(marital, avoid, use = "pairwise.complete.obs", method = "spearman"))

with(Descriptives, corr.test(marital, leavehouse, use = "pairwise.complete.obs", method = "spearman"))

with(Descriptives, corr.test(marital, help, use = "pairwise.complete.obs", method = "spearman"))

with(Descriptives, corr.test(marital, internet, use = "pairwise.complete.obs", method = "spearman"))

with(Descriptives, corr.test(marital, covidstress, use = "pairwise.complete.obs", method = "spearman"))

with(Descriptives, corr.test(marital, mentalhealth, use = "pairwise.complete.obs", method = "spearman"))

with(Descriptives, corr.test(marital, physicalhealth, use = "pairwise.complete.obs", method = "spearman"))

with(Descriptives, corr.test(marital, alcohol, use = "pairwise.complete.obs", method = "spearman"))

with(Descriptives, corr.test(marital, dietintake, use = "pairwise.complete.obs", method = "spearman"))

with(Descriptives, corr.test(marital, healthydiet, use = "pairwise.complete.obs", method = "spearman"))

with(Descriptives, corr.test(marital, physactivity, use = "pairwise.complete.obs", method = "spearman"))


# Binary marital status ---------------------------------------------------

print(with(Descriptives, corr.test(binarymarital, knowledge, use = "pairwise.complete.obs", method = "spearman")), digits=3)

print(with(Descriptives, corr.test(binarymarital, guidance, use = "pairwise.complete.obs", method = "spearman")), digits=3)

print(with(Descriptives, corr.test(binarymarital, adherence, use = "pairwise.complete.obs", method = "spearman")), digits=3)

print(with(Descriptives, corr.test(binarymarital, avoid, use = "pairwise.complete.obs", method = "spearman")), digits=3)

print(with(Descriptives, corr.test(binarymarital, leavehouse, use = "pairwise.complete.obs", method = "spearman")), digits=3)

print(with(Descriptives, corr.test(binarymarital, help, use = "pairwise.complete.obs", method = "spearman")),digits=3)

print(with(Descriptives, corr.test(binarymarital, internet, use = "pairwise.complete.obs", method = "spearman")), digits=3)

print(with(Descriptives, corr.test(binarymarital, covidstress, use = "pairwise.complete.obs", method = "spearman")), digits=3)

print(with(Descriptives, corr.test(binarymarital, mentalhealth, use = "pairwise.complete.obs", method = "spearman")), digits=3)

print(with(Descriptives, corr.test(binarymarital, physicalhealth, use = "pairwise.complete.obs", method = "spearman")), digits=3)

print(with(Descriptives, corr.test(binarymarital, alcohol, use = "pairwise.complete.obs", method = "spearman")), digits=3)

print(with(Descriptives, corr.test(binarymarital, dietintake, use = "pairwise.complete.obs", method = "spearman")), digits=3)

print(with(Descriptives, corr.test(binarymarital, healthydiet, use = "pairwise.complete.obs", method = "spearman")), digits=3)

print(with(Descriptives, corr.test(binarymarital, physactivity, use = "pairwise.complete.obs", method = "spearman")), digits=3)

# Living alone ----------------------------------------------------------

with(Descriptives, corr.test(alone, knowledge, use = "pairwise.complete.obs", method = "spearman"))

with(Descriptives, corr.test(alone, guidance, use = "pairwise.complete.obs", method = "spearman"))

#with(Descriptives, corr.test(alone, adherence, use = "pairwise.complete.obs", #method = "spearman"))

with(Descriptives, corr.test(alone, avoid, use = "pairwise.complete.obs", method = "spearman"))

with(Descriptives, corr.test(alone, leavehouse, use = "pairwise.complete.obs", method = "spearman"))

with(Descriptives, corr.test(alone, help, use = "pairwise.complete.obs", method = "spearman"))

with(Descriptives, corr.test(alone, internet, use = "pairwise.complete.obs", method = "spearman"))

with(Descriptives, corr.test(alone, covidstress, use = "pairwise.complete.obs", method = "spearman"))

with(Descriptives, corr.test(alone, mentalhealth, use = "pairwise.complete.obs", method = "spearman"))

with(Descriptives, corr.test(alone, physicalhealth, use = "pairwise.complete.obs", method = "spearman"))

with(Descriptives, corr.test(alone, alcohol, use = "pairwise.complete.obs", method = "spearman"))

with(Descriptives, corr.test(alone, dietintake, use = "pairwise.complete.obs", method = "spearman"))

with(Descriptives, corr.test(alone, healthydiet, use = "pairwise.complete.obs", method = "spearman"))

with(Descriptives, corr.test(alone, physactivity, use = "pairwise.complete.obs", method = "spearman"))


# Living alone from covid questionnaire -----------------------------------
print(with(Descriptives, corr.test(covidalone, knowledge, use = "pairwise.complete.obs", method = "spearman")), digits=3)

print(with(Descriptives, corr.test(covidalone, guidance, use = "pairwise.complete.obs", method = "spearman")), digits=3)

print(with(Descriptives, corr.test(covidalone, adherence, use = "pairwise.complete.obs", method = "spearman")), digits=3)

print(with(Descriptives, corr.test(covidalone, avoid, use = "pairwise.complete.obs", method = "spearman")), digits=3)

print(with(Descriptives, corr.test(covidalone, leavehouse, use = "pairwise.complete.obs", method = "spearman")), digits=3)

print(with(Descriptives, corr.test(covidalone, help, use = "pairwise.complete.obs", method = "spearman")),digits=3)

print(with(Descriptives, corr.test(covidalone, internet, use = "pairwise.complete.obs", method = "spearman")), digits=3)

print(with(Descriptives, corr.test(covidalone, covidstress, use = "pairwise.complete.obs", method = "spearman")), digits=3)

print(with(Descriptives, corr.test(covidalone, mentalhealth, use = "pairwise.complete.obs", method = "spearman")), digits=3)

print(with(Descriptives, corr.test(covidalone, physicalhealth, use = "pairwise.complete.obs", method = "spearman")), digits=3)

print(with(Descriptives, corr.test(covidalone, alcohol, use = "pairwise.complete.obs", method = "spearman")), digits=3)

print(with(Descriptives, corr.test(covidalone, dietintake, use = "pairwise.complete.obs", method = "spearman")), digits=3)

print(with(Descriptives, corr.test(covidalone, healthydiet, use = "pairwise.complete.obs", method = "spearman")), digits=3)

print(with(Descriptives, corr.test(covidalone, physactivity, use = "pairwise.complete.obs", method = "spearman")), digits=3)

# Geography of residence --------------------------------------------------

with(Descriptives, corr.test(geog, knowledge, use = "pairwise.complete.obs", method = "spearman"))

with(Descriptives, corr.test(geog, guidance, use = "pairwise.complete.obs", method = "spearman"))

with(Descriptives, corr.test(geog, adherence, use = "pairwise.complete.obs", method = "spearman"))

with(Descriptives, corr.test(geog, avoid, use = "pairwise.complete.obs", method = "spearman"))

with(Descriptives, corr.test(geog, leavehouse, use = "pairwise.complete.obs", method = "spearman"))

with(Descriptives, corr.test(geog, help, use = "pairwise.complete.obs", method = "spearman"))

with(Descriptives, corr.test(geog, internet, use = "pairwise.complete.obs", method = "spearman"))

with(Descriptives, corr.test(geog, covidstress, use = "pairwise.complete.obs", method = "spearman"))

with(Descriptives, corr.test(geog, mentalhealth, use = "pairwise.complete.obs", method = "spearman"))

with(Descriptives, corr.test(geog, physicalhealth, use = "pairwise.complete.obs", method = "spearman"))

with(Descriptives, corr.test(geog, alcohol, use = "pairwise.complete.obs", method = "spearman"))

with(Descriptives, corr.test(geog, dietintake, use = "pairwise.complete.obs", method = "spearman"))

with(Descriptives, corr.test(geog, healthydiet, use = "pairwise.complete.obs", method = "spearman"))

with(Descriptives, corr.test(geog, physactivity, use = "pairwise.complete.obs", method = "spearman"))


# age-adjusted mht11 ------------------------------------------------------

with(Descriptives, corr.test(adjustedmht11, knowledge, use = "pairwise.complete.obs", method = "spearman"))

with(Descriptives, corr.test(adjustedmht11, guidance, use = "pairwise.complete.obs", method = "spearman"))

with(Descriptives, corr.test(adjustedmht11, adherence, use = "pairwise.complete.obs", method = "spearman"))

with(Descriptives, corr.test(adjustedmht11, avoid, use = "pairwise.complete.obs", method = "spearman"))

with(Descriptives, corr.test(adjustedmht11, leavehouse, use = "pairwise.complete.obs", method = "spearman"))

with(Descriptives, corr.test(adjustedmht11, help, use = "pairwise.complete.obs", method = "spearman"))

with(Descriptives, corr.test(adjustedmht11, internet, use = "pairwise.complete.obs", method = "spearman"))

with(Descriptives, corr.test(adjustedmht11, covidstress, use = "pairwise.complete.obs", method = "spearman"))

with(Descriptives, corr.test(adjustedmht11, mentalhealth, use = "pairwise.complete.obs", method = "spearman"))

with(Descriptives, corr.test(adjustedmht11, physicalhealth, use = "pairwise.complete.obs", method = "spearman"))

with(Descriptives, corr.test(adjustedmht11, alcohol, use = "pairwise.complete.obs", method = "spearman"))

with(Descriptives, corr.test(adjustedmht11, dietintake, use = "pairwise.complete.obs", method = "spearman"))

with(Descriptives, corr.test(adjustedmht11, healthydiet, use = "pairwise.complete.obs", method = "spearman"))

with(Descriptives, corr.test(adjustedmht11, physactivity, use = "pairwise.complete.obs", method = "spearman"))


# age-adjusted mht79 ------------------------------------------------------

with(Descriptives, corr.test(adjustedmht79, knowledge, use = "pairwise.complete.obs", method = "spearman"))

with(Descriptives, corr.test(adjustedmht79, guidance, use = "pairwise.complete.obs", method = "spearman"))

with(Descriptives, corr.test(adjustedmht79, adherence, use = "pairwise.complete.obs", method = "spearman"))

with(Descriptives, corr.test(adjustedmht79, avoid, use = "pairwise.complete.obs", method = "spearman"))
with(Descriptives, plot(adjustedmht79, avoid))

with(Descriptives, corr.test(adjustedmht79, leavehouse, use = "pairwise.complete.obs", method = "spearman"))

with(Descriptives, corr.test(adjustedmht79, help, use = "pairwise.complete.obs", method = "spearman"))

with(Descriptives, corr.test(adjustedmht79, internet, use = "pairwise.complete.obs", method = "spearman"))

with(Descriptives, corr.test(adjustedmht79, covidstress, use = "pairwise.complete.obs", method = "spearman"))

with(Descriptives, corr.test(adjustedmht79, mentalhealth, use = "pairwise.complete.obs", method = "spearman"))

with(Descriptives, corr.test(adjustedmht79, physicalhealth, use = "pairwise.complete.obs", method = "spearman"))

with(Descriptives, corr.test(adjustedmht79, alcohol, use = "pairwise.complete.obs", method = "spearman"))

with(Descriptives, corr.test(adjustedmht79, dietintake, use = "pairwise.complete.obs", method = "spearman"))

with(Descriptives, corr.test(adjustedmht79, healthydiet, use = "pairwise.complete.obs", method = "spearman"))

with(Descriptives, corr.test(adjustedmht79, physactivity, use = "pairwise.complete.obs", method = "spearman"))


# grip strength -----------------------------------------------------------

with(Descriptives, corr.test(grip, knowledge, use = "pairwise.complete.obs", method = "spearman"))

with(Descriptives, corr.test(grip, guidance, use = "pairwise.complete.obs", method = "spearman"))

with(Descriptives, corr.test(grip, adherence, use = "pairwise.complete.obs", method = "spearman"))

with(Descriptives, corr.test(grip, avoid, use = "pairwise.complete.obs", method = "spearman"))
with(Descriptives, plot(grip, avoid))

with(Descriptives, corr.test(grip, leavehouse, use = "pairwise.complete.obs", method = "spearman"))

with(Descriptives, corr.test(grip, help, use = "pairwise.complete.obs", method = "spearman"))

with(Descriptives, corr.test(grip, internet, use = "pairwise.complete.obs", method = "spearman"))

with(Descriptives, corr.test(grip, covidstress, use = "pairwise.complete.obs", method = "spearman"))

with(Descriptives, corr.test(grip, mentalhealth, use = "pairwise.complete.obs", method = "spearman"))

with(Descriptives, corr.test(grip, physicalhealth, use = "pairwise.complete.obs", method = "spearman"))

with(Descriptives, corr.test(grip, alcohol, use = "pairwise.complete.obs", method = "spearman"))

with(Descriptives, corr.test(grip, dietintake, use = "pairwise.complete.obs", method = "spearman"))

with(Descriptives, corr.test(grip, healthydiet, use = "pairwise.complete.obs", method = "spearman"))

with(Descriptives, corr.test(grip, physactivity, use = "pairwise.complete.obs", method = "spearman"))

# Lung function (fev1) ----------------------------------------------------


with(Descriptives, corr.test(lung, knowledge, use = "pairwise.complete.obs", method = "spearman"))

with(Descriptives, corr.test(lung, guidance, use = "pairwise.complete.obs", method = "spearman"))
with(Descriptives, plot(lung, guidance))

print(with(Descriptives, corr.test(lung, adherence, use = "pairwise.complete.obs", method = "spearman")), digits = 3)

with(Descriptives, corr.test(lung, avoid, use = "pairwise.complete.obs", method = "spearman"))

with(Descriptives, corr.test(lung, leavehouse, use = "pairwise.complete.obs", method = "spearman"))

with(Descriptives, corr.test(lung, help, use = "pairwise.complete.obs", method = "spearman"))

with(Descriptives, corr.test(lung, internet, use = "pairwise.complete.obs", method = "spearman"))

with(Descriptives, corr.test(lung, covidstress, use = "pairwise.complete.obs", method = "spearman"))

with(Descriptives, corr.test(lung, mentalhealth, use = "pairwise.complete.obs", method = "spearman"))

with(Descriptives, corr.test(lung, physicalhealth, use = "pairwise.complete.obs", method = "spearman"))

with(Descriptives, corr.test(lung, alcohol, use = "pairwise.complete.obs", method = "spearman"))

with(Descriptives, corr.test(lung, dietintake, use = "pairwise.complete.obs", method = "spearman"))

with(Descriptives, corr.test(lung, healthydiet, use = "pairwise.complete.obs", method = "spearman"))

with(Descriptives, corr.test(lung, physactivity, use = "pairwise.complete.obs", method = "spearman"))

# BMI ---------------------------------------------------------------------

with(Descriptives, corr.test(bmi, knowledge, use = "pairwise.complete.obs", method = "spearman"))

with(Descriptives, corr.test(bmi, guidance, use = "pairwise.complete.obs", method = "spearman"))
with(Descriptives, plot(bmi, guidance))

with(Descriptives, corr.test(bmi, adherence, use = "pairwise.complete.obs", method = "spearman"))

with(Descriptives, corr.test(bmi, avoid, use = "pairwise.complete.obs", method = "spearman"))

with(Descriptives, corr.test(bmi, leavehouse, use = "pairwise.complete.obs", method = "spearman"))

with(Descriptives, corr.test(bmi, help, use = "pairwise.complete.obs", method = "spearman"))

with(Descriptives, corr.test(bmi, internet, use = "pairwise.complete.obs", method = "spearman"))

with(Descriptives, corr.test(bmi, covidstress, use = "pairwise.complete.obs", method = "spearman"))

with(Descriptives, corr.test(bmi, mentalhealth, use = "pairwise.complete.obs", method = "spearman"))

with(Descriptives, corr.test(bmi, physicalhealth, use = "pairwise.complete.obs", method = "spearman"))

with(Descriptives, corr.test(bmi, alcohol, use = "pairwise.complete.obs", method = "spearman"))

with(Descriptives, corr.test(bmi, dietintake, use = "pairwise.complete.obs", method = "spearman"))

with(Descriptives, corr.test(bmi, healthydiet, use = "pairwise.complete.obs", method = "spearman"))

with(Descriptives, corr.test(bmi, physactivity, use = "pairwise.complete.obs", method = "spearman"))

# cvd ---------------------------------------------------------------------

with(Descriptives, corr.test(cvd, knowledge, use = "pairwise.complete.obs", method = "spearman"))

with(Descriptives, corr.test(cvd, guidance, use = "pairwise.complete.obs", method = "spearman"))
with(Descriptives, plot(cvd, guidance))

print(with(Descriptives, corr.test(cvd, adherence, use = "pairwise.complete.obs", method = "spearman")), digits = 3)

with(Descriptives, corr.test(cvd, avoid, use = "pairwise.complete.obs", method = "spearman"))

with(Descriptives, corr.test(cvd, leavehouse, use = "pairwise.complete.obs", method = "spearman"))

with(Descriptives, corr.test(cvd, help, use = "pairwise.complete.obs", method = "spearman"))

with(Descriptives, corr.test(cvd, internet, use = "pairwise.complete.obs", method = "spearman"))

with(Descriptives, corr.test(cvd, covidstress, use = "pairwise.complete.obs", method = "spearman"))

with(Descriptives, corr.test(cvd, mentalhealth, use = "pairwise.complete.obs", method = "spearman"))

with(Descriptives, corr.test(cvd, physicalhealth, use = "pairwise.complete.obs", method = "spearman"))

print(with(Descriptives, corr.test(cvd, alcohol, use = "pairwise.complete.obs", method = "spearman")), digits=3)

print(with(Descriptives, corr.test(cvd, dietintake, use = "pairwise.complete.obs", method = "spearman")), digits=3)

print(with(Descriptives, corr.test(cvd, healthydiet, use = "pairwise.complete.obs", method = "spearman")), digits=3)

print(with(Descriptives, corr.test(cvd, physactivity, use = "pairwise.complete.obs", method = "spearman")), digits=3)


# diabetes ----------------------------------------------------------------

print(with(Descriptives, corr.test(diab, knowledge, use = "pairwise.complete.obs", method = "spearman")), digits=3)

print(with(Descriptives, corr.test(diab, guidance, use = "pairwise.complete.obs", method = "spearman")), digits=3)
with(Descriptives, plot(diab, guidance))

print(with(Descriptives, corr.test(diab, adherence, use = "pairwise.complete.obs", method = "spearman")), digits=3)

print(with(Descriptives, corr.test(diab, avoid, use = "pairwise.complete.obs", method = "spearman")), digits=3)

print(with(Descriptives, corr.test(diab, leavehouse, use = "pairwise.complete.obs", method = "spearman")), digits=3)

print(with(Descriptives, corr.test(diab, help, use = "pairwise.complete.obs", method = "spearman")),digits=3)

print(with(Descriptives, corr.test(diab, internet, use = "pairwise.complete.obs", method = "spearman")), digits=3)

print(with(Descriptives, corr.test(diab, covidstress, use = "pairwise.complete.obs", method = "spearman")), digits=3)

print(with(Descriptives, corr.test(diab, mentalhealth, use = "pairwise.complete.obs", method = "spearman")), digits=3)

print(with(Descriptives, corr.test(diab, physicalhealth, use = "pairwise.complete.obs", method = "spearman")), digits=3)

print(with(Descriptives, corr.test(diab, alcohol, use = "pairwise.complete.obs", method = "spearman")), digits=3)

print(with(Descriptives, corr.test(diab, dietintake, use = "pairwise.complete.obs", method = "spearman")), digits=3)

print(with(Descriptives, corr.test(diab, healthydiet, use = "pairwise.complete.obs", method = "spearman")), digits=3)

print(with(Descriptives, corr.test(diab, physactivity, use = "pairwise.complete.obs", method = "spearman")), digits=3)


# Hypertension ------------------------------------------------------------

print(with(Descriptives, corr.test(hibp, knowledge, use = "pairwise.complete.obs", method = "spearman")), digits=3)

print(with(Descriptives, corr.test(hibp, guidance, use = "pairwise.complete.obs", method = "spearman")), digits=3)
with(Descriptives, plot(diab, guidance))

print(with(Descriptives, corr.test(hibp, adherence, use = "pairwise.complete.obs", method = "spearman")), digits=3)

print(with(Descriptives, corr.test(hibp, avoid, use = "pairwise.complete.obs", method = "spearman")), digits=3)

print(with(Descriptives, corr.test(hibp, leavehouse, use = "pairwise.complete.obs", method = "spearman")), digits=3)

print(with(Descriptives, corr.test(hibp, help, use = "pairwise.complete.obs", method = "spearman")),digits=3)

print(with(Descriptives, corr.test(hibp, internet, use = "pairwise.complete.obs", method = "spearman")), digits=3)

print(with(Descriptives, corr.test(hibp, covidstress, use = "pairwise.complete.obs", method = "spearman")), digits=3)

print(with(Descriptives, corr.test(hibp, mentalhealth, use = "pairwise.complete.obs", method = "spearman")), digits=3)

print(with(Descriptives, corr.test(hibp, physicalhealth, use = "pairwise.complete.obs", method = "spearman")), digits=3)

print(with(Descriptives, corr.test(hibp, alcohol, use = "pairwise.complete.obs", method = "spearman")), digits=3)

print(with(Descriptives, corr.test(hibp, dietintake, use = "pairwise.complete.obs", method = "spearman")), digits=3)

print(with(Descriptives, corr.test(hibp, healthydiet, use = "pairwise.complete.obs", method = "spearman")), digits=3)

print(with(Descriptives, corr.test(hibp, physactivity, use = "pairwise.complete.obs", method = "spearman")), digits=3)


# hba1c -------------------------------------------------------------------

print(with(Descriptives, corr.test(hba1c, knowledge, use = "pairwise.complete.obs", method = "spearman")), digits=3)

print(with(Descriptives, corr.test(hba1c, guidance, use = "pairwise.complete.obs", method = "spearman")), digits=3)
with(Descriptives, plot(hba1c, guidance))

print(with(Descriptives, corr.test(hba1c, adherence, use = "pairwise.complete.obs", method = "spearman")), digits=3)

print(with(Descriptives, corr.test(hba1c, avoid, use = "pairwise.complete.obs", method = "spearman")), digits=3)

print(with(Descriptives, corr.test(hba1c, leavehouse, use = "pairwise.complete.obs", method = "spearman")), digits=3)

print(with(Descriptives, corr.test(hba1c, help, use = "pairwise.complete.obs", method = "spearman")),digits=3)

print(with(Descriptives, corr.test(hba1c, internet, use = "pairwise.complete.obs", method = "spearman")), digits=3)

print(with(Descriptives, corr.test(hba1c, covidstress, use = "pairwise.complete.obs", method = "spearman")), digits=3)

print(with(Descriptives, corr.test(hba1c, mentalhealth, use = "pairwise.complete.obs", method = "spearman")), digits=3)

print(with(Descriptives, corr.test(hba1c, physicalhealth, use = "pairwise.complete.obs", method = "spearman")), digits=3)

print(with(Descriptives, corr.test(hba1c, alcohol, use = "pairwise.complete.obs", method = "spearman")), digits=3)

print(with(Descriptives, corr.test(hba1c, dietintake, use = "pairwise.complete.obs", method = "spearman")), digits=3)

print(with(Descriptives, corr.test(hba1c, healthydiet, use = "pairwise.complete.obs", method = "spearman")), digits=3)

print(with(Descriptives, corr.test(hba1c, physactivity, use = "pairwise.complete.obs", method = "spearman")), digits=3)


# self reported health ----------------------------------------------------


print(with(Descriptives, corr.test(srh, knowledge, use = "pairwise.complete.obs", method = "spearman")),digits=3)

print(with(Descriptives, corr.test(srh, guidance, use = "pairwise.complete.obs", method = "spearman")), digits=3)
with(Descriptives, plot(srh, guidance))

print(with(Descriptives, corr.test(srh, adherence, use = "pairwise.complete.obs", method = "spearman")), digits=3)

print(with(Descriptives, corr.test(srh, avoid, use = "pairwise.complete.obs", method = "spearman")), digits=3)

print(with(Descriptives, corr.test(srh, leavehouse, use = "pairwise.complete.obs", method = "spearman")), digits=3)

print(with(Descriptives, corr.test(srh, help, use = "pairwise.complete.obs", method = "spearman")),digits=3)

print(with(Descriptives, corr.test(srh, internet, use = "pairwise.complete.obs", method = "spearman")), digits=3)

print(with(Descriptives, corr.test(srh, covidstress, use = "pairwise.complete.obs", method = "spearman")), digits=3)

print(with(Descriptives, corr.test(srh, mentalhealth, use = "pairwise.complete.obs", method = "spearman")), digits=3)

print(with(Descriptives, corr.test(srh, physicalhealth, use = "pairwise.complete.obs", method = "spearman")), digits=3)

print(with(Descriptives, corr.test(srh, alcohol, use = "pairwise.complete.obs", method = "spearman")), digits=3)

print(with(Descriptives, corr.test(srh, dietintake, use = "pairwise.complete.obs", method = "spearman")), digits=3)

print(with(Descriptives, corr.test(srh, healthydiet, use = "pairwise.complete.obs", method = "spearman")), digits=3)

print(with(Descriptives, corr.test(srh, physactivity, use = "pairwise.complete.obs", method = "spearman")), digits=3)


# Townsend disability -----------------------------------------------------

print(with(Descriptives, corr.test(townsend, knowledge, use = "pairwise.complete.obs", method = "spearman")),digits=3)

print(with(Descriptives, corr.test(townsend, guidance, use = "pairwise.complete.obs", method = "spearman")), digits=3)
with(Descriptives, plot(townsend, guidance))

print(with(Descriptives, corr.test(townsend, adherence, use = "pairwise.complete.obs", method = "spearman")), digits=3)

print(with(Descriptives, corr.test(townsend, avoid, use = "pairwise.complete.obs", method = "spearman")), digits=3)

print(with(Descriptives, corr.test(townsend, leavehouse, use = "pairwise.complete.obs", method = "spearman")), digits=3)

print(with(Descriptives, corr.test(townsend, help, use = "pairwise.complete.obs", method = "spearman")),digits=3)

print(with(Descriptives, corr.test(townsend, internet, use = "pairwise.complete.obs", method = "spearman")), digits=3)

print(with(Descriptives, corr.test(townsend, covidstress, use = "pairwise.complete.obs", method = "spearman")), digits=3)

print(with(Descriptives, corr.test(townsend, mentalhealth, use = "pairwise.complete.obs", method = "spearman")), digits=3)

print(with(Descriptives, corr.test(townsend, physicalhealth, use = "pairwise.complete.obs", method = "spearman")), digits=3)

print(with(Descriptives, corr.test(townsend, alcohol, use = "pairwise.complete.obs", method = "spearman")), digits=3)

print(with(Descriptives, corr.test(townsend, dietintake, use = "pairwise.complete.obs", method = "spearman")), digits=3)

print(with(Descriptives, corr.test(townsend, healthydiet, use = "pairwise.complete.obs", method = "spearman")), digits=3)

print(with(Descriptives, corr.test(townsend, physactivity, use = "pairwise.complete.obs", method = "spearman")), digits=3)


# General Health Literacy -------------------------------------------------

print(with(Descriptives, corr.test(GHL, knowledge, use = "pairwise.complete.obs", method = "spearman")),digits=3)

print(with(Descriptives, corr.test(GHL, guidance, use = "pairwise.complete.obs", method = "spearman")), digits=3)
with(Descriptives, plot(GHL, guidance))

print(with(Descriptives, corr.test(GHL, adherence, use = "pairwise.complete.obs", method = "spearman")), digits=3)

print(with(Descriptives, corr.test(GHL, avoid, use = "pairwise.complete.obs", method = "spearman")), digits=3)

print(with(Descriptives, corr.test(GHL, leavehouse, use = "pairwise.complete.obs", method = "spearman")), digits=3)

print(with(Descriptives, corr.test(GHL, help, use = "pairwise.complete.obs", method = "spearman")),digits=3)

print(with(Descriptives, corr.test(GHL, internet, use = "pairwise.complete.obs", method = "spearman")), digits=3)

print(with(Descriptives, corr.test(GHL, covidstress, use = "pairwise.complete.obs", method = "spearman")), digits=3)

print(with(Descriptives, corr.test(GHL, mentalhealth, use = "pairwise.complete.obs", method = "spearman")), digits=3)

print(with(Descriptives, corr.test(GHL, physicalhealth, use = "pairwise.complete.obs", method = "spearman")), digits=3)

print(with(Descriptives, corr.test(GHL, alcohol, use = "pairwise.complete.obs", method = "spearman")), digits=3)

print(with(Descriptives, corr.test(GHL, dietintake, use = "pairwise.complete.obs", method = "spearman")), digits=3)

print(with(Descriptives, corr.test(GHL, healthydiet, use = "pairwise.complete.obs", method = "spearman")), digits=3)

print(with(Descriptives, corr.test(GHL, physactivity, use = "pairwise.complete.obs", method = "spearman")), digits=3)

# HADS Anxiety -----------------------------------------------------------

print(with(Descriptives, corr.test(HADSA, knowledge, use = "pairwise.complete.obs", method = "spearman")),digits=3)

print(with(Descriptives, corr.test(HADSA, guidance, use = "pairwise.complete.obs", method = "spearman")), digits=3)
with(Descriptives, plot(HADSA, guidance))

print(with(Descriptives, corr.test(HADSA, adherence, use = "pairwise.complete.obs", method = "spearman")), digits=3)

print(with(Descriptives, corr.test(HADSA, avoid, use = "pairwise.complete.obs", method = "spearman")), digits=3)

print(with(Descriptives, corr.test(HADSA, leavehouse, use = "pairwise.complete.obs", method = "spearman")), digits=3)

print(with(Descriptives, corr.test(HADSA, help, use = "pairwise.complete.obs", method = "spearman")),digits=3)

print(with(Descriptives, corr.test(HADSA, internet, use = "pairwise.complete.obs", method = "spearman")), digits=3)

print(with(Descriptives, corr.test(HADSA, covidstress, use = "pairwise.complete.obs", method = "spearman")), digits=3)

print(with(Descriptives, corr.test(HADSA, mentalhealth, use = "pairwise.complete.obs", method = "spearman")), digits=3)

print(with(Descriptives, corr.test(HADSA, physicalhealth, use = "pairwise.complete.obs", method = "spearman")), digits=3)

print(with(Descriptives, corr.test(HADSA, alcohol, use = "pairwise.complete.obs", method = "spearman")), digits=3)

print(with(Descriptives, corr.test(HADSA, dietintake, use = "pairwise.complete.obs", method = "spearman")), digits=3)

print(with(Descriptives, corr.test(HADSA, healthydiet, use = "pairwise.complete.obs", method = "spearman")), digits=3)

print(with(Descriptives, corr.test(HADSA, physactivity, use = "pairwise.complete.obs", method = "spearman")), digits=3)


# HADS depression ---------------------------------------------------------

print(with(Descriptives, corr.test(HADSD, knowledge, use = "pairwise.complete.obs", method = "spearman")),digits=3)

print(with(Descriptives, corr.test(HADSD, guidance, use = "pairwise.complete.obs", method = "spearman")), digits=3)
with(Descriptives, plot(HADSD, guidance))

print(with(Descriptives, corr.test(HADSD, adherence, use = "pairwise.complete.obs", method = "spearman")), digits=3)

print(with(Descriptives, corr.test(HADSD, avoid, use = "pairwise.complete.obs", method = "spearman")), digits=3)

print(with(Descriptives, corr.test(HADSD, leavehouse, use = "pairwise.complete.obs", method = "spearman")), digits=3)

print(with(Descriptives, corr.test(HADSD, help, use = "pairwise.complete.obs", method = "spearman")),digits=3)

print(with(Descriptives, corr.test(HADSD, internet, use = "pairwise.complete.obs", method = "spearman")), digits=3)

print(with(Descriptives, corr.test(HADSD, covidstress, use = "pairwise.complete.obs", method = "spearman")), digits=3)

print(with(Descriptives, corr.test(HADSA, mentalhealth, use = "pairwise.complete.obs", method = "spearman")), digits=3)

print(with(Descriptives, corr.test(HADSD, physicalhealth, use = "pairwise.complete.obs", method = "spearman")), digits=3)

print(with(Descriptives, corr.test(HADSD, alcohol, use = "pairwise.complete.obs", method = "spearman")), digits=3)

print(with(Descriptives, corr.test(HADSD, dietintake, use = "pairwise.complete.obs", method = "spearman")), digits=3)

print(with(Descriptives, corr.test(HADSD, healthydiet, use = "pairwise.complete.obs", method = "spearman")), digits=3)

print(with(Descriptives, corr.test(HADSD, physactivity, use = "pairwise.complete.obs", method = "spearman")), digits=3)


# COVID-RELATED ANXIETY ---------------------------------------------------

print(with(Descriptives, corr.test(covidstress, knowledge, use = "pairwise.complete.obs", method = "spearman")),digits=3)

print(with(Descriptives, corr.test(covidstress, guidance, use = "pairwise.complete.obs", method = "spearman")), digits=3)

print(with(Descriptives, corr.test(covidstress, adherence, use = "pairwise.complete.obs", method = "spearman")), digits=3)

print(with(Descriptives, corr.test(covidstress, avoid, use = "pairwise.complete.obs", method = "spearman")), digits=3)

print(with(Descriptives, corr.test(covidstress, leavehouse, use = "pairwise.complete.obs", method = "spearman")), digits=3)

print(with(Descriptives, corr.test(covidstress, help, use = "pairwise.complete.obs", method = "spearman")),digits=3)

print(with(Descriptives, corr.test(covidstress, internet, use = "pairwise.complete.obs", method = "spearman")), digits=3)

print(with(Descriptives, corr.test(covidstress, covidstress, use = "pairwise.complete.obs", method = "spearman")), digits=3)

print(with(Descriptives, corr.test(covidstress, mentalhealth, use = "pairwise.complete.obs", method = "spearman")), digits=3)

print(with(Descriptives, corr.test(covidstress, physicalhealth, use = "pairwise.complete.obs", method = "spearman")), digits=3)

print(with(Descriptives, corr.test(HADSD, alcohol, use = "pairwise.complete.obs", method = "spearman")), digits=3)

print(with(Descriptives, corr.test(covidstress, dietintake, use = "pairwise.complete.obs", method = "spearman")), digits=3)

print(with(Descriptives, corr.test(covidstress, healthydiet, use = "pairwise.complete.obs", method = "spearman")), digits=3)

print(with(Descriptives, corr.test(covidstress, physactivity, use = "pairwise.complete.obs", method = "spearman")), digits=3)


# Emotional Stability -----------------------------------------------------

print(with(Descriptives, corr.test(emo, knowledge, use = "pairwise.complete.obs", method = "spearman")),digits=3)

print(with(Descriptives, corr.test(emo, guidance, use = "pairwise.complete.obs", method = "spearman")), digits=3)

print(with(Descriptives, corr.test(emo, adherence, use = "pairwise.complete.obs", method = "spearman")), digits=3)

print(with(Descriptives, corr.test(emo, avoid, use = "pairwise.complete.obs", method = "spearman")), digits=3)

print(with(Descriptives, corr.test(emo, leavehouse, use = "pairwise.complete.obs", method = "spearman")), digits=3)

print(with(Descriptives, corr.test(emo, help, use = "pairwise.complete.obs", method = "spearman")),digits=3)

print(with(Descriptives, corr.test(emo, internet, use = "pairwise.complete.obs", method = "spearman")), digits=3)

print(with(Descriptives, corr.test(emo, covidstress, use = "pairwise.complete.obs", method = "spearman")), digits=3)

print(with(Descriptives, corr.test(emo, mentalhealth, use = "pairwise.complete.obs", method = "spearman")), digits=3)

print(with(Descriptives, corr.test(emo, physicalhealth, use = "pairwise.complete.obs", method = "spearman")), digits=3)

print(with(Descriptives, corr.test(emo, alcohol, use = "pairwise.complete.obs", method = "spearman")), digits=3)

print(with(Descriptives, corr.test(emo, dietintake, use = "pairwise.complete.obs", method = "spearman")), digits=3)

print(with(Descriptives, corr.test(emo, healthydiet, use = "pairwise.complete.obs", method = "spearman")), digits=3)

print(with(Descriptives, corr.test(emo, physactivity, use = "pairwise.complete.obs", method = "spearman")), digits=3)


# Extraversion ------------------------------------------------------------

print(with(Descriptives, corr.test(ex, knowledge, use = "pairwise.complete.obs", method = "spearman")),digits=3)

print(with(Descriptives, corr.test(ex, guidance, use = "pairwise.complete.obs", method = "spearman")), digits=3)

print(with(Descriptives, corr.test(ex, adherence, use = "pairwise.complete.obs", method = "spearman")), digits=3)

print(with(Descriptives, corr.test(ex, avoid, use = "pairwise.complete.obs", method = "spearman")), digits=3)

print(with(Descriptives, corr.test(ex, leavehouse, use = "pairwise.complete.obs", method = "spearman")), digits=3)

print(with(Descriptives, corr.test(ex, help, use = "pairwise.complete.obs", method = "spearman")),digits=3)

print(with(Descriptives, corr.test(ex, internet, use = "pairwise.complete.obs", method = "spearman")), digits=3)

print(with(Descriptives, corr.test(ex, covidstress, use = "pairwise.complete.obs", method = "spearman")), digits=3)

print(with(Descriptives, corr.test(ex, mentalhealth, use = "pairwise.complete.obs", method = "spearman")), digits=3)

print(with(Descriptives, corr.test(ex, physicalhealth, use = "pairwise.complete.obs", method = "spearman")), digits=3)

print(with(Descriptives, corr.test(ex, alcohol, use = "pairwise.complete.obs", method = "spearman")), digits=3)

print(with(Descriptives, corr.test(ex, dietintake, use = "pairwise.complete.obs", method = "spearman")), digits=3)

print(with(Descriptives, corr.test(ex, healthydiet, use = "pairwise.complete.obs", method = "spearman")), digits=3)

print(with(Descriptives, corr.test(ex, physactivity, use = "pairwise.complete.obs", method = "spearman")), digits=3)

# Conscientiousness -------------------------------------------------------

print(with(Descriptives, corr.test(CON, knowledge, use = "pairwise.complete.obs", method = "spearman")),digits=3)

print(with(Descriptives, corr.test(CON, guidance, use = "pairwise.complete.obs", method = "spearman")), digits=3)

print(with(Descriptives, corr.test(CON, adherence, use = "pairwise.complete.obs", method = "spearman")), digits=3)

print(with(Descriptives, corr.test(CON, avoid, use = "pairwise.complete.obs", method = "spearman")), digits=3)

print(with(Descriptives, corr.test(CON, leavehouse, use = "pairwise.complete.obs", method = "spearman")), digits=3)

print(with(Descriptives, corr.test(CON, help, use = "pairwise.complete.obs", method = "spearman")),digits=3)

print(with(Descriptives, corr.test(CON, internet, use = "pairwise.complete.obs", method = "spearman")), digits=3)

print(with(Descriptives, corr.test(CON, covidstress, use = "pairwise.complete.obs", method = "spearman")), digits=3)

print(with(Descriptives, corr.test(CON, mentalhealth, use = "pairwise.complete.obs", method = "spearman")), digits=3)

print(with(Descriptives, corr.test(CON, physicalhealth, use = "pairwise.complete.obs", method = "spearman")), digits=3)

print(with(Descriptives, corr.test(CON, alcohol, use = "pairwise.complete.obs", method = "spearman")), digits=3)

print(with(Descriptives, corr.test(CON, dietintake, use = "pairwise.complete.obs", method = "spearman")), digits=3)

print(with(Descriptives, corr.test(CON, healthydiet, use = "pairwise.complete.obs", method = "spearman")), digits=3)

print(with(Descriptives, corr.test(CON, physactivity, use = "pairwise.complete.obs", method = "spearman")), digits=3)
# Coding check ------------------------------------------------------------------

summary(Descriptives$sex)
summary(Descriptives$pptclass)
summary(Descriptives$binarymarital)
summary(Descriptives$covidalone)
summary(Descriptives$geog)
summary(Descriptives$cvd)
summary(Descriptives$diab)
summary(Descriptives$hibp)
summary(Descriptives$srh)
summary(Descriptives$knowlege)
summary(Descriptives$CIQ_GetsHelp)
summary(Descriptives$CIQ_InternetUseChange)
summary(Descriptives$CIQ_LastTwoWeeks_Stress)


# Figure 1 (symptom duration) -------------------------------------------
Duration_Aches <- data.frame(subset(Descriptives$CIQ_DurationAches, (!is.na(Descriptives$CIQ_DurationAches))))
colnames(Duration_Aches) <- c("Duration")
Duration_Aches$Symptom <- c("Aches & Pains")
head(Duration_Aches)

Duration_ShortBreath <- data.frame(subset(Descriptives$CIQ_DurationShortBreath, (!is.na(Descriptives$CIQ_DurationShortBreath))))
colnames(Duration_ShortBreath) <- c("Duration")
Duration_ShortBreath$Symptom <- c("Shortness of \nBreath")
head(Duration_ShortBreath)

Duration_DryCough <- data.frame(subset(Descriptives$CIQ_DurationDryCough, (!is.na(Descriptives$CIQ_DurationDryCough))))
colnames(Duration_DryCough) <- c("Duration")
Duration_DryCough$Symptom <- c("Dry Cough")
head(Duration_DryCough)

Duration_Fatigue <- data.frame(subset(Descriptives$CIQ_DurationFatigue, (!is.na(Descriptives$CIQ_DurationFatigue))))
colnames(Duration_Fatigue) <- c("Duration")
Duration_Fatigue$Symptom <- c("Fatigue")
head(Duration_Fatigue)

Duration_RunnyNose <- data.frame(subset(Descriptives$CIQ_DurationRunnyNose, (!is.na(Descriptives$CIQ_DurationRunnyNose))))
colnames(Duration_RunnyNose) <- c("Duration")
Duration_RunnyNose$Symptom <- c("Runny Nose")
head(Duration_RunnyNose)

Duration_Sneezing <- data.frame(subset(Descriptives$CIQ_DurationSneezing, (!is.na(Descriptives$CIQ_DurationSneezing))))
colnames(Duration_Sneezing) <- c("Duration")
Duration_Sneezing$Symptom <- c("Sneezing")
head(Duration_Sneezing)

Duration_LossTaste <- data.frame(subset(Descriptives$CIQ_DurationLostTaste, (!is.na(Descriptives$CIQ_DurationLostTaste))))
colnames(Duration_LossTaste) <- c("Duration")
Duration_LossTaste$Symptom <- c("Loss of Taste")
head(Duration_LossTaste)

Duration_SkinLesions <- data.frame(subset(Descriptives$CIQ_DurationSkinLesions, (!is.na(Descriptives$CIQ_DurationSkinLesions))))
colnames(Duration_SkinLesions) <- c("Duration")
Duration_SkinLesions$Symptom <- c("Skin Lesions")
head(Duration_SkinLesions)

Duration_Symptoms <- rbind(Duration_Aches, Duration_DryCough, Duration_Fatigue, Duration_LossTaste, Duration_RunnyNose, Duration_ShortBreath, Duration_SkinLesions, Duration_Sneezing)
Duration_Symptoms$Duration <- factor(Duration_Symptoms$Duration, levels = c("A few days (up to one week)",
                                                                            "One to two weeks",
                                                                            "Two to three weeks",
                                                                            "Three to four weeks",
                                                                            "More than one month"))

ggplot(Duration_Symptoms, aes(x=Symptom, fill=(Duration)))+
    geom_bar(position="stack", stat = "count")+
    scale_fill_discrete(labels = c("A few days (up to 1 week)", "1-2 weeks", "2-3 weeks", "3-4 weeks", "More than 1 month"), drop=FALSE)+
    scale_x_discrete(drop=FALSE) + 
    theme_apa() + labs(y= "Frequency" , x = "COVID-19 Symptom", fill=guide_legend(title="Duration of Symptom"))


# Figure 2 (types of news source)  -------------------------------------------------

NewsSource <- data.frame(Descriptives$CIQ_SourceTV_BBC,
                         Descriptives$CIQ_SourceTV_OtherNews,
                         Descriptives$CIQ_Source_Radio,
                         Descriptives$CIQ_Source_Newspaper_Broadsheet,
                         Descriptives$CIQ_Source_Newspaper_Tabloid,
                         Descriptives$CIQ_SourceWeb_BBC,
                         Descriptives$CIQ_SourceWeb_OtherNews,
                         Descriptives$CIQ_Source_Web_NHS,
                         Descriptives$CIQ_Source_Web_GP,
                         Descriptives$CIQ_Source_Web_Government,
                         Descriptives$CIQ_Source_Web_WHO,
                         Descriptives$CIQ_Source_OtherHeathWebEtc,
                         Descriptives$CIQ_Source_SocialMedia,
                         Descriptives$CIQ_Source_Workplace,
                         Descriptives$CIQ_Source_FamAndFriends,
                         Descriptives$CIQ_Source_MessagingServices,
                         Descriptives$CIQ_Source_NoneOfAbove)
colnames(NewsSource) <- c("TV_BBC", 
                          "TV_OtherNews",
                          "Radio",
                          "Paper_Broadsheet",
                          "Paper_Tabloid",
                          "Web_BBC",
                          "Web_OtherNews",
                          "Web_NHS",
                          "Web_GP",
                          "Web_Govt",
                          "Web_WHO",
                          "Web_OtherHealth",
                          "SocialMedia",
                          "Workplace",
                          "FamFriends",
                          "Messaging",
                          "None")
NewsSource <- pivot_longer(data = NewsSource, names_to = "Source", 
                           cols = c(TV_BBC, 
                                    TV_OtherNews,
                                    Radio,
                                    Paper_Broadsheet,
                                    Paper_Tabloid,
                                    Web_BBC,
                                    Web_OtherNews,
                                    Web_NHS,
                                    Web_GP,
                                    Web_Govt,
                                    Web_WHO,
                                    Web_OtherHealth,
                                    SocialMedia,
                                    Workplace,
                                    FamFriends,
                                    Messaging,
                                    None))                   
                                                       
summary(NewsSource)
NewsSource <- subset(NewsSource, !is.na(NewsSource$value))
NewsSource$Source <- factor(NewsSource$Source, levels=c("TV_BBC", 
                                                        "TV_OtherNews",
                                                        "Radio",
                                                        "Paper_Broadsheet",
                                                        "Paper_Tabloid",
                                                        "Web_BBC",
                                                        "Web_OtherNews",
                                                        "Web_NHS",
                                                        "Web_GP",
                                                        "Web_Govt",
                                                        "Web_WHO",
                                                        "Web_OtherHealth",
                                                        "SocialMedia",
                                                        "Workplace",
                                                        "FamFriends",
                                                        "Messaging",
                                                        "None"))

ggplot(NewsSource, aes(x=Source)) + 
    geom_histogram(position="dodge", stat="count") +
    theme_apa() + scale_x_discrete(labels = c("BBC news",
                                              "Other television news bulletins (e.g., ITV, Channel 4)",
                                              "Radio or Podcasts",
                                              "Broadsheet newspapers",
                                              "Tabloid newspapers",
                                              "BBC news website",
                                              "Other news websites (e.g., ITV, Channel 4)",
                                              "NHS websites",
                                              "GP Practice Website",
                                              "Government websites",
                                              "WHO Website",
                                              "Other health websites & resources",
                                              "Social media",
                                              "Workplace",
                                              "Family and Friends",
                                              "WhatsApp or other messaging services",
                                              "None of the above"), drop=FALSE) + 
    scale_y_continuous(breaks = c(0,25,50,75,100,125,150, 175))+
    labs(y= "Frequency" , x = "Source of News") + coord_flip()

# Figure 3 (most helpful news source) ------------------------------------------

BestNewsSource <- data.frame(Descriptives$CIQ_BestSource_TV_BBC,
                             Descriptives$CIQ_BestSource_TV_OtherNews,
                             Descriptives$CIQ_BestSource_Radio,
                             Descriptives$CIQ_BestSource_Newspaper_Broadsheet,
                             Descriptives$CIQ_BestSource_Newspaper_Tabloid,
                             Descriptives$CIQ_BestSource_Web_BBC,
                             Descriptives$CIQ_BestSource_Web_OtherNews,
                             Descriptives$CIQ_BestSource_Web_NHS,
                             Descriptives$CIQ_BestSource_Web_GP,
                             Descriptives$CIQ_BestSource_Web_Government,
                             Descriptives$CIQ_BestSource_Web_WHO,
                             Descriptives$CIQ_BestSource_OtherHealthWebEtc,
                             Descriptives$CIQ_BestSource_SocialMedia,
                             Descriptives$CIQ_BestSource_Workplace,
                             Descriptives$CIQ_BestSource_FamAndFriends,
                             Descriptives$CIQ_BestSource_MessagingServices)
colnames(BestNewsSource) <- c("TV_BBC", 
                          "TV_OtherNews",
                          "Radio",
                          "Paper_Broadsheet",
                          "Paper_Tabloid",
                          "Web_BBC",
                          "Web_OtherNews",
                          "Web_NHS",
                          "Web_GP",
                          "Web_Govt",
                          "Web_WHO",
                          "Web_OtherHealth",
                          "SocialMedia",
                          "Workplace",
                          "FamFriends",
                          "Messaging")
BestNewsSource <- pivot_longer(data = BestNewsSource, names_to = "Source", 
                           cols = c(TV_BBC, 
                                    TV_OtherNews,
                                    Radio,
                                    Paper_Broadsheet,
                                    Paper_Tabloid,
                                    Web_BBC,
                                    Web_OtherNews, 
                                    Web_NHS,
                                    Web_GP,
                                    Web_Govt,
                                    Web_WHO,
                                    Web_OtherHealth,
                                    SocialMedia,
                                    Workplace,
                                    FamFriends,
                                    Messaging))                   

summary(BestNewsSource)
BestNewsSource <- subset(BestNewsSource, !is.na(BestNewsSource$value))
BestNewsSource$Source <- factor(BestNewsSource$Source, levels=c("TV_BBC", 
                                                        "TV_OtherNews",
                                                        "Radio",
                                                        "Paper_Broadsheet",
                                                        "Paper_Tabloid",
                                                        "Web_BBC",
                                                        "Web_OtherNews",
                                                        "Web_NHS",
                                                        "Web_GP",
                                                        "Web_Govt",
                                                        "Web_WHO",
                                                        "Web_OtherHealth",
                                                        "SocialMedia",
                                                        "Workplace",
                                                        "FamFriends",
                                                        "Messaging"))

ggplot(BestNewsSource, aes(x=Source)) + 
    geom_histogram(position="dodge", stat="count") +
    theme_apa() + scale_x_discrete(labels = c("BBC news",
                                              "Other television news bulletins (e.g., ITV, Channel 4)",
                                              "Radio or Podcasts",
                                              "Broadsheet newspapers",
                                              "Tabloid newspapers",
                                              "BBC news website",
                                              "Other news websites (e.g., ITV, Channel 4)",
                                              "NHS websites",
                                              "GP Practice Website",
                                              "Government websites",
                                              "WHO Website",
                                              "Other health websites & resources",
                                              "Social media",
                                              "Workplace",
                                              "Family and Friends",
                                              "WhatsApp or other messaging services",
                                              "None of the above"), drop=FALSE) + 
    scale_y_continuous(breaks = c(0,25,50,75,100,125,150, 175))+
    labs(y= "Frequency" , x = "Best Source of News") + coord_flip()

# Figure 4 (type of local initiative)  -----------------------------------------------

Descriptives$CIQ_LocalInitiatives


# Figure 5 (contact with family) ----------------------------------------------

#CALLING
Contact_Family_Call_Now <- data.frame(subset(Descriptives$CIQ_NowFreq_CallFamily, 
                                             (!is.na(Descriptives$CIQ_NowFreq_CallFamily))))
Contact_Family_Call_Now$Method <- c("Call_Now")
colnames(Contact_Family_Call_Now) <- c("Freq", "Method")
summary(Contact_Family_Call_Now) 

Contact_Family_Call_Prev <- data.frame(subset(Descriptives$CIQ_PriorFreq_CallFamily, 
                                              (!is.na(Descriptives$CIQ_PriorFreq_CallFamily))))
Contact_Family_Call_Prev$Method <- c("Call_Prev")
colnames(Contact_Family_Call_Prev) <- c("Freq", "Method")
summary(Contact_Family_Call_Prev) 

#MEETING
Contact_Family_Meet_Now <- data.frame(subset(Descriptives$CIQ_NowFreq_MeetFamily, 
                                             (!is.na(Descriptives$CIQ_NowFreq_MeetFamily))))
Contact_Family_Meet_Now$Method <- c("Meet_Now")
colnames(Contact_Family_Meet_Now) <- c("Freq", "Method")
head(Contact_Family_Meet_Now) 

Contact_Family_Meet_Prev <- data.frame(subset(Descriptives$CIQ_PriorFreq_MeetFamily, 
                                              (!is.na(Descriptives$CIQ_PriorFreq_MeetFamily))))
Contact_Family_Meet_Prev$Method <- c("Meet_Prev")
colnames(Contact_Family_Meet_Prev) <- c("Freq", "Method")
head(Contact_Family_Meet_Prev) 

#MESSAGE
Contact_Family_Msg_Now <- data.frame(subset(Descriptives$CIQ_NowFreq_MsgFamily, 
                                            (!is.na(Descriptives$CIQ_NowFreq_MsgFamily))))
Contact_Family_Msg_Now$Method <- c("Msg_Now")
colnames(Contact_Family_Msg_Now) <- c("Freq", "Method")
head(Contact_Family_Msg_Now) 

Contact_Family_Msg_Prev <- data.frame(subset(Descriptives$CIQ_PriorFreq_MsgFamily, 
                                             (!is.na(Descriptives$CIQ_PriorFreq_MsgFamily))))
Contact_Family_Msg_Prev$Method <- c("Msg_Prev")
colnames(Contact_Family_Msg_Prev) <- c("Freq", "Method")
head(Contact_Family_Msg_Prev) 

#VIDEO CALL
Contact_Family_Vid_Now <- data.frame(subset(Descriptives$CIQ_NowFreq_VidCall_Family, 
                                            (!is.na(Descriptives$CIQ_NowFreq_VidCall_Family))))
Contact_Family_Vid_Now$Method <- c("Vid_Now")
colnames(Contact_Family_Vid_Now) <- c("Freq", "Method")
head(Contact_Family_Vid_Now) 

Contact_Family_Vid_Prev <- data.frame(subset(Descriptives$CIQ_PriorFreq_VidCall_Family, 
                                             (!is.na(Descriptives$CIQ_PriorFreq_VidCall_Family))))
Contact_Family_Vid_Prev$Method <- c("Vid_Prev")
colnames(Contact_Family_Vid_Prev) <- c("Freq", "Method")
head(Contact_Family_Vid_Prev) 

Contact_Family <- rbind(Contact_Family_Call_Prev, Contact_Family_Call_Now,
                        Contact_Family_Meet_Prev, Contact_Family_Meet_Now,
                        Contact_Family_Msg_Prev, Contact_Family_Msg_Now,
                        Contact_Family_Vid_Prev, Contact_Family_Vid_Now)
Contact_Family$Method <- factor(Contact_Family$Method)
Contact_Family$Freq <- factor(Contact_Family$Freq, levels=c("Never","Rarely","Less than once a week","1-2 days a week","3-4 days a week","Every day/almost every day"))
summary(Contact_Family$Freq)

ggplot(Contact_Family, aes(fill=Freq, x=Method)) + 
    geom_histogram(position="stack", stat="count") +
    coord_flip() + theme_apa() + scale_x_discrete(labels = c("Now \n Phone call",
                                                             "Previously \n Phone call",
                                                             "Now \n Meet in person",
                                                             "Previously \n Meet in person",
                                                             "Now \n Text or \n instant message",
                                                             "Previously \n Text or \n instant message",
                                                             "Now \n Video Call",
                                                             "Previously \n Video Call")) + 
    scale_y_continuous(breaks = c(0,25,50,75,100,125,150,175))+
    labs(y= "Frequency" , x = "Method of Contact")  
 


# Figure 6 (contact with friends) ----------------------------------------------

#CALLING
Contact_Friends_Call_Now <- data.frame(subset(Descriptives$CIQ_NowFreq_CallFriends, 
                                             (!is.na(Descriptives$CIQ_NowFreq_CallFriends))))
Contact_Friends_Call_Now$Method <- c("Call_Now")
colnames(Contact_Friends_Call_Now) <- c("Freq", "Method")
summary(Contact_Friends_Call_Now) 

Contact_Friends_Call_Prev <- data.frame(subset(Descriptives$CIQ_PriorFreq_CallFriends, 
                                              (!is.na(Descriptives$CIQ_PriorFreq_CallFriends))))
Contact_Friends_Call_Prev$Method <- c("Call_Prev")
colnames(Contact_Friends_Call_Prev) <- c("Freq", "Method")
summary(Contact_Friends_Call_Prev) 

#MEETING
Contact_Friends_Meet_Now <- data.frame(subset(Descriptives$CIQ_NowFreq_MeetFriends, 
                                             (!is.na(Descriptives$CIQ_NowFreq_MeetFriends))))
Contact_Friends_Meet_Now$Method <- c("Meet_Now")
colnames(Contact_Friends_Meet_Now) <- c("Freq", "Method")
head(Contact_Friends_Meet_Now) 

Contact_Friends_Meet_Prev <- data.frame(subset(Descriptives$CIQ_PriorFreq_MeetFriends, 
                                              (!is.na(Descriptives$CIQ_PriorFreq_MeetFriends))))
Contact_Friends_Meet_Prev$Method <- c("Meet_Prev")
colnames(Contact_Friends_Meet_Prev) <- c("Freq", "Method")
head(Contact_Friends_Meet_Prev) 

#MESSAGE
Contact_Friends_Msg_Now <- data.frame(subset(Descriptives$CIQ_NowFreq_MsgFriends, 
                                            (!is.na(Descriptives$CIQ_NowFreq_MsgFriends))))
Contact_Friends_Msg_Now$Method <- c("Msg_Now")
colnames(Contact_Friends_Msg_Now) <- c("Freq", "Method")
head(Contact_Friends_Msg_Now) 

Contact_Friends_Msg_Prev <- data.frame(subset(Descriptives$CIQ_PriorFreq_MsgFriends, 
                                             (!is.na(Descriptives$CIQ_PriorFreq_MsgFriends))))
Contact_Friends_Msg_Prev$Method <- c("Msg_Prev")
colnames(Contact_Friends_Msg_Prev) <- c("Freq", "Method")
head(Contact_Friends_Msg_Prev) 

#VIDEO CALL
Contact_Friends_Vid_Now <- data.frame(subset(Descriptives$CIQ_NowFreq_VidCall_Friends, 
                                            (!is.na(Descriptives$CIQ_NowFreq_VidCall_Friends))))
Contact_Friends_Vid_Now$Method <- c("Vid_Now")
colnames(Contact_Friends_Vid_Now) <- c("Freq", "Method")
head(Contact_Friends_Vid_Now) 

Contact_Friends_Vid_Prev <- data.frame(subset(Descriptives$CIQ_PriorFreq_VidCall_Friends, 
                                             (!is.na(Descriptives$CIQ_PriorFreq_VidCall_Friends))))
Contact_Friends_Vid_Prev$Method <- c("Vid_Prev")
colnames(Contact_Friends_Vid_Prev) <- c("Freq", "Method")
head(Contact_Friends_Vid_Prev) 

Contact_Friends <- rbind(Contact_Friends_Call_Prev, Contact_Friends_Call_Now,
                        Contact_Friends_Meet_Prev, Contact_Friends_Meet_Now,
                        Contact_Friends_Msg_Prev, Contact_Friends_Msg_Now,
                        Contact_Friends_Vid_Prev, Contact_Friends_Vid_Now)
Contact_Friends$Method <- factor(Contact_Friends$Method)
Contact_Friends$Freq <- factor(Contact_Friends$Freq, levels=c("Never","Rarely","Less than once a week","1-2 days a week","3-4 days a week","Every day/almost every day"))
summary(Contact_Friends$Freq)

ggplot(Contact_Friends, aes(fill=Freq, x=Method)) + 
    geom_histogram(position="stack", stat="count") +
    coord_flip() + theme_apa() + scale_x_discrete(labels = c("Now \n Phone call",
                                                             "Previously \n Phone call",
                                                             "Now \n Meet in person",
                                                             "Previously \n Meet in person",
                                                             "Now \n Text or \n instant message",
                                                             "Previously \n Text or \n instant message",
                                                             "Now \n Video Call",
                                                             "Previously \n Video Call")) + 
    scale_y_continuous(breaks = c(0,25,50,75,100,125,150,175))+
    labs(y= "Frequency" , x = "Method of Contact")  


# Figure 7 (self-reported physical health)  -----------------------------------------

#PHYSICAL HEALTH CHANGE
PhysHealthChange <- data.frame(summary(Descriptives$CIQ_BeforeMarch23_PhysicalHealth), summary(Descriptives$CIQ_SinceMarch23_PhysicalHealth))
PhysHealthChange <- subset(PhysHealthChange[1:5,1:2])
colnames(PhysHealthChange) <- c("BeforePhysHealth", "SincePhysHealth")
PhysHealthChange <- pivot_longer(PhysHealthChange, c(BeforePhysHealth, SincePhysHealth))
PhysHealthChange[ ,3] <- c("Excellent", "Excellent", "Very Good", "Very Good", "Good", "Good", "Fair", "Fair", "Poor", "Poor")
PhysHealthChange[ ,1] <- c("Before lockdown", "Since lockdown") 
colnames(PhysHealthChange) <- c("BeforeSince", "Freq", "Rating")
PhysHealthChange$Freq <- as.numeric(PhysHealthChange$Freq)
PhysHealthChange$Rating <- factor(PhysHealthChange$Rating, 
                                  levels = c("Excellent", "Very Good", "Good", "Fair", "Poor"))
PhysHealthChange
levels(PhysHealthChange$Rating)

#STACKED BAR PLOT
PhysHealthChange %>% 
    ggplot(aes(x = Rating, y = Freq, fill = BeforeSince))+
    geom_bar(stat = "identity", position = 'dodge')+ 
    theme_apa() + 
    scale_x_discrete(labels = c("Excellent", "Very good", 'Good', 'Fair', 'Poor')) + 
    scale_fill_discrete()+
    scale_y_continuous(breaks = c(0,25,50,75,100,125,150,175))+
    labs(y="Frequency", x="Self-reported physical health") +
    scale_fill_grey(
        start = 0.2,
        end = 0.6,
        na.value = "red",
        aesthetics = "fill")


#DIVERGENT BAR PLOT
PhysHealthChange <- PhysHealthChange %>% 
    mutate(Freq = ifelse(BeforeSince == "Since", Freq, -1*Freq))

Rating_Values <- c(-100,-75,-50,-25,0,25,50,75,100)
PhysHealthChange %>%
    ggplot(aes(x = Rating, y = Freq, fill = BeforeSince))+
    geom_bar(stat = "identity")+ coord_flip() + theme_apa(remove.x.gridlines = F) + 
    scale_x_discrete() + scale_fill_discrete(name="Time Period", labels = c("Before COVID-19\n Measures", "After COVID-19\n Measures"))+
    scale_y_continuous(breaks = Rating_Values,labels = abs(Rating_Values))+
    labs(y="Frequency", x="Self-rated general physical health ")


# Figure 8 (self-reported emotional and mental health)  ------------------------------------------------------------------

#MENTAL HEALTH CHANGE
MentalHealthChange <- data.frame(summary(Descriptives$CIQ_BeforeMarch23_MentalHealth), summary(Descriptives$CIQ_SinceMarch23_MentaHealth))
MentalHealthChange <- subset(MentalHealthChange[1:5,1:2])
colnames(MentalHealthChange) <- c("BeforeMentalHealth", "SinceMentalHealth")
MentalHealthChange <- pivot_longer(MentalHealthChange, c(BeforeMentalHealth, SinceMentalHealth))
MentalHealthChange[ ,3] <- c("Excellent", "Excellent", "Very Good", "Very Good", "Good", "Good", "Fair", "Fair", "Poor", "Poor")
MentalHealthChange[ ,1] <- c("Before lockdown", "Since lockdown") 
colnames(MentalHealthChange) <- c("BeforeSince", "Freq", "Rating")
MentalHealthChange$Freq <- as.numeric(MentalHealthChange$Freq)
MentalHealthChange$Rating <- factor(MentalHealthChange$Rating, 
                                    levels = c("Excellent", "Very Good", "Good", "Fair", "Poor"))
MentalHealthChange
levels(MentalHealthChange$Rating)

#STACKED BAR PLOT
MentalHealthChange %>% 
    ggplot(aes(x = Rating, y = Freq, fill = BeforeSince))+
    geom_bar(stat = "identity", position = 'dodge')+ 
    theme_apa() + 
    scale_x_discrete(labels = c("Excellent", "Very good", 'Good', 'Fair', 'Poor')) + 
    scale_fill_discrete()+
    scale_y_continuous(breaks = c(0,25,50,75,100,125,150,175))+
    labs(y="Frequency", x="Self-reported mental health")+
    scale_fill_grey(
        start = 0.2,
        end = 0.6,
        na.value = "red",
        aesthetics = "fill")

#DIVERGENT BAR PLOT
MentalHealthChange <- MentalHealthChange %>%
    mutate(Freq = ifelse(BeforeSince == "Since", Freq, -1*Freq))

MentalHealthChange %>%
    ggplot(aes(x = Rating, y = Freq, fill = BeforeSince))+
    geom_bar(stat = "identity")+ coord_flip() + theme_apa(remove.x.gridlines = F) + 
    scale_x_discrete() + scale_fill_discrete(name="Time Period", labels = c("Before COVID-19\n Measures", "After COVID-19\n Measures"))+
    scale_y_continuous(breaks = Rating_Values,labels = abs(Rating_Values))+
    labs(y="Frequency", x="Self-rated general emotional and mental health")


# Figure 9 (pastimes returned to or taken up during lockdown) -------------

Pasttime <- data.frame(Descriptives$CIQ_Pastime_Exercise,
                       Descriptives$CIQ_Pastime_Relax,
                       Descriptives$CIQ_Pastime_Dancing,
                       Descriptives$CIQ_Pastime_Sing,
                       Descriptives$CIQ_Pastime_PlayingMusic,
                       Descriptives$CIQ_Pastime_ListeningMusic,
                       Descriptives$CIQ_Pastime_Writing,
                       Descriptives$CIQ_Pastime_Cooking,
                       Descriptives$CIQ_Pastime_Baking,
                       Descriptives$CIQ_Pastime_ArtsCrafts,
                       Descriptives$CIQ_Pastime_Gardening,
                       Descriptives$CIQ_Pastime_Reading,
                       Descriptives$CIQ_Pastime_OnlineLearning,
                       Descriptives$CIQ_Pastime_Language,
                       Descriptives$CIQ_Pastime_TvFilms,
                       Descriptives$CIQ_Pastime_RadioAudioBks,
                       Descriptives$CIQ_Pastime_BoardGames,
                       Descriptives$CIQ_Pastime_OnlineGames,
                       Descriptives$CIQ_Pastime_OtherActivity,
                       Descriptives$CIQ_Pastime_NoChange)
colnames(Pasttime) <- c("Exercise", 
                        "Relax",
                        "Dancing",
                        "Singing",
                        "PlayingMusic",
                        "ListeningMusic",
                        "Writing",
                        "Cooking",
                        "Baking",
                        "ArtsCrafts",
                        "Gardening",
                        "Reading",
                        "OnlineLearning",
                        "LearningLanguage",
                        "TVFilms",
                        "RadioAudiobooks",
                        "BoardGames",
                        "OnlineGames",
                        "OtherActivity",
                        "NoChange")
Pasttime <- pivot_longer(data = Pasttime, names_to = "Activity",
                         cols = c(Exercise, 
                                  Relax,
                                  Dancing,
                                  Singing,
                                  PlayingMusic,
                                  ListeningMusic,
                                  Writing,
                                  Cooking,
                                  Baking,
                                  ArtsCrafts,
                                  Gardening,
                                  Reading,
                                  OnlineLearning,
                                  LearningLanguage,
                                  TVFilms,
                                  RadioAudiobooks,
                                  BoardGames,
                                  OnlineGames,
                                  OtherActivity,
                                  NoChange))                   

summary(Pasttime)
Pasttime <- subset(Pasttime, !is.na(Pasttime$value))
Pasttime$Activity <- factor(Pasttime$Activity, levels=c("Exercise", 
                                                        "Relax",
                                                        "Dancing",
                                                        "Singing",
                                                        "PlayingMusic",
                                                        "ListeningMusic",
                                                        "Writing",
                                                        "Cooking",
                                                        "Baking",
                                                        "ArtsCrafts",
                                                        "Gardening",
                                                        "Reading",
                                                        "OnlineLearning",
                                                        "LearningLanguage",
                                                        "TVFilms",
                                                        "RadioAudiobooks",
                                                        "BoardGames",
                                                        "OnlineGames",
                                                        "OtherActivity",
                                                        "NoChange"))

ggplot(Pasttime, aes(x=Activity)) +
    geom_histogram(position="dodge", stat="count") +
    theme_apa() + scale_fill_discrete(drop=FALSE) + scale_x_discrete(drop=TRUE,
                                                                     labels=c("Work out or exercise",
                                                                              "Relaxation or meditation",
                                                                              "Dancing",
                                                                              "Playing music",
                                                                              "Listening to music",
                                                                              "Writing for pleasure",
                                                                              "Cooking class/learning to cook",
                                                                              "Baking",
                                                                              "Arts and crafts",
                                                                              "Gardening",
                                                                              "Reading",
                                                                              "Online educational course",
                                                                              "Learning a language",
                                                                              "Watching television/films",
                                                                              "Listening to the radio/audiobooks",
                                                                              "Board or card games",
                                                                              "Online gaming",
                                                                              "Other activity not listed above",
                                                                              "No change in activities"))+
    labs(y="Frequency", x="Pasttime started or returned to since COVID-19 measures in place")+
    scale_y_continuous(breaks=c(0,10,20,30,40,50,60,70))+coord_flip()


# Correlations (selected predictors+outcomes) -----------------------------

#Age (days at CIQ)
print(with(Descriptives, corr.test(agedays_CIQ, as.numeric(binaryinternet),
                                   use = "pairwise.complete.obs", method = "spearman")), digits=3)
print(with(Descriptives, corr.test(agedays_CIQ, as.numeric(binarynervous),
                                   use = "pairwise.complete.obs", method = "spearman")), digits=3)
print(with(Descriptives, corr.test(agedays_CIQ, as.numeric(getshelp),
                                   use = "pairwise.complete.obs", method = "spearman")), digits=3)
print(with(Descriptives, corr.test(agedays_CIQ, as.numeric(pasttime),
                                   use = "pairwise.complete.obs", method = "spearman")), digits=3)
print(with(Descriptives, corr.test(agedays_CIQ, as.numeric(lonely),
                                   use = "pairwise.complete.obs", method = "spearman")), digits=3)
print(with(Descriptives, corr.test(agedays_CIQ, as.numeric(changephys),
                                   use = "pairwise.complete.obs", method = "spearman")), digits=3)
print(with(Descriptives, corr.test(agedays_CIQ, as.numeric(routine),
                                   use = "pairwise.complete.obs", method = "spearman")), digits=3)
print(with(Descriptives, corr.test(agedays_CIQ, as.numeric(leavehouse),
                                   use = "pairwise.complete.obs", method = "spearman")), digits=3)
print(with(Descriptives, corr.test(agedays_CIQ, as.numeric(physicalhealth),
                                   use = "pairwise.complete.obs", method = "spearman")), digits=3)
print(with(Descriptives, corr.test(agedays_CIQ, as.numeric(mentalhealth),
                                   use = "pairwise.complete.obs", method = "spearman")), digits=3)

#Sex
print(with(Descriptives, corr.test(sex, as.numeric(binaryinternet),
                                   use = "pairwise.complete.obs", method = "spearman")), digits=3)
print(with(Descriptives, corr.test(sex, as.numeric(binarynervous),
                                   use = "pairwise.complete.obs", method = "spearman")), digits=3)
print(with(Descriptives, corr.test(sex, as.numeric(getshelp),
                                   use = "pairwise.complete.obs", method = "spearman")), digits=3)
print(with(Descriptives, corr.test(sex, as.numeric(pasttime),
                                   use = "pairwise.complete.obs", method = "spearman")), digits=3)
print(with(Descriptives, corr.test(sex, as.numeric(lonely),
                                   use = "pairwise.complete.obs", method = "spearman")), digits=3)
print(with(Descriptives, corr.test(sex, as.numeric(changephys),
                                   use = "pairwise.complete.obs", method = "spearman")), digits=3)
print(with(Descriptives, corr.test(sex, as.numeric(routine),
                                   use = "pairwise.complete.obs", method = "spearman")), digits=3)
print(with(Descriptives, corr.test(sex, as.numeric(leavehouse),
                                   use = "pairwise.complete.obs", method = "spearman")), digits=3)
print(with(Descriptives, corr.test(sex, as.numeric(physicalhealth),
                                   use = "pairwise.complete.obs", method = "spearman")), digits=3)
print(with(Descriptives, corr.test(sex, as.numeric(mentalhealth),
                                   use = "pairwise.complete.obs", method = "spearman")), digits=3)

#Adult SES
print(with(Descriptives, corr.test(pptclass, as.numeric(binaryinternet),
                                   use = "pairwise.complete.obs", method = "spearman")), digits=3)
print(with(Descriptives, corr.test(pptclass, as.numeric(binarynervous),
                                   use = "pairwise.complete.obs", method = "spearman")), digits=3)
print(with(Descriptives, corr.test(pptclass, as.numeric(getshelp),
                                   use = "pairwise.complete.obs", method = "spearman")), digits=3)
print(with(Descriptives, corr.test(pptclass, as.numeric(pasttime),
                                   use = "pairwise.complete.obs", method = "spearman")), digits=3)
print(with(Descriptives, corr.test(pptclass, as.numeric(lonely),
                                   use = "pairwise.complete.obs", method = "spearman")), digits=3)
print(with(Descriptives, corr.test(pptclass, as.numeric(changephys),
                                   use = "pairwise.complete.obs", method = "spearman")), digits=3)
print(with(Descriptives, corr.test(pptclass, as.numeric(routine),
                                   use = "pairwise.complete.obs", method = "spearman")), digits=3)
print(with(Descriptives, corr.test(pptclass, as.numeric(leavehouse),
                                   use = "pairwise.complete.obs", method = "spearman")), digits=3)
print(with(Descriptives, corr.test(pptclass, as.numeric(physicalhealth),
                                   use = "pairwise.complete.obs", method = "spearman")), digits=3)
print(with(Descriptives, corr.test(pptclass, as.numeric(mentalhealth),
                                   use = "pairwise.complete.obs", method = "spearman")), digits=3)

#Living alone
print(with(Descriptives, corr.test(covidalone, as.numeric(binaryinternet),
                                   use = "pairwise.complete.obs", method = "spearman")), digits=3)
print(with(Descriptives, corr.test(covidalone, as.numeric(binarynervous),
                                   use = "pairwise.complete.obs", method = "spearman")), digits=3)
print(with(Descriptives, corr.test(covidalone, as.numeric(getshelp),
                                   use = "pairwise.complete.obs", method = "spearman")), digits=3)
print(with(Descriptives, corr.test(covidalone, as.numeric(pasttime),
                                   use = "pairwise.complete.obs", method = "spearman")), digits=3)
print(with(Descriptives, corr.test(covidalone, as.numeric(lonely),
                                   use = "pairwise.complete.obs", method = "spearman")), digits=8)
print(with(Descriptives, corr.test(covidalone, as.numeric(changephys),
                                   use = "pairwise.complete.obs", method = "spearman")), digits=3)
print(with(Descriptives, corr.test(covidalone, as.numeric(routine),
                                   use = "pairwise.complete.obs", method = "spearman")), digits=3)
print(with(Descriptives, corr.test(covidalone, as.numeric(leavehouse),
                                   use = "pairwise.complete.obs", method = "spearman")), digits=3)
print(with(Descriptives, corr.test(covidalone, as.numeric(physicalhealth),
                                   use = "pairwise.complete.obs", method = "spearman")), digits=3)
print(with(Descriptives, corr.test(covidalone, as.numeric(mentalhealth),
                                   use = "pairwise.complete.obs", method = "spearman")), digits=3)

#general intelligence (g)
print(with(Descriptives, corr.test(g, as.numeric(binaryinternet),
                                   use = "pairwise.complete.obs", method = "spearman")), digits=3)
print(with(Descriptives, corr.test(g, as.numeric(binarynervous),
                                   use = "pairwise.complete.obs", method = "spearman")), digits=3)
print(with(Descriptives, corr.test(g, as.numeric(getshelp),
                                   use = "pairwise.complete.obs", method = "spearman")), digits=3)
print(with(Descriptives, corr.test(g, as.numeric(pasttime),
                                   use = "pairwise.complete.obs", method = "spearman")), digits=3)
print(with(Descriptives, corr.test(g, as.numeric(lonely),
                                   use = "pairwise.complete.obs", method = "spearman")), digits=3)
print(with(Descriptives, corr.test(g, as.numeric(changephys),
                                   use = "pairwise.complete.obs", method = "spearman")), digits=4)
print(with(Descriptives, corr.test(g, as.numeric(routine),
                                   use = "pairwise.complete.obs", method = "spearman")), digits=3)
print(with(Descriptives, corr.test(g, as.numeric(leavehouse),
                                   use = "pairwise.complete.obs", method = "spearman")), digits=3)
print(with(Descriptives, corr.test(g, as.numeric(physicalhealth),
                                   use = "pairwise.complete.obs", method = "spearman")), digits=5)
print(with(Descriptives, corr.test(g, as.numeric(mentalhealth),
                                   use = "pairwise.complete.obs", method = "spearman")), digits=3)

#General health literacy
print(with(Descriptives, corr.test(GHL, as.numeric(binaryinternet),
                                   use = "pairwise.complete.obs", method = "spearman")), digits=3)
print(with(Descriptives, corr.test(GHL, as.numeric(binarynervous),
                                   use = "pairwise.complete.obs", method = "spearman")), digits=3)
print(with(Descriptives, corr.test(GHL, as.numeric(getshelp),
                                   use = "pairwise.complete.obs", method = "spearman")), digits=3)
print(with(Descriptives, corr.test(GHL, as.numeric(pasttime),
                                   use = "pairwise.complete.obs", method = "spearman")), digits=3)
print(with(Descriptives, corr.test(GHL, as.numeric(lonely),
                                   use = "pairwise.complete.obs", method = "spearman")), digits=3)
print(with(Descriptives, corr.test(GHL, as.numeric(changephys),
                                   use = "pairwise.complete.obs", method = "spearman")), digits=3)
print(with(Descriptives, corr.test(GHL, as.numeric(routine),
                                   use = "pairwise.complete.obs", method = "spearman")), digits=3)
print(with(Descriptives, corr.test(GHL, as.numeric(leavehouse),
                                   use = "pairwise.complete.obs", method = "spearman")), digits=3)
print(with(Descriptives, corr.test(GHL, as.numeric(physicalhealth),
                                   use = "pairwise.complete.obs", method = "spearman")), digits=3)
print(with(Descriptives, corr.test(GHL, as.numeric(mentalhealth),
                                   use = "pairwise.complete.obs", method = "spearman")), digits=3)

#Chronic diseases
print(with(Descriptives, corr.test(disease, as.numeric(binaryinternet),
                                   use = "pairwise.complete.obs", method = "spearman")), digits=3)
print(with(Descriptives, corr.test(disease, as.numeric(binarynervous),
                                   use = "pairwise.complete.obs", method = "spearman")), digits=3)
print(with(Descriptives, corr.test(disease, as.numeric(getshelp),
                                   use = "pairwise.complete.obs", method = "spearman")), digits=3)
print(with(Descriptives, corr.test(disease, as.numeric(pasttime),
                                   use = "pairwise.complete.obs", method = "spearman")), digits=3)
print(with(Descriptives, corr.test(disease, as.numeric(lonely),
                                   use = "pairwise.complete.obs", method = "spearman")), digits=3)
print(with(Descriptives, corr.test(disease, as.numeric(changephys),
                                   use = "pairwise.complete.obs", method = "spearman")), digits=3)
print(with(Descriptives, corr.test(disease, as.numeric(routine),
                                   use = "pairwise.complete.obs", method = "spearman")), digits=3)
print(with(Descriptives, corr.test(disease, as.numeric(leavehouse),
                                   use = "pairwise.complete.obs", method = "spearman")), digits=3)
print(with(Descriptives, corr.test(disease, as.numeric(physicalhealth),
                                   use = "pairwise.complete.obs", method = "spearman")), digits=3)
print(with(Descriptives, corr.test(disease, as.numeric(mentalhealth),
                                   use = "pairwise.complete.obs", method = "spearman")), digits=3)

#Townsend score
print(with(Descriptives, corr.test(townsend, as.numeric(binaryinternet),
                                   use = "pairwise.complete.obs", method = "spearman")), digits=3)
print(with(Descriptives, corr.test(townsend, as.numeric(binarynervous),
                                   use = "pairwise.complete.obs", method = "spearman")), digits=3)
print(with(Descriptives, corr.test(townsend, as.numeric(getshelp),
                                   use = "pairwise.complete.obs", method = "spearman")), digits=3)
print(with(Descriptives, corr.test(townsend, as.numeric(pasttime),
                                   use = "pairwise.complete.obs", method = "spearman")), digits=3)
print(with(Descriptives, corr.test(townsend, as.numeric(lonely),
                                   use = "pairwise.complete.obs", method = "spearman")), digits=3)
print(with(Descriptives, corr.test(townsend, as.numeric(changephys),
                                   use = "pairwise.complete.obs", method = "spearman")), digits=3)
print(with(Descriptives, corr.test(townsend, as.numeric(routine),
                                   use = "pairwise.complete.obs", method = "spearman")), digits=3)
print(with(Descriptives, corr.test(townsend, as.numeric(leavehouse),
                                   use = "pairwise.complete.obs", method = "spearman")), digits=3)
print(with(Descriptives, corr.test(townsend, as.numeric(physicalhealth),
                                   use = "pairwise.complete.obs", method = "spearman")), digits=5)
print(with(Descriptives, corr.test(townsend, as.numeric(mentalhealth),
                                   use = "pairwise.complete.obs", method = "spearman")), digits=3)

#Self rated health
print(with(Descriptives, corr.test(srh, as.numeric(binaryinternet),
                                   use = "pairwise.complete.obs", method = "spearman")), digits=3)
print(with(Descriptives, corr.test(srh, as.numeric(binarynervous),
                                   use = "pairwise.complete.obs", method = "spearman")), digits=3)
print(with(Descriptives, corr.test(srh, as.numeric(getshelp),
                                   use = "pairwise.complete.obs", method = "spearman")), digits=3)
print(with(Descriptives, corr.test(srh, as.numeric(pasttime),
                                   use = "pairwise.complete.obs", method = "spearman")), digits=3)
print(with(Descriptives, corr.test(srh, as.numeric(lonely),
                                   use = "pairwise.complete.obs", method = "spearman")), digits=3)
print(with(Descriptives, corr.test(srh, as.numeric(changephys),
                                   use = "pairwise.complete.obs", method = "spearman")), digits=3)
print(with(Descriptives, corr.test(srh, as.numeric(routine),
                                   use = "pairwise.complete.obs", method = "spearman")), digits=3)
print(with(Descriptives, corr.test(srh, as.numeric(leavehouse),
                                   use = "pairwise.complete.obs", method = "spearman")), digits=5)
print(with(Descriptives, corr.test(srh, as.numeric(physicalhealth),
                                   use = "pairwise.complete.obs", method = "spearman")), digits=13)
print(with(Descriptives, corr.test(srh, as.numeric(mentalhealth),
                                   use = "pairwise.complete.obs", method = "spearman")), digits=5)

#HADS Anxiety symptoms
print(with(Descriptives, corr.test(HADSA, as.numeric(binaryinternet),
                                   use = "pairwise.complete.obs", method = "spearman")), digits=4)
print(with(Descriptives, corr.test(HADSA, as.numeric(binarynervous),
                                   use = "pairwise.complete.obs", method = "spearman")), digits=5)
print(with(Descriptives, corr.test(HADSA, as.numeric(getshelp),
                                   use = "pairwise.complete.obs", method = "spearman")), digits=3)
print(with(Descriptives, corr.test(HADSA, as.numeric(pasttime),
                                   use = "pairwise.complete.obs", method = "spearman")), digits=5)
print(with(Descriptives, corr.test(HADSA, as.numeric(lonely),
                                   use = "pairwise.complete.obs", method = "spearman")), digits=8)
print(with(Descriptives, corr.test(HADSA, as.numeric(physactivity),
                                   use = "pairwise.complete.obs", method = "spearman")), digits=5)
print(with(Descriptives, corr.test(HADSA, as.numeric(routine),
                                   use = "pairwise.complete.obs", method = "spearman")), digits=3)
print(with(Descriptives, corr.test(HADSA, as.numeric(leavehouse),
                                   use = "pairwise.complete.obs", method = "spearman")), digits=3)
print(with(Descriptives, corr.test(HADSA, as.numeric(physicalhealth),
                                   use = "pairwise.complete.obs", method = "spearman")), digits=3)
print(with(Descriptives, corr.test(HADSA, as.numeric(mentalhealth),
                                   use = "pairwise.complete.obs", method = "spearman")), digits=7)

#HADS Depression Symptoms
print(with(Descriptives, corr.test(HADSD, as.numeric(binaryinternet),
                                   use = "pairwise.complete.obs", method = "spearman")), digits=3)
print(with(Descriptives, corr.test(HADSD, as.numeric(binarynervous),
                                   use = "pairwise.complete.obs", method = "spearman")), digits=3)
print(with(Descriptives, corr.test(HADSD, as.numeric(getshelp),
                                   use = "pairwise.complete.obs", method = "spearman")), digits=3)
print(with(Descriptives, corr.test(HADSD, as.numeric(pasttime),
                                   use = "pairwise.complete.obs", method = "spearman")), digits=3)
print(with(Descriptives, corr.test(HADSD, as.numeric(lonely),
                                   use = "pairwise.complete.obs", method = "spearman")), digits=8)
print(with(Descriptives, corr.test(HADSD, as.numeric(changephys),
                                   use = "pairwise.complete.obs", method = "spearman")), digits=3)
print(with(Descriptives, corr.test(HADSD, as.numeric(routine),
                                   use = "pairwise.complete.obs", method = "spearman")), digits=3)
print(with(Descriptives, corr.test(HADSD, as.numeric(leavehouse),
                                   use = "pairwise.complete.obs", method = "spearman")), digits=3)
print(with(Descriptives, corr.test(HADSD, as.numeric(physicalhealth),
                                   use = "pairwise.complete.obs", method = "spearman")), digits=5)
print(with(Descriptives, corr.test(HADSD, as.numeric(mentalhealth),
                                   use = "pairwise.complete.obs", method = "spearman")), digits=4)

#Personality - emotional stability
print(with(Descriptives, corr.test(emo, as.numeric(binaryinternet),
                                   use = "pairwise.complete.obs", method = "spearman")), digits=3)
print(with(Descriptives, corr.test(emo, as.numeric(binarynervous),
                                   use = "pairwise.complete.obs", method = "spearman")), digits=7)
print(with(Descriptives, corr.test(emo, as.numeric(getshelp),
                                   use = "pairwise.complete.obs", method = "spearman")), digits=3)
print(with(Descriptives, corr.test(emo, as.numeric(pasttime),
                                   use = "pairwise.complete.obs", method = "spearman")), digits=7)
print(with(Descriptives, corr.test(emo, as.numeric(lonely),
                                   use = "pairwise.complete.obs", method = "spearman")), digits=8)
print(with(Descriptives, corr.test(emo, as.numeric(changephys),
                                   use = "pairwise.complete.obs", method = "spearman")), digits=3)
print(with(Descriptives, corr.test(emo, as.numeric(routine),
                                   use = "pairwise.complete.obs", method = "spearman")), digits=3)
print(with(Descriptives, corr.test(emo, as.numeric(leavehouse),
                                   use = "pairwise.complete.obs", method = "spearman")), digits=3)
print(with(Descriptives, corr.test(emo, as.numeric(physicalhealth),
                                   use = "pairwise.complete.obs", method = "spearman")), digits=5)
print(with(Descriptives, corr.test(emo, as.numeric(mentalhealth),
                                   use = "pairwise.complete.obs", method = "spearman")), digits=9)

#Personality - conscientiousness
print(with(Descriptives, corr.test(CON, as.numeric(binaryinternet),
                                   use = "pairwise.complete.obs", method = "spearman")), digits=3)
print(with(Descriptives, corr.test(CON, as.numeric(binarynervous),
                                   use = "pairwise.complete.obs", method = "spearman")), digits=3)
print(with(Descriptives, corr.test(CON, as.numeric(getshelp),
                                   use = "pairwise.complete.obs", method = "spearman")), digits=3)
print(with(Descriptives, corr.test(CON, as.numeric(pasttime),
                                   use = "pairwise.complete.obs", method = "spearman")), digits=3)
print(with(Descriptives, corr.test(CON, as.numeric(lonely),
                                   use = "pairwise.complete.obs", method = "spearman")), digits=3)
print(with(Descriptives, corr.test(CON, as.numeric(changephys),
                                   use = "pairwise.complete.obs", method = "spearman")), digits=3)
print(with(Descriptives, corr.test(CON, as.numeric(routine),
                                   use = "pairwise.complete.obs", method = "spearman")), digits=3)
print(with(Descriptives, corr.test(CON, as.numeric(leavehouse),
                                   use = "pairwise.complete.obs", method = "spearman")), digits=3)
print(with(Descriptives, corr.test(CON, as.numeric(physicalhealth),
                                   use = "pairwise.complete.obs", method = "spearman")), digits=3)
print(with(Descriptives, corr.test(CON, as.numeric(mentalhealth),
                                   use = "pairwise.complete.obs", method = "spearman")), digits=3)


#Personality - Extraversion
print(with(Descriptives, corr.test(ex, as.numeric(binaryinternet),
                                   use = "pairwise.complete.obs", method = "spearman")), digits=3)
print(with(Descriptives, corr.test(ex, as.numeric(binarynervous),
                                   use = "pairwise.complete.obs", method = "spearman")), digits=3)
print(with(Descriptives, corr.test(ex, as.numeric(getshelp),
                                   use = "pairwise.complete.obs", method = "spearman")), digits=3)
print(with(Descriptives, corr.test(ex, as.numeric(pasttime),
                                   use = "pairwise.complete.obs", method = "spearman")), digits=3)
print(with(Descriptives, corr.test(ex, as.numeric(lonely),
                                   use = "pairwise.complete.obs", method = "spearman")), digits=3)
print(with(Descriptives, corr.test(ex, as.numeric(changephys),
                                   use = "pairwise.complete.obs", method = "spearman")), digits=3)
print(with(Descriptives, corr.test(ex, as.numeric(routine),
                                   use = "pairwise.complete.obs", method = "spearman")), digits=3)
print(with(Descriptives, corr.test(ex, as.numeric(leavehouse),
                                   use = "pairwise.complete.obs", method = "spearman")), digits=3)
print(with(Descriptives, corr.test(ex, as.numeric(physicalhealth),
                                   use = "pairwise.complete.obs", method = "spearman")), digits=4)
print(with(Descriptives, corr.test(ex, as.numeric(mentalhealth),
                                   use = "pairwise.complete.obs", method = "spearman")), digits=3)


# Regressions -------------------------------------------------------------

# Binary logistic regression ----------------------------------------------
Descriptives <- filter(Descriptives, attendw5 == "Yes")

# Outcome = internet
class(Descriptives$binaryinternet)
Descriptives$binaryinternet <- factor(Descriptives$binaryinternet, ordered = FALSE)
levels(Descriptives$binaryinternet)
summary(Descriptives$binaryinternet)

internet1 <- glm (formula = relevel(binaryinternet, "same internet") ~ + scale(agedays_CIQ) + sex, data = Descriptives, family = binomial, na.action = na.omit)
summary(internet1)
anova(internet1, test = "Chisq")
exp(coef(internet1))
exp(confint(internet1))


internet2 <- glm (formula = relevel(binaryinternet, "same internet") ~ + scale(agedays_CIQ) + sex + covidalone, data = Descriptives, family = binomial, na.action = na.omit)
summary(internet2)
anova(internet2, test = "Chisq")
exp(coef(internet2))
exp(confint(internet2))

internet3 <- glm (formula = relevel(binaryinternet, "same internet") ~ + scale(agedays_CIQ) + sex + covidalone + scale(g), data = Descriptives, family = binomial, na.action = na.omit)
summary(internet3)
anova(internet3, test = "Chisq")
exp(coef(internet3))
exp(confint(internet3))

internet4 <- glm (formula = relevel(binaryinternet, "same internet") ~ + scale(agedays_CIQ) + sex + covidalone + scale(g) + scale(HADSA), data = Descriptives, family = binomial, na.action = na.omit)
summary(internet4)
anova(internet4, test = "Chisq")
exp(coef(internet4))
exp(confint(internet4))

# Outcome = covid-related stress
class(Descriptives$binarynervous)
Descriptives$binarynervous <- factor(Descriptives$binarynervous, ordered = FALSE)
levels(Descriptives$binarynervous)
summary(Descriptives$binarynervous)

#glm
nervous1 <- glm (formula = relevel(binarynervous, "never nervous") ~ + scale(agedays_CIQ) + sex, data = Descriptives, family = binomial, na.action = na.omit)
summary(nervous1)
anova(nervous1, test = "Chisq")
exp(coef(nervous1))
exp(confint(nervous1))

nervous2 <- glm (formula = relevel(binarynervous, "never nervous") ~ + scale(agedays_CIQ) + sex + covidalone, data = Descriptives, family = binomial, na.action = na.omit)
summary(nervous2)
anova(nervous2, test = "Chisq")
exp(coef(nervous2))
exp(confint(nervous2))

nervous3 <- glm (formula = relevel(binarynervous, "never nervous") ~ + scale(agedays_CIQ) + sex + covidalone + scale(HADSA), data = Descriptives, family = binomial, na.action = na.omit)
summary(nervous3)
anova(nervous3, test = "Chisq")
exp(coef(nervous3))
exp(confint(nervous3))

nervous4 <- glm (formula = relevel(binarynervous, "never nervous") ~ + scale(agedays_CIQ) + sex + covidalone + scale(HADSA) + scale(emo), data = Descriptives, family = binomial, na.action = na.omit)
summary(nervous4)
anova(nervous4, test = "Chisq")
exp(coef(nervous4))
exp(confint(nervous4))


# Outcome = pastime
class(Descriptives$pasttime)
Descriptives$pasttime <- factor(Descriptives$pasttime, ordered = FALSE)
levels(Descriptives$pasttime)
summary(Descriptives$pasttime)

pastime1 <- glm (formula = relevel(pasttime, "no new hobby") ~ + scale(agedays_CIQ) + sex, data = Descriptives, family = binomial, na.action = na.omit)
summary(pastime1)
anova(pastime1 , test = "Chisq")
exp(coef(pastime1))
exp(confint(pastime1))

pastime2 <- glm (formula = relevel(pasttime, "no new hobby") ~ + scale(agedays_CIQ) + sex + scale(GHL), data = Descriptives, family = binomial, na.action = na.omit)
summary(pastime2)
anova(pastime2 , test = "Chisq")
exp(coef(pastime2))
exp(confint(pastime2))

# Outcome = lonely
class(Descriptives$lonely)
Descriptives$lonely <- factor(Descriptives$lonely, ordered = FALSE)
levels(Descriptives$lonely)
summary(Descriptives$lonely)

lonely1 <- glm (formula = relevel(lonely, "none of the time") ~ + scale(agedays_CIQ) + sex, data = Descriptives, family = binomial, na.action = na.omit)
summary(lonely1)
anova(lonely1 , test = "Chisq")
exp(coef(lonely1))
exp(confint(lonely1))

lonely2 <- glm (formula = relevel(lonely, "none of the time") ~ + scale(agedays_CIQ) + sex + covidalone, data = Descriptives, family = binomial, na.action = na.omit)
summary(lonely2)
anova(lonely2 , test = "Chisq")
exp(coef(lonely2))
exp(confint(lonely2))

Descriptives$srh <- factor(Descriptives$srh, ordered = FALSE)

lonely3 <- glm (formula = relevel(lonely, "none of the time") ~ + scale(agedays_CIQ) + sex + covidalone + scale(townsend) + relevel(srh, "poor"), data = Descriptives, family = binomial, na.action = na.omit)
summary(lonely3)
anova(lonely3 , test = "Chisq")
exp(coef(lonely3))
exp(confint(lonely3))

lonely4 <- glm (formula = relevel(lonely, "none of the time") ~ + scale(agedays_CIQ) + sex + covidalone + scale(townsend) + relevel(srh, "poor") + scale(HADSA), data = Descriptives, family = binomial, na.action = na.omit)
summary(lonely4)
anova(lonely4, test = "Chisq")
exp(coef(lonely4))
exp(confint(lonely4))

lonely5 <- glm (formula = relevel(lonely, "none of the time") ~ + scale(agedays_CIQ) + sex + covidalone + scale(townsend) + relevel(srh, "poor") + scale(HADSA) + scale(emo), data = Descriptives, family = binomial, na.action = na.omit)
summary(lonely5)
anova(lonely5, test = "Chisq")
exp(coef(lonely5))
exp(confint(lonely5))


# Ordinal regressions (physical activity) ---------------------------------

#Model 1 
changephys_m1 <- clm(changephys ~ scale(agedays_CIQ) + sex, data = Descriptives, link = "logit", 
                  na.action = na.omit, convergence = "warn")
summary(changephys_m1)
exp(changephys_m1$beta) #Odds Ratio 
exp(confint(changephys_m1)) #CIs

#The anova method produces an analysis of deviance (ANODE) tables also based on Wald ??2-tests 
#and provides tables with type I, II and III hypothesis tests using the SAS definitions.
#type II tests attempt to respect the principle of marginality and test each term
#after all others while ignoring higher order interactions.
Anova(changephys_m1, Method="Chisq", type=2)

#we can use drop1 to obtain likelihood ratio tests of the explanatory variables while 
#controlling for the remaining variables:
drop1(changephys_m1, test="Chi")

nominal_test(changephys_m1) #check for proportionality - little diff in logLiks, p>.05 - passes

plot(profile(changephys_m1, alpha=1e-4))#shows relative profile likelihoods for the regression parameters
#e.g. scaled age parameter values -0.3 to 0.4 well supported by data, values outside have little likelihood
scale_test(changephys_m1)
slice.changephys_m1 <- slice(changephys_m1, lambda = 5)
par(mfrow = c(2, 3))
plot(slice.changephys_m1) 
#log-likelihood function is fairly well behaved and relatively closely quadratic for most parameters

convergence(changephys_m1) #(0) successful convergence

#Model 2
changephys_m2 <- clm(changephys ~ scale(agedays_CIQ) + sex + scale(pptclass), data = Descriptives, link = "logit", 
                     na.action = na.omit, convergence = "warn")
summary(changephys_m2)
exp(changephys_m2$beta) #Odds Ratio
exp(confint(changephys_m2)) #CIs

convergence(changephys_m2) #(0) successful convergence
Anova(changephys_m2, Method="Chisq", type=2) #analysis of deviance (ANODE) tables

nominal_test(changephys_m2) #check for proportionality - little diff in logLiks, p>.05 - passes

plot(profile(changephys_m2, alpha=1e-4))#shows relative profile likelihoods for the regression parameters
scale_test(changephys_m2)
slice.changephys_m2 <- slice(changephys_m2, lambda = 5)
par(mfrow = c(2, 3))
plot(slice.changephys_m2) #log-likelihood function is fairly well behaved and relatively closely quadratic for most parameters

#Model 3
changephys_m3 <- clm(changephys ~ scale(agedays_CIQ) + sex + scale(pptclass) + scale(g), data = Descriptives, link = "logit", 
                     na.action = na.omit, convergence = "warn")
summary(changephys_m3)
exp(changephys_m3$beta) #Odds Ratio
exp(confint(changephys_m3)) #CIs

convergence(changephys_m3) #(0) successful convergence
Anova(changephys_m3, Method="Chisq", type=2) #analysis of deviance (ANODE) tables

nominal_test(changephys_m3) #check for proportionality - little diff in logLiks, p>.05 - passes
scale_test(changephys_m3) 

plot(profile(changephys_m3, alpha=1e-4))#shows relative profile likelihoods for the regression parameters
slice.changephys_m3 <- slice(changephys_m3, lambda = 5)
par(mfrow = c(2, 3))
plot(slice.changephys_m3) #log-likelihood function is well behaved and closely quadratic for most parameters


# Ordinal Regression (Daily Routine) --------------------------------------

#Model 1
routine_m1 <- clm(routine ~ scale(agedays_CIQ) + sex, data = Descriptives, link = "logit", 
                     na.action = na.omit, convergence = "warn")
summary(routine_m1)
exp(routine_m1$beta) #Odds Ratio
exp(confint(routine_m1)) #CIs

convergence(routine_m1) #(0) successful convergence
Anova(routine_m1, Method="Chisq", type=2) 

nominal_test(routine_m1) 
scale_test(routine_m1) 

plot(profile(routine_m1, alpha=1e-4))
slice.routine_m1 <- slice(routine_m1, lambda = 5)
par(mfrow = c(2, 3))
plot(slice.routine_m1) 

#Model 2
routine_m2 <- clm(routine ~ scale(agedays_CIQ) + sex+ covidalone, data = Descriptives, link = "logit", 
                  na.action = na.omit, convergence = "warn")
summary(routine_m2)
exp(routine_m2$beta) #Odds Ratio
exp(confint(routine_m2)) #CIs

convergence(routine_m2) #(0) successful convergence
Anova(routine_m2, Method="Chisq", type=2) 

nominal_test(routine_m2) 
scale_test(routine_m2) 

plot(profile(routine_m2, alpha=1e-4))
slice.routine_m2 <- slice(routine_m2, lambda = 5)
par(mfrow = c(2, 3))
plot(slice.routine_m2) 

#Model 3
routine_m3 <- clm(routine ~ scale(agedays_CIQ) + sex + covidalone + GHL, data = Descriptives, link = "logit", 
                  na.action = na.omit, convergence = "warn")
summary(routine_m3)
exp(routine_m3$beta) #Odds Ratio
exp(confint(routine_m3)) #CIs

convergence(routine_m3) #(0) successful convergence
Anova(routine_m3, Method="Chisq", type=2) 

nominal_test(routine_m3) 
scale_test(routine_m3) 

plot(profile(routine_m3, alpha=1e-4))
slice.routine_m3 <- slice(routine_m3, lambda = 5)
par(mfrow = c(2, 3))
plot(slice.routine_m3) 

# Ordinal regression (leaving house) --------------------------------------
Descriptives$leavehouse <- as.factor(Descriptives$leavehouse)
#model 1
leavehouse_m1 <- clm(leavehouse ~ scale(agedays_CIQ) + sex, data = Descriptives, link = "logit", 
                  na.action = na.omit, convergence = "warn")
summary(leavehouse_m1)
exp(leavehouse_m1$beta) #Odds Ratio
exp(confint(leavehouse_m1)) #CIs

convergence(leavehouse_m1) #(0) successful convergence
Anova(leavehouse_m1, Method="Chisq", type=2) 

nominal_test(leavehouse_m1) 
scale_test(leavehouse_m1) 

plot(profile(leavehouse_m1, alpha=1e-4))
slice.leavehouse_m1 <- slice(leavehouse_m1, lambda = 5)
par(mfrow = c(2, 3))
plot(slice.leavehouse_m1) 

#model 2
leavehouse_m2 <- clm(leavehouse ~ scale(agedays_CIQ) + sex + scale(pptclass), data = Descriptives, link = "logit", 
                     na.action = na.omit, convergence = "warn")
summary(leavehouse_m2)
exp(leavehouse_m2$beta) #Odds Ratio
exp(confint(leavehouse_m2)) #CIs

convergence(leavehouse_m2) #(0) successful convergence
Anova(leavehouse_m2, Method="Chisq", type=2) 

nominal_test(leavehouse_m2) 
scale_test(leavehouse_m2) 

plot(profile(leavehouse_m2, alpha=1e-4))
slice.leavehouse_m2 <- slice(leavehouse_m2, lambda = 5)
par(mfrow = c(2, 3))
plot(slice.leavehouse_m2) 

#model 3
leavehouse_m3 <- clm(leavehouse ~ scale(agedays_CIQ) + sex + scale(pptclass) + scale(g), data = Descriptives, link = "logit", 
                     na.action = na.omit, convergence = "warn")
summary(leavehouse_m3)
exp(leavehouse_m3$beta) #Odds Ratio
exp(confint(leavehouse_m3)) #CIs

convergence(leavehouse_m3) #(0) successful convergence
Anova(leavehouse_m3, Method="Chisq", type=2) 

nominal_test(leavehouse_m3) 
scale_test(leavehouse_m3) 

plot(profile(leavehouse_m3, alpha=1e-4))
slice.leavehouse_m3 <- slice(leavehouse_m3, lambda = 5)
par(mfrow = c(2, 3))
plot(slice.leavehouse_m3) 

#model 4
leavehouse_m4 <- clm(leavehouse ~ scale(agedays_CIQ) + sex + scale(pptclass) + scale(g) + disease + scale(townsend) + srh, data = Descriptives, link = "logit", 
                     na.action = na.omit, convergence = "warn")
summary(leavehouse_m4)
exp(leavehouse_m4$beta) #Odds Ratio
exp(confint(leavehouse_m4)) #CIs

convergence(leavehouse_m4) #(0) successful convergence
Anova(leavehouse_m4, Method="Chisq", type=2) 

nominal_test(leavehouse_m4) 
scale_test(leavehouse_m4) 

plot(profile(leavehouse_m4, alpha=1e-4))
slice.leavehouse_m4 <- slice(leavehouse_m4, lambda = 5)
par(mfrow = c(2, 3))
plot(slice.leavehouse_m4) 


# Ordinal regression (physical health) ------------------------------------

#model 1
physhealth_m1 <- clm(physhealthafter ~ scale(agedays_CIQ) + sex, data = Descriptives, link = "logit", 
                       na.action = na.omit, convergence = "warn")
summary(physhealth_m1)
exp(physhealth_m1$beta) #Odds Ratio
exp(confint(physhealth_m1)) #CIs

convergence(physhealth_m1) #(0) successful convergence
Anova(physhealth_m1, Method="Chisq", type=2) 

nominal_test(physhealth_m1) 
scale_test(physhealth_m1)

plot(profile(physhealth_m1, alpha=1e-4))
slice.physhealth_m1 <- slice(physhealth_m1, lambda = 5)
par(mfrow = c(2, 3))
plot(slice.physhealth_m1) 

#model 2
physhealth_m2 <- clm(physhealthafter ~ scale(agedays_CIQ) + sex + scale(g), data = Descriptives, link = "logit", 
                     na.action = na.omit, convergence = "warn")
summary(physhealth_m2)
exp(physhealth_m2$beta) #Odds Ratio
exp(confint(physhealth_m2)) #CIs

convergence(physhealth_m2) #(0) successful convergence
Anova(physhealth_m2, Method="Chisq", type=2) 

nominal_test(physhealth_m2) 
scale_test(physhealth_m2)

plot(profile(physhealth_m2, alpha=1e-4))
slice.physhealth_m2 <- slice(physhealth_m2, lambda = 5)
par(mfrow = c(2, 3))
plot(slice.physhealth_m2) 

#model 3
physhealth_m3 <- clm(physhealthafter ~ scale(agedays_CIQ) + sex + scale(g) + disease + scale(townsend) + srh, data = Descriptives, link = "logit", 
                     na.action = na.omit, convergence = "warn")
summary(physhealth_m3)
exp(physhealth_m3$beta) #Odds Ratio
exp(confint(physhealth_m3)) #CIs

convergence(physhealth_m3) #(0) successful convergence
Anova(physhealth_m3, Method="Chisq", type=2) 

nominal_test(physhealth_m3) 
scale_test(physhealth_m3)

plot(profile(physhealth_m3, alpha=1e-4))
slice.physhealth_m3 <- slice(physhealth_m3, lambda = 5)
par(mfrow = c(2, 3))
plot(slice.physhealth_m3) 

#model 4
physhealth_m4 <- clm(physhealthafter ~ scale(agedays_CIQ) + sex + scale(g) + disease + scale(townsend) + srh + scale(HADSA) + scale(HADSD), data = Descriptives, link = "logit", 
                     na.action = na.omit, convergence = "warn")
summary(physhealth_m4)
exp(physhealth_m4$beta) #Odds Ratio
exp(confint(physhealth_m4)) #CIs

convergence(physhealth_m4) #(0) successful convergence
Anova(physhealth_m4, Method="Chisq", type=2) 

nominal_test(physhealth_m4) 
scale_test(physhealth_m4)

plot(profile(physhealth_m4, alpha=1e-4))
slice.physhealth_m4 <- slice(physhealth_m4, lambda = 5)
par(mfrow = c(2, 3))
plot(slice.physhealth_m4) 

#model 5
physhealth_m5 <- clm(physhealthafter ~ scale(agedays_CIQ) + sex + scale(g) + disease + scale(townsend) + srh + scale(HADSA) + scale(HADSD) + scale(emo) + scale(CON) + scale(ex), data = Descriptives, link = "logit", 
                     na.action = na.omit, convergence = "warn")
summary(physhealth_m5)
exp(physhealth_m5$beta) #Odds Ratio
exp(confint(physhealth_m5)) #CIs

convergence(physhealth_m5) #(0) successful convergence
Anova(physhealth_m5, Method="Chisq", type=2) 

nominal_test(physhealth_m5) 
scale_test(physhealth_m5)

plot(profile(physhealth_m5, alpha=1e-4))
slice.physhealth_m5 <- slice(physhealth_m5, lambda = 5)
par(mfrow = c(2, 3))
plot(slice.physhealth_m5) 

# Ordinal regression (mental health) -----------------------------------

#model 1
Descriptives$mentalhealthafter <- as.factor(Descriptives$mentalhealthafter)
mentalhealth_m1 <- clm(mentalhealthafter ~ scale(agedays_CIQ) + sex, data = Descriptives, link = "logit", 
                     na.action = na.omit, convergence = "warn")
summary(mentalhealth_m1)
exp(mentalhealth_m1$beta) #Odds Ratio
exp(confint(mentalhealth_m1)) #CIs

convergence(mentalhealth_m1) #(0) successful convergence
Anova(mentalhealth_m1, Method="Chisq", type=2) 

nominal_test(mentalhealth_m1) 
scale_test(mentalhealth_m1)

plot(profile(mentalhealth_m1, alpha=1e-4))
slice.mentalhealth_m1 <- slice(mentalhealth_m1, lambda = 5)
par(mfrow = c(2, 3))
plot(slice.mentalhealth_m1) 

#model 2
mentalhealth_m2 <- clm(mentalhealthafter ~ scale(agedays_CIQ) + sex + covidalone, data = Descriptives, link = "logit", 
                       na.action = na.omit, convergence = "warn")
summary(mentalhealth_m2)
exp(mentalhealth_m2$beta) #Odds Ratio
exp(confint(mentalhealth_m2)) #CIs

convergence(mentalhealth_m2) #(0) successful convergence
Anova(mentalhealth_m2, Method="Chisq", type=2) 

nominal_test(mentalhealth_m2) 
scale_test(mentalhealth_m2) 

plot(profile(mentalhealth_m2, alpha=1e-4))
slice.mentalhealth_m2 <- slice(mentalhealth_m2, lambda = 5)
par(mfrow = c(2, 3))
plot(slice.mentalhealth_m2) 

#model 3
mentalhealth_m3 <- clm(mentalhealthafter ~ scale(agedays_CIQ) + sex + covidalone + disease + srh, data = Descriptives, link = "logit", 
                       na.action = na.omit, convergence = "warn")
summary(mentalhealth_m3)
exp(mentalhealth_m3$beta) #Odds Ratio
exp(confint(mentalhealth_m3)) #CIs

convergence(mentalhealth_m3) #(0) successful convergence
Anova(mentalhealth_m3, Method="Chisq", type=2) 

nominal_test(mentalhealth_m3) 
scale_test(mentalhealth_m3) 

plot(profile(mentalhealth_m3, alpha=1e-4))
slice.mentalhealth_m3 <- slice(mentalhealth_m3, lambda = 5)
par(mfrow = c(2, 3))
plot(slice.mentalhealth_m3) 

#model 4
mentalhealth_m4 <- clm(mentalhealthafter ~ scale(agedays_CIQ) + sex + covidalone + disease + srh + scale(HADSA) + scale(HADSD), data = Descriptives, link = "logit", 
                       na.action = na.omit, convergence = "warn")
summary(mentalhealth_m4)
exp(mentalhealth_m4$beta) #Odds Ratio
exp(confint(mentalhealth_m4)) #CIs

convergence(mentalhealth_m4) #(0) successful convergence
Anova(mentalhealth_m4, Method="Chisq", type=2) 

nominal_test(mentalhealth_m4) 
scale_test(mentalhealth_m4) 

plot(profile(mentalhealth_m4, alpha=1e-4))
slice.mentalhealth_m4 <- slice(mentalhealth_m4, lambda = 5)
par(mfrow = c(2, 3))
plot(slice.mentalhealth_m4) 

#model 5
mentalhealth_m5 <- clm(mentalhealthafter ~ scale(agedays_CIQ) + sex + covidalone + disease + srh + scale(HADSA) + scale(HADSD) + scale(emo) + scale(ex), data = Descriptives, link = "logit", 
                       na.action = na.omit, convergence = "warn")
summary(mentalhealth_m5)
exp(mentalhealth_m5$beta) #Odds Ratio
exp(confint(mentalhealth_m5)) #CIs

convergence(mentalhealth_m5) #(0) successful convergence
Anova(mentalhealth_m5, Method="Chisq", type=2) 

nominal_test(mentalhealth_m5) 
scale_test(mentalhealth_m5) 

plot(profile(mentalhealth_m5, alpha=1e-4))
slice.mentalhealth_m5 <- slice(mentalhealth_m5, lambda = 5)
par(mfrow = c(2, 3))
plot(slice.mentalhealth_m5) 


# Wilcoxon ----------------------------------------------------------------

Descriptives$physhealthafter <- as.numeric(Descriptives$physhealthafter)
Descriptives$physhealthbefore <- as.numeric(Descriptives$physhealthbefore)
wilcox.test(Descriptives$physhealthbefore, Descriptives$physhealthafter, 
            paired = TRUE, alternative = "two.sided", conf.int = T, na.action=na.omit, correct=FALSE)
Wilcoxon_PhysHealth<-wilcox.test(Descriptives$physhealthbefore, Descriptives$physhealthafter, paired = TRUE, alternative = "two.sided", conf.int = T, na.action=na.omit, correct=FALSE)
Z_PhysHealth<-qnorm(Wilcoxon_PhysHealth$p.value/2)
summary(table(Descriptives$physhealthbefore, Descriptives$physhealthafter))
abs(Z_PhysHealth)/sqrt(172)

Descriptives$mentalhealthafter <- as.numeric(Descriptives$mentalhealthafter)
Descriptives$mentalhealthbefore <- as.numeric(Descriptives$mentalhealthbefore)
wilcox.test(Descriptives$mentalhealthbefore, Descriptives$mentalhealthafter, 
            paired = TRUE, alternative = "two.sided", conf.int = T, na.action=na.omit, correct=FALSE)
Wilcoxon_MentalHealth<-wilcox.test(Descriptives$mentalhealthbefore, Descriptives$mentalhealthafter, paired = TRUE, alternative = "two.sided", conf.int = T, na.action=na.omit, correct=FALSE)
Z_MentalHealth<-qnorm(Wilcoxon_MentalHealth$p.value/2)
summary(table(Descriptives$mentalhealthafter, Descriptives$mentalhealthbefore))
abs(Z_MentalHealth)/sqrt(177)

summary(Descriptives$mentalhealthafter)
summary(Descriptives$physhealthafter)
        
        


# Correlation matrix ------------------------------------------------------
corrmat_data <- as.data.frame(Descriptives[c('agedays_CIQ', 'sex', 'pptclass', 'covidalone', 'g', 'GHL', 'disease',
                          'townsend', 'srh', 'HADSA', 'HADSD', 'emo', 'CON', 'ex', 'leavehouse', 'binaryinternet',
                          'getshelp', 'routine', 'physicalhealth', 'mentalhealth', 'binarynervous', 'lonely',
                          'physactivity', 'pasttime')])
head(corrmat_data) 
corrmat_data$binaryinternet <- as.numeric(corrmat_data$binaryinternet)
corrmat_data$getshelp <- as.numeric(corrmat_data$getshelp)
corrmat_data$routine <- as.numeric(corrmat_data$routine)
corrmat_data$binarynervous <- as.numeric(corrmat_data$binarynervous)
corrmat_data$lonely <- as.numeric(corrmat_data$lonely)
corrmat_data$pasttime <- as.numeric(corrmat_data$pasttime)
corrmat_data$leavehouse <- as.numeric(corrmat_data$leavehouse)
corrmat_data$srh <- as.numeric(corrmat_data$srh)
str(corrmat_data)

corrmat_s <- cor(corrmat_data, use="pairwise.complete.obs", method = "spearman")
corrmat_p <- cor(corrmat_data, use="pairwise.complete.obs", method = "pearson")
corrmat_s <- round(corrmat_s, 3)
corrmat_p <- round(corrmat_p, 3)
write.csv(corrmat_p, "corrmat_p.csv")
write.csv(corrmat_s, "corrmat_s.csv")
```
